# Effects of Different Exercise Interventions on Body Composition in Women with Overweight and Obesity: A Systematic Review and Network Meta-analysis

**DOI:** 10.1101/2025.10.19.25338160

**Authors:** Shiwei Song, Yincheng Wei, Haoze Zhang, Andrew Soundy

## Abstract

**Objective:** This study aimed to investigate the effects of different types of exercise on body composition in women with overweight or obesity and to compare the relative effectiveness of these interventions using a systematic review and network meta-analysis.

**Methods:** A systematic search of 7 electronic databases was performed from database inception to September 2025. Randomized controlled trials (RCTs) involving women with overweight and obesity were included according to predefined eligibility criteria. Interventions consisted of 7 different types of exercise, while comparators were no exercise or normal-lifestyle condition. The primary outcome measure was body fat percentage (BFP). Secondary outcome measures included, body mass index (BMI), waist circumference (WC), fat mass (FM), and lean body mass (LBM). Risk of bias assessment was undertaken. Network meta-analysis was performed using a random-effects model, and consistency was assessed with surface under the cumulative ranking curve (SUCRA) used for ranking the relative effects of exercise types. Certainty assessment was performed using the Confidence in Network Meta-Analysis (CINeMA) framework.

**Results:** A total of 44 randomized controlled trials comprising 2384 female participants were included. The network meta-analysis revealed that, compared to the control, multiple exercise modalities significantly reduced body fat percentage (BFP) in women with overweight or obesity. Aerobic exercise of vigorous intensity (AE(V)) was ranked as the most effective intervention for reducing BFP (SUCRA = 79.7), whereas resistance training (RT) demonstrated the weakest effect (SUCRA = 18.3). The effectiveness of exercise modalities exhibited distinct outcome-specific patterns: AE(V) was optimal for reducing body mass index (SUCRA = 93.9), AE(MV) was most beneficial for decreasing fat mass (SUCRA = 80.9), and waist circumference (SUCRA = 67.6). Furthermore, subgroup analysis by publication date indicated that the optimal exercise modality for reducing BFP has changed over time, with AE(V) being superior in recent studies and AE(M) in earlier studies.

**Conclusion:** The effects of exercise on body composition in women with overweight and obesity are outcome-specific. AE(V) may be most effective for reducing BMI and BFP, AE(MV) for improving WC and FM, while RT shows potential for enhancing LBM. The finding that the optimal modality for reducing BFP has changed over time highlights the need for contemporary, evidence-based prescriptions. These results provide a critical evidence base for tailoring exercise programs to individual patient goals in clinical practice.

## INTRODUCTION

Overweight and obesity among women have become pressing global public health concerns, given their strong association with a range of chronic conditions, including cardiovascular diseases.^1–5^ According to data from the World Health Organization (WHO), the prevalence of obesity among women has steadily increased over the past decades.^6,7^ Obesity in women is associated not only with elevated risks of cardiovascular disease, type 2 diabetes, and metabolic syndrome, but also with adverse effects on reproductive and mental health.^8^ Optimization of body composition including reductions in body mass index (BMI), body fat percentage (BFP), waist circumference (WC), and fat mass (FM), together with the preservation of lean body mass (LBM) is regarded as a key target for improving obesity-related health outcomes.^9^ As a non-pharmacological and cost-effective strategy, exercise interventions have been widely implemented in weight management among overweight and obese populations.^10^

Previous research has demonstrated that different types of exercise exert varying effects on body composition. For instance, aerobic exercise has been shown to enhance energy expenditure and fat oxidation, thereby contributing to reductions in body weight and body fat percentage.^11^ Alternatively, resistance training increases muscle mass, elevates basal metabolic rate, and prevents the loss of lean body mass during weight loss.^12^ High-intensity interval training has attracted increasing attention due to its time efficiency and pronounced metabolic effects.^13^ Moreover, combined exercise modalities, such as aerobic and resistance training, may provide greater benefits by simultaneously reducing fat mass and preserving lean body mass.^14^

Similarly, despite the growing number of randomized controlled trials investigating exercise interventions in overweight and obese populations, most existing meta-analyses have been limited to traditional pairwise comparisons. These analyses typically focus on direct comparisons between a few commonly studied modalities, such as aerobic exercise, resistance training, combined aerobic and resistance training, whole-body vibration training, and high-intensity interval training. However, they often fail to account for the broader landscape of available interventions. While such pairwise approaches have yielded valuable insights into the efficacy of individual exercise types, their scope remains inherently constrained. In particular, they are unable to incorporate indirect evidence from trials that do not share a common comparator, and they lack the capacity to generate a comprehensive ranking of intervention effectiveness.

For instance, Campa et al. (2020) investigated the effects of different resistance training frequencies on body composition in overweight and obese women, reporting improvements in lean mass and reductions in fat mass, but without comparing RT to other modalities such as HIIT or AE.^15^ Liu et al. (2022) synthesized evidence on various resistance exercise forms, yet their pairwise meta-analysis excluded indirect comparisons and lacked a unified ranking framework.^16^ Amare et al. (2024) compared AE, RT, and AE+RT in terms of fat mass and glucolipid metabolism, but did not include emerging modalities like WBVT or HIIT.^17^ Jayedi et al. (2024) conducted a dose-response meta-analysis of aerobic exercise, confirming linear reductions in body fat and waist circumference with increasing AE duration, yet did not assess comparative effectiveness across modalities.^18^

These fragmented findings highlight the limitations of pairwise meta-analytic approaches in synthesizing the full spectrum of exercise interventions. Without a unified analytical framework that accommodates both direct and indirect comparisons, it remains difficult to determine which exercise modality offers the greatest benefit for improving body composition outcomes.

Importantly, network meta-analysis (NMA) enables the integration of direct and indirect evidence, the ranking of multiple interventions, and the generation of more comprehensive evidence to inform clinical decision-making.^19^ Therefore, this study was designed to systematically evaluate the effects of different types of exercise on body composition in overweight or obese women using NMA, with the aim of identifying the most effective exercise strategies.

## METHODS

### Registration

This systematic review and NMA were conducted in accordance with the Preferred Reporting Items for Systematic Reviews and Meta-Analyses (PRISMA) guidelines,^20^ and the search reported according the PRISMA statement for reporting literature searches.^21^ The study protocol was prospectively registered in the International Prospective Register of Systematic Reviews (PROSPERO; ID: CRD420251161064).

### Literature search strategy

A systematic electronic search was conducted by two authors independently. A third author was available for arbitration. A total of 7 electronic databases were searched from inception until September 2025 including; PubMed, Embase, CINAHL, SportsDicus, the Cochrane Library, Web of Science, and Scopus. Medical Subject Headings (MeSH) and Emtree terms were used for keyword matching respectively. Keywords used for searching included terms (“adipose tissue hyperplasia” OR adipositas OR adiposity OR “alimentary obesity” OR “body weight, excess” OR corpulency OR “fat overload” OR “syndrome nutritional” OR obesity OR obesitas OR overweigh) AND (females OR woman OR women OR female) AND (Exercises OR “Exercise, Physical” OR “Exercises, Physical” OR “Physical Exercise” OR “Physical Exercises” OR “Exercise, Aerobic” OR “Aerobic Exercise” OR “Aerobic Exercises” OR “Exercises, Aerobic” OR “Exercise Training” OR “Exercise Trainings” OR “Training, Exercise” OR “Trainings, Exercise” OR “Physical Activity” OR “Activities, Physical” OR “Activity, Physical” OR “Physical Activities” OR “Tai Chi” OR “resistance training” OR “strength training” OR “combined training” OR Qigong OR “Whole-Body Vibration Training” OR Baduanjin) AND (“body component” OR “lean body mass” OR “body weight” OR “body adiposity index” OR “body fat” OR “body fat percentage”) AND (“randomized controlled trial” OR randomized OR placebo). Detailed search strategies for each database are provided in Appendix S2. The search strategy was developed according to the core elements of the PICOS framework: (P) Population: women with overweight or obesity; (I) Intervention: exercise; (C) Comparator: no exercise or normal-lifestyle condition; (O) Outcomes: the primary outcome was BFP, with secondary outcomes including LBM, BMI, WC, and FM; and (S) Study design: RCTs.

In addition, electronic searches of two search engines was performed for the first 30 pages of results including: Google Scholar and ScienceDirect. Grey literature was searched using ProQuest and GreyMatters. Citation chasing was performed on all included articles.

### Eligibility criteria

Studies were included if they met the following criteria set out according to the PICOS framework with a section for other criteria added.

Population (P): participants were overweight and/or obese, defined as BMI > 25 kg/m² (overweight) and/or BMI ≥ 30 kg/m² (obese) for European populations, and BMI ≥ 24 kg/m² (overweight) and/or BMI ≥ 28 kg/m² (obese) for Asian populations. If BMI data were unavailable, BFP was used as the criterion (female BFP ≥ 30), and participants had to be adult women aged 18–65 years. For studies including older women (>65 years), data had to be reported separately, and the older subgroup could not exceed 30% of the total sample to minimize confounding effects of age on body composition. Studies that combined exercise interventions with other interventions, unless control and exercise groups without caloric restriction were included. Animal studies were excluded.

Intervention (I): the intervention group received at least 8 weeks of structured exercise training. For the purpose of this review exercise was generally defined as physical activity that is structured, planned and repetitive and has a goal of improvement or maintenance of an individual’s physical fitness^22^. Sub-classification of exercise acceptable for this review are identified in Table 1. Studies were excluded if they reported only the acute effects of a single exercise session.

**Table 1.**
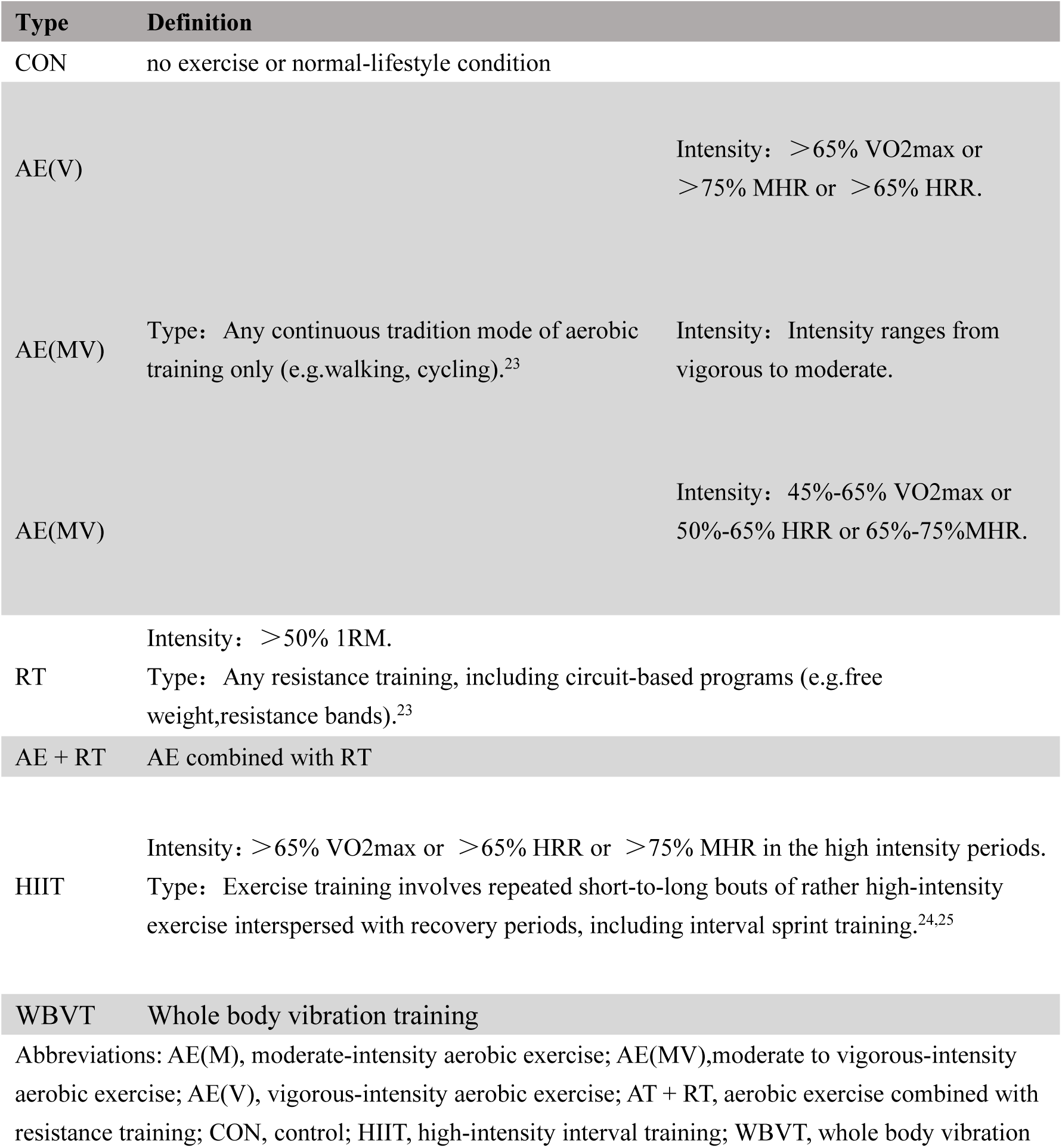
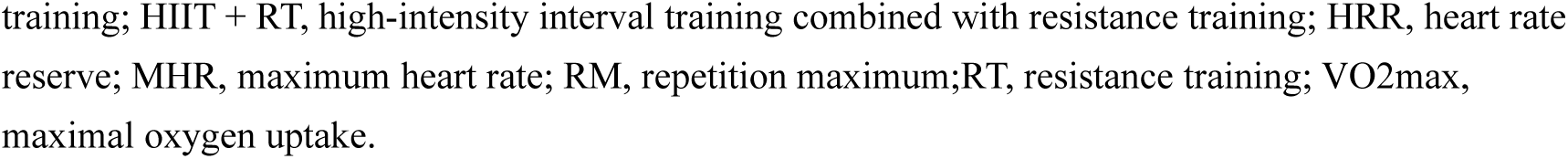
The classifications of exercise training.

| Type | Definition |
| --- | --- |
| CON | no exercise or normal-lifestyle condition |
| AE(V) | Intensity: >65% VO<sub>2</sub>max or >75% MHR or >65% HRR. |
| AE(MV) | Type: Any continuous tradition mode of aerobic training only (e.g. walking, cycling).<sup>23</sup> Intensity: Intensity ranges from vigorous to moderate. |
| AE(MV) | Intensity: 45%-65% VO<sub>2</sub>max or 50%-65% HRR or 65%-75%MHR. |
| RT | Intensity: >50% 1RM.<br>Type: Any resistance training, including circuit-based programs (e.g. free weight, resistance bands).<sup>23</sup> |
| AE + RT | AE combined with RT |
| HIIT | Intensity: >65% VO<sub>2</sub>max or >65% HRR or >75% MHR in the high intensity periods.<br>Type: Exercise training involves repeated short-to-long bouts of rather high-intensity exercise interspersed with recovery periods, including interval sprint training.<sup>24,25</sup> |
| WBVT | Whole body vibration training |

Abbreviations: AE(M), moderate-intensity aerobic exercise; AE(MV), moderate to vigorous-intensity aerobic exercise; AE(V), vigorous-intensity aerobic exercise; AT + RT, aerobic exercise combined with resistance training; CON, control; HIIT, high-intensity interval training; WBVT, whole body vibration
training; HIIT + RT, high-intensity interval training combined with resistance training; HRR, heart rate reserve; MHR, maximum heart rate; RM, repetition maximum; RT, resistance training; VO<sub>2</sub>max, maximal oxygen uptake.

Comparator (C): Studies required an active or inactive control group. The control group could involve no exercise or normal-lifestyle condition.

Outcomes (O): Studies had to assess changes in body fat percentage before and after the intervention were assessed, with no restriction on measurement methods.

Study design (S): Only randomized controlled trials were included. Studies were excluded if they were qualitative in nature, a literature review, editorial letters, or abstracts from conference proceedings.

Other criteria: Studies were excluded if they were duplicate publications. No date restriction was placed on studies. Only English-language RCTs published up to September 2025 were included.

Two researchers independently screened all retrieved studies according to the predefined inclusion and exclusion criteria. A third reviewer was available for arbitration if required. Multiple reports from the same trial were consolidated, with the first or most comprehensive report selected as the primary reference.

### Exercise categories

Eight categories were used to classify the exercise interventions for the included RCTs:

1. Control group: no exercise or normal-lifestyle condition (CON)
2. RT
3. AE combined with RT (AT + RT)
4. HIIT
5. AE, moderate intensity (AE[M])
6. AE, vigorous intensity (AE[V])
7. AE, moderate to vigorous intensity (AE[MV])
8. WBVT

Vigorous exercise intensity is defined as a maximal oxygen uptake (VO2max) >65%, heart rate reserve (HRR) >65%, or maximum heart rate (MHR) >75%, while moderate exercise corresponds to 45%-65% VO2max, 50%-65% HRR, or >65%-75% MHR.^23^ Table 1 provides a detailed listing of the definitions for each exercise category.

### Data extraction

Data were independently extracted by two researchers, and any discrepancies were resolved through discussion, with a third author consulted if necessary. Extracted information included the first author, year of publication, country, participant characteristics (number of participants in experimental and control groups, sex, age, body fat percentage, body mass index, lean body mass, fat mass, and waist circumference), intervention details (type of exercise, intensity, duration, frequency, intervention period, and whether supervised or unsupervised), as well as outcome measurement methods and units. When data were incomplete, corresponding authors were contacted via email to obtain the missing information.

### Risk of bias and CINeMA assessment

The risk of bias (ROB) of included studies was independently assessed by two researchers using the Cochrane Risk of Bias Assessment Tool,^26^ which evaluates seven domains: (a) random sequence generation, (b) allocation concealment, (c) blinding of participants and personnel, (d) blinding of outcome assessment, (e) incomplete outcome data, (f) selective reporting, and (g) other sources of bias. Because participant blinding is difficult to implement in exercise interventions, this domain was excluded from the overall ROB scoring; instead, the blinding of personnel in outcome assessment was used as a quality criterion. A composite scoring method was applied to classify each study: studies with no high-risk domains and no more than three unclear-risk domains were considered low risk; studies with at least one high-risk domain, or with four or more unclear-risk domains in the absence of high-risk domains, were classified as moderate risk; all other studies were considered high risk.^27^

The certainty of evidence for primary and secondary outcome network estimates was assessed using the Confidence in Network Meta-Analysis (CINeMA) framework, which encompasses recommendation grading, assessment, formulation, and evaluation.^28^

### Data synthesis and statistical analyses

The pre- and post-intervention change values for the experimental and control groups were combined to estimate the effect. The standard deviation (SD) of the changes was calculated according to the formula provided in the Cochrane Handbook for Systematic Reviews of Interventions (version 6.3).^29^

A random-effects multivariable non-parametric meta-analysis was performed using STATA 16.0 (StataCorp, College Station, TX, USA).^30^ Following the frequentist framework and current PRISMA guidelines for non-parametric meta-analyses, pooled estimates and 95% CIs were calculated.^20^ Owing to differences in outcome measurement tools across studies, standardized mean differences (SMDs) were used to assess the effect sizes of BFP, BMI, FM, LBM, and WC.

The relationships between exercise interventions were illustrated using a network evidence diagram, in which the lines connecting nodes represent direct comparisons between interventions, and the size of each node and the thickness of connecting lines are proportional to the number of studies. A network contribution plot was also generated to quantify the contribution of each direct comparison.

The transitivity assumption of NMA was evaluated by examining the inclusion criteria of individual studies, assessing whether all participants could theoretically be randomized to any intervention, and applying a consistency model.^31^ Transitivity, a core assumption of NMA, assumes that indirect comparisons can reliably reflect unobserved direct comparisons when effect modifiers are evenly distributed across studies.^30,32^ Consistency within each closed-loop was assessed by calculating inconsistency factors (IFs) with 95% CIs, with consistency indicated when the lower limit of the 95% CI included 0.^33^ An inconsistency model was applied to test for overall inconsistency. When inconsistency was not significant (p > 0.05), a consistency model was used.^34^ Local inconsistency was further evaluated using node-splitting analysis, and results were considered reliable when p > 0.05. The Surface Under the Cumulative Ranking curve (SUCRA) was applied to rank and compare the relative effects of different exercise interventions.^35^ SUCRA values range from 0 to 100, with higher values indicating greater efficacy; 100 represents the most effective intervention with no uncertainty, and 0 represents the least effective intervention with no uncertainty.^36^ To assess potential publication bias due to small-study effects, a network funnel plot was constructed and visually inspected for symmetry.

## RESULTS

### Literature selection

The flowchart of the study selection process is presented in Figure 1. A total of 17,351 articles were initially identified, 16,301 of which were from database and 1050 from the Google scholar and ScienceDirect. After removing 6,535 duplicates, 9,178 records remained for screening. During the title and abstract review, 8,894 articles were excluded. Full texts of the remaining 293 articles were assessed for eligibility, resulting in the exclusion of 250 studies. Ultimately, 44 articles were included in the quantitative synthesis, one of which was identified via an alternative screening approach.

**Figure 1.**
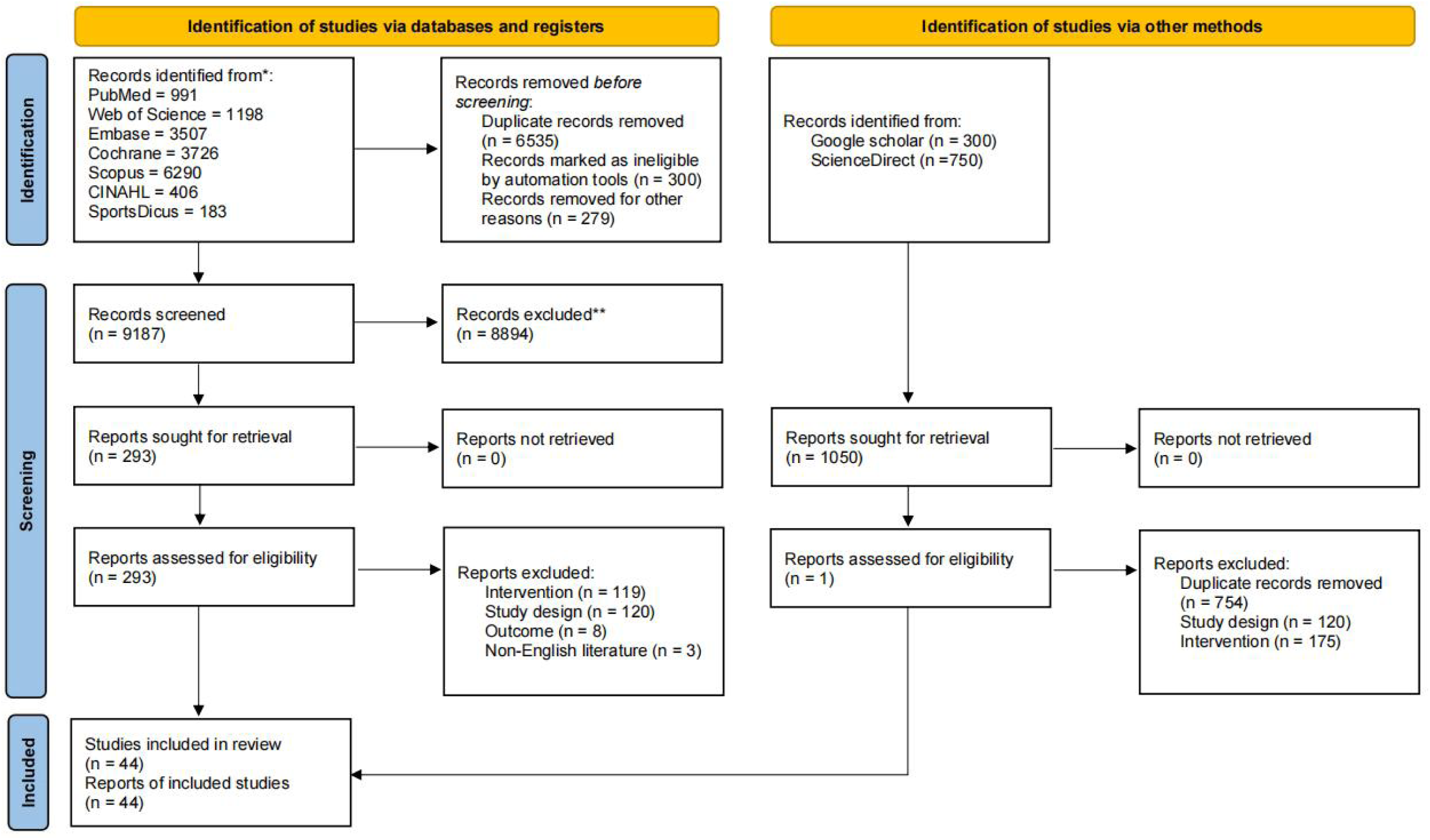
Preferred Reporting Items for Systematic Reviews and Meta-Analyses (PRISMA) flow diagram depicting the study selection process.

### Characteristics of the included studies

The characteristics of the included studies are summarized in Appendix S3, with a complete list provided in Appendix S4. These studies were published between 2006 and 2025 and conducted across North America (4/44, 9%) study), South America (3/44, 7%), Asia (16/44, 36%), Europe (19/44, 43%), and Africa (2 studies). The experimental groups comprised a total of 1,629 female participants with overweight or obesity, whereas the control groups included 755 participants. Participant ages ranged from 11 to 65 years. Baseline characteristics included age, BMI, BFP, FM and LBM.

Exercise interventions were categorized as AE(M), AE(MV), AE(V), RT, AE + RT, HIIT and WBVT. Several studies applied progressive overload principles, which limited the ability to clearly differentiate training intensity between RT and AE + RT groups. The durations reported in Appendix S3 excluded warm-up and cool-down sessions unless otherwise specified. On average, the intervention period was 12.1 weeks (SD = 5.2), ranging from 8 to 40 weeks, with over one-sixth (13.6%) of studies lasting longer than 12 weeks. Participants trained an average of 3.3 sessions per week (SD = 0.85), and all interventions were delivered under supervised conditions. Based on the forest plot, 2 studies in BFP and 1 study in BMI were excluded from the meta-analysis due to pronounced heterogeneity, as evidenced by outlying effect sizes and overly broad confidence intervals (Appendix 11).^37–39^

Regarding outcome assessment, BFP was measured using multiple methods, including bioelectrical impedance analysis (BIA), dual-energy X-ray absorptiometry (DEXA), skinfold thickness with Siri/Jackson formulas (ST), and hydrostatic weighing (HW). BIA devices from InBody, TANITA, and Omron Healthcare, as well as the electronic scale Ohaus, were employed across studies. Secondary outcome measures included BMI, LBM, FM, and WC.

### Results of ROB assessment

Details of the risk of bias (ROB) assessment for each study are provided in Appendix S5. A total of 28 (28/44, 64%) studies explicitly reported their randomisation methods. Regarding allocation procedures, 19 (19/44, 43%) studies demonstrated evidence of allocation concealment, and 23 (23/44, 52%) studies reported the use of blinding in outcome assessment. Additionally, 41 (41/44, 93%) studies were judged to have a low risk of selective reporting. For other sources of bias, studies were rated as high risk when they involved small sample sizes (fewer than 10 participants per group), lacked supervision mechanisms, or exhibited substantial measurement error in outcome assessment. Overall, 27 (27/44, 61%) studies were assessed as having a low risk of bias, 14 (14/44, 32%) as having some concerns, and 3 (3/44, 7%) as having a high risk of bias.

### Network meta-analysis

A network meta-analysis of BFP as the primary outcome was conducted, along with secondary outcomes including LBM, BMI, WC, and FM. This analysis was designed to examine whether the effects of different exercise modalities on secondary outcomes were consistent with changes in body fat percentage, thereby providing a more comprehensive evaluation of intervention effectiveness. In addition, subgroup analyses stratified by publication date were performed for primary outcomes to determine whether publication date influenced the treatment effects.

BFP supporting materials and secondary outcomes are shown as follows.

Figure 2 presents the network evidence diagram illustrating the network meta-analysis of included studies on the effects of exercise types on BFP and secondary outcomes (LBM, BMI, WC, and FM). Node size represents the sample size for each exercise modality, while the thickness of connecting lines indicates the number of studies comparing the respective interventions. AE(M) (13/44, 30%) was the most frequently investigated intervention, whereas WBVT (1/44, 2%) was the least common.

**Figure 2.**
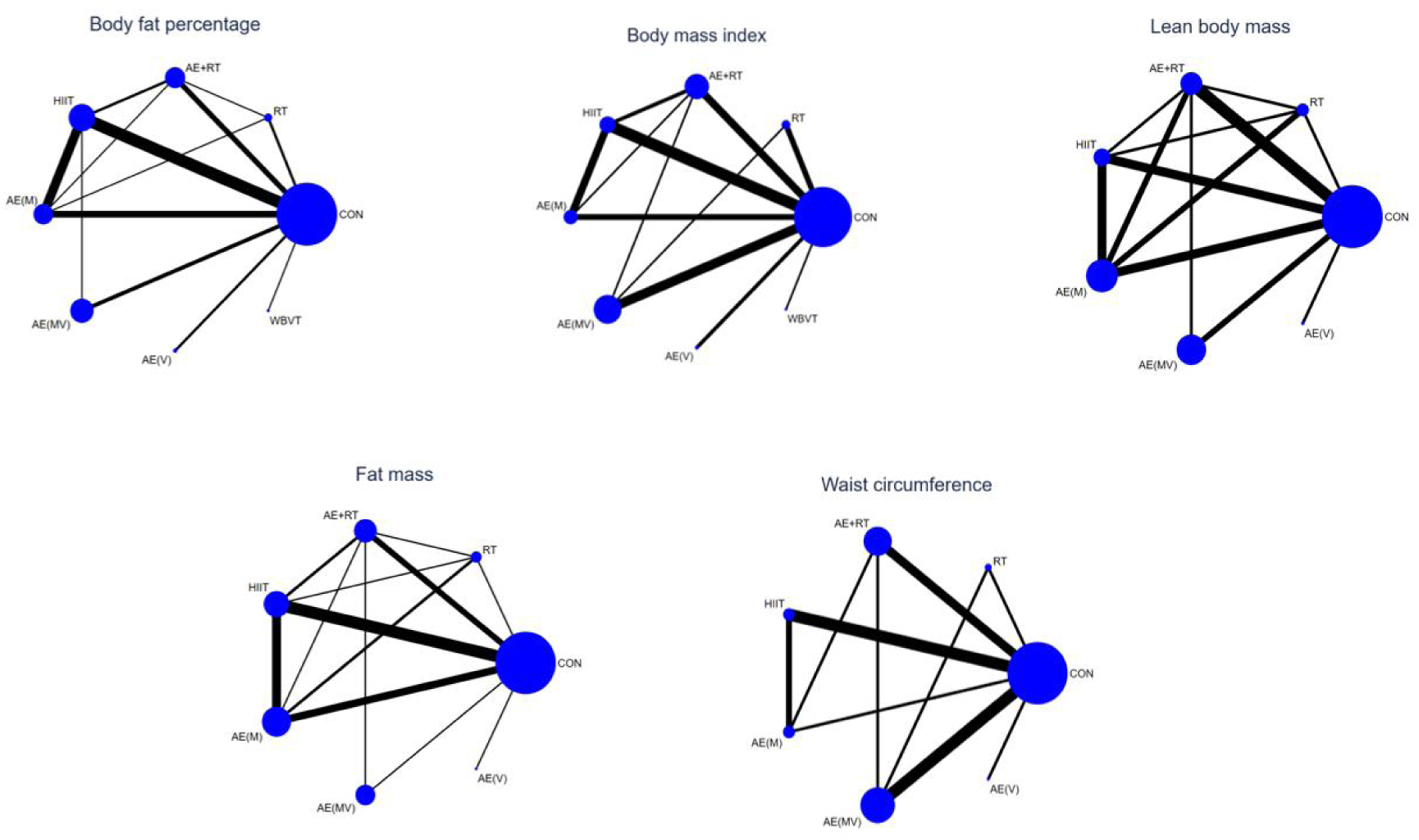
Network plot presenting the effects of different exercise type on body fat percentage and secondary outcomes in women with overweight and obesity.

Appendix S6 presents the network contribution plot, illustrating the contributions of direct and indirect comparisons to the NMA and the number of studies informing each direct comparison.

Inconsistency between BFP and secondary outcomes was assessed using loop-specific heterogeneity estimates, inconsistency models, and node-splitting analysis (Appendix S7). Loop-specific heterogeneity estimates indicated good consistency for all closed loops of BFP, BMI, and WC, whereas two loops were inconsistent for LBM and three loops were inconsistent for FM. The inconsistency model yielded p-values >0.05 for BFP, BMI, LBM, WC, and FM, indicating no significant inconsistency. Node-splitting analysis further confirmed that, overall, there was no inconsistency between direct and indirect evidence, supporting the reliability of the results.

Forest plots for eligible comparisons of BFP and secondary outcomes, including 95% CIs and 95% prediction intervals (95% PrIs), are presented in Appendix S8.

Funnel plots for BFP and secondary outcomes were constructed to assess potential publication bias in the NMA (Appendix S9). The plots were approximately symmetrical for all outcomes, suggesting a low likelihood of publication bias or small-study effects.

The SUCRA probabilities for each intervention in the network, for BFP and secondary outcomes, are presented in Appendix S10. Higher SUCRA values indicate a greater likelihood that an intervention ranks as the most effective within the network.

### Pooled estimates of primary outcomes

Table 2 and 3 present the results of the league table analysis for BFP. Compared with the control group, AE + RT (SMD = -0.84, 95% CI [-1.31, -0.37], p < 0.001), HIIT (SMD = -0.89, 95% CI [-1.24, -0.53], p < 0.001), AE(M) (SMD = -0.76, 95% CI [-1.19, -0.32], p < 0.001), AE(MV) (SMD = -0.87, 95% CI [-1.40, -0.34], p < 0.001), and AE(V) (SMD = -1.15, 95% CI [-2.05, -0.25], p < 0.001)significantly reduced BFP. SUCRA ranking indicated that AE(V) (SUCRA = 79.7) was most likely the optimal intervention for reducing BFP, followed by HIIT (SUCRA = 66.3), AE(MV) (SUCRA = 63.7), AE + RT (SUCRA = 60.7), WBVT (SUCRA = 54.2) and AE(M) (SUCRA = 52.2) whereas RT (SUCRA = 18.3) was the least effective intervention. Neither RT nor WBVT demonstrated statistically significant effects on BFP compared with control or other exercise modalities.

**Table 2.**
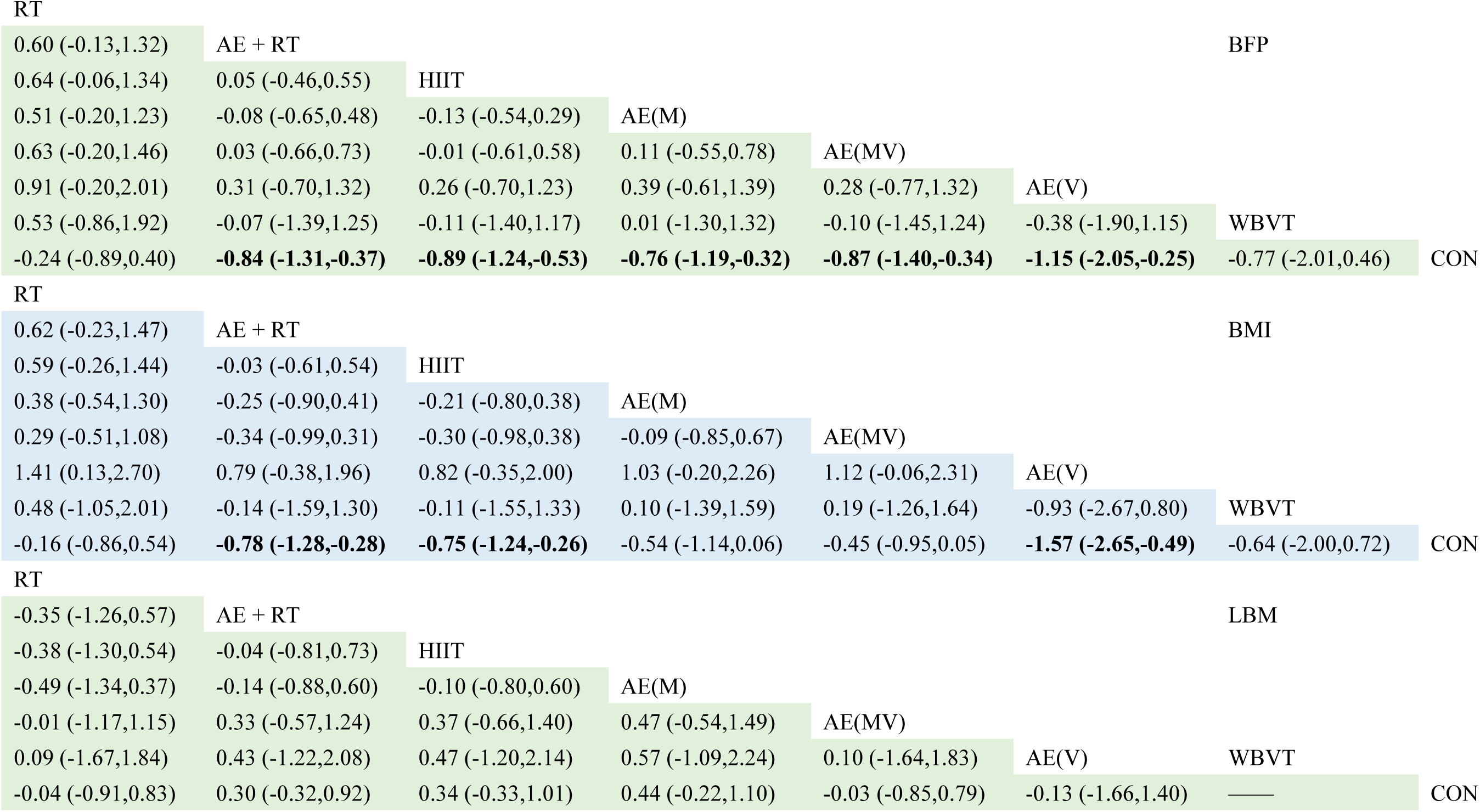
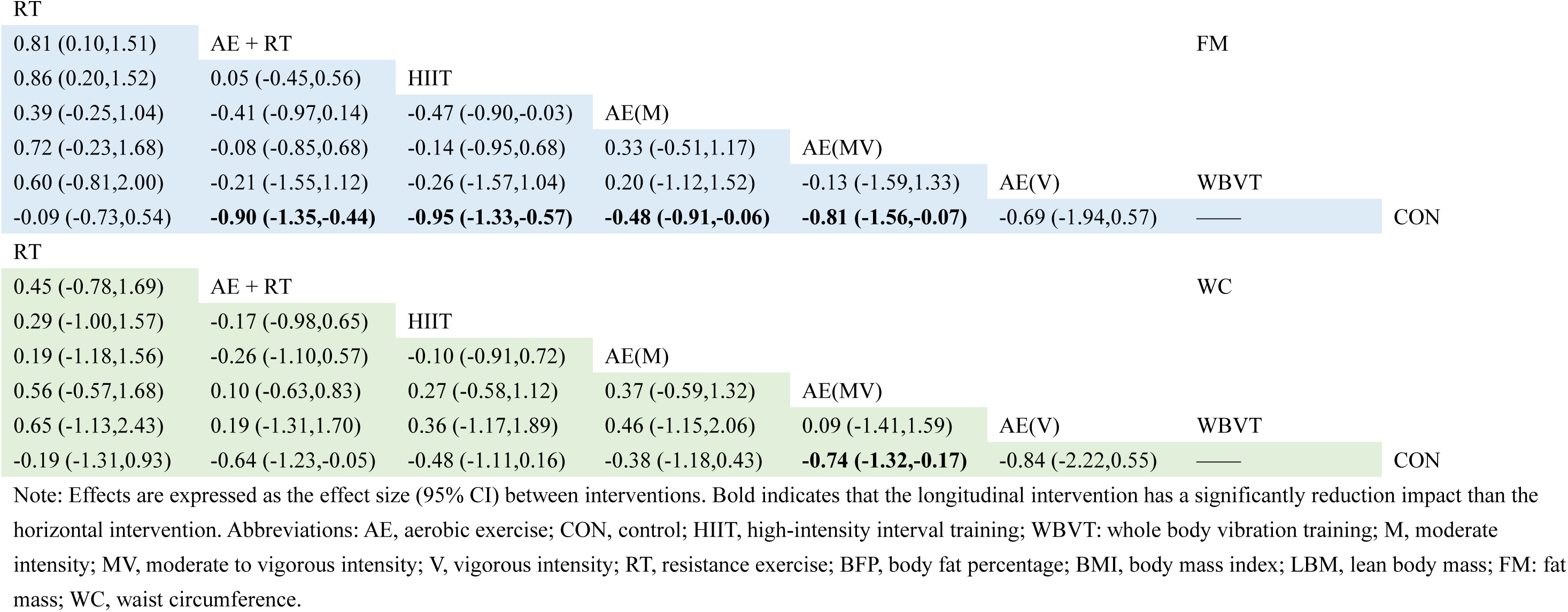
Network meta-analysis matrix of BFP and secondary outcomes.

|  |  |  |  |  |  |  |  |
| --- | --- | --- | --- | --- | --- | --- | --- |
| 0.60 (-0.13,1.32) | AE + RT |  |  |  |  |  | BFP |
| 0.64 (-0.06,1.34) | 0.05 (-0.46,0.55) | HIIT |  |  |  |  |  |
| 0.51 (-0.20,1.23) | -0.08 (-0.65,0.48) | -0.13 (-0.54,0.29) | AE(M) |  |  |  |  |
| 0.63 (-0.20,1.46) | 0.03 (-0.66,0.73) | -0.01 (-0.61,0.58) | 0.11 (-0.55,0.78) | AE(MV) |  |  |  |
| 0.91 (-0.20,2.01) | 0.31 (-0.70,1.32) | 0.26 (-0.70,1.23) | 0.39 (-0.61,1.39) | 0.28 (-0.77,1.32) | AE(V) |  |  |
| 0.53 (-0.86,1.92) | -0.07 (-1.39,1.25) | -0.11 (-1.40,1.17) | 0.01 (-1.30,1.32) | -0.10 (-1.45,1.24) | -0.38 (-1.90,1.15) | WBVT |  |
| -0.24 (-0.89,0.40) | **-0.84 (-1.31,-0.37)** | **-0.89 (-1.24,-0.53)** | **-0.76 (-1.19,-0.32)** | **-0.87 (-1.40,-0.34)** | **-1.15 (-2.05,-0.25)** | -0.77 (-2.01,0.46) | CON |
| RT |  |  |  |  |  |  |  |
| 0.62 (-0.23,1.47) | AE + RT |  |  |  |  |  | BMI |
| 0.59 (-0.26,1.44) | -0.03 (-0.61,0.54) | HIIT |  |  |  |  |  |
| 0.38 (-0.54,1.30) | -0.25 (-0.90,0.41) | -0.21 (-0.80,0.38) | AE(M) |  |  |  |  |
| 0.29 (-0.51,1.08) | -0.34 (-0.99,0.31) | -0.30 (-0.98,0.38) | -0.09 (-0.85,0.67) | AE(MV) |  |  |  |
| 1.41 (0.13,2.70) | 0.79 (-0.38,1.96) | 0.82 (-0.35,2.00) | 1.03 (-0.20,2.26) | 1.12 (-0.06,2.31) | AE(V) |  |  |
| 0.48 (-1.05,2.01) | -0.14 (-1.59,1.30) | -0.11 (-1.55,1.33) | 0.10 (-1.39,1.59) | 0.19 (-1.26,1.64) | -0.93 (-2.67,0.80) | WBVT |  |
| -0.16 (-0.86,0.54) | **-0.78 (-1.28,-0.28)** | **-0.75 (-1.24,-0.26)** | -0.54 (-1.14,0.06) | -0.45 (-0.95,0.05) | **-1.57 (-2.65,-0.49)** | -0.64 (-2.00,0.72) | CON |
| RT |  |  |  |  |  |  |  |
| -0.35 (-1.26,0.57) | AE + RT |  |  |  |  |  | LBM |
| -0.38 (-1.30,0.54) | -0.04 (-0.81,0.73) | HIIT |  |  |  |  |  |
| -0.49 (-1.34,0.37) | -0.14 (-0.88,0.60) | -0.10 (-0.80,0.60) | AE(M) |  |  |  |  |
| -0.01 (-1.17,1.15) | 0.33 (-0.57,1.24) | 0.37 (-0.66,1.40) | 0.47 (-0.54,1.49) | AE(MV) |  |  |  |
| 0.09 (-1.67,1.84) | 0.43 (-1.22,2.08) | 0.47 (-1.20,2.14) | 0.57 (-1.09,2.24) | 0.10 (-1.64,1.83) | AE(V) | WBVT |  |
| -0.04 (-0.91,0.83) | 0.30 (-0.32,0.92) | 0.34 (-0.33,1.01) | 0.44 (-0.22,1.10) | -0.03 (-0.85,0.79) | -0.13 (-1.66,1.40) | — | CON |
| RT |  |  |  |  |  |  |  |
| 0.81 (0.10,1.51) | AE + RT |  |  |  |  | FM |  |
| 0.86 (0.20,1.52) | 0.05 (-0.45,0.56) | HIIT |  |  |  |  |  |
| 0.39 (-0.25,1.04) | -0.41 (-0.97,0.14) | -0.47 (-0.90,-0.03) | AE(M) |  |  |  |  |
| 0.72 (-0.23,1.68) | -0.08 (-0.85,0.68) | -0.14 (-0.95,0.68) | 0.33 (-0.51,1.17) | AE(MV) |  |  |  |
| 0.60 (-0.81,2.00) | -0.21 (-1.55,1.12) | -0.26 (-1.57,1.04) | 0.20 (-1.12,1.52) | -0.13 (-1.59,1.33) | AE(V) | WBVT |  |
| -0.09 (-0.73,0.54) | **-0.90 (-1.35,-0.44)** | **-0.95 (-1.33,-0.57)** | **-0.48 (-0.91,-0.06)** | **-0.81 (-1.56,-0.07)** | -0.69 (-1.94,0.57) | — | CON |
| RT |  |  |  |  |  |  |  |
| 0.45 (-0.78,1.69) | AE + RT |  |  |  |  | WC |  |
| 0.29 (-1.00,1.57) | -0.17 (-0.98,0.65) | HIIT |  |  |  |  |  |
| 0.19 (-1.18,1.56) | -0.26 (-1.10,0.57) | -0.10 (-0.91,0.72) | AE(M) |  |  |  |  |
| 0.56 (-0.57,1.68) | 0.10 (-0.63,0.83) | 0.27 (-0.58,1.12) | 0.37 (-0.59,1.32) | AE(MV) |  |  |  |
| 0.65 (-1.13,2.43) | 0.19 (-1.31,1.70) | 0.36 (-1.17,1.89) | 0.46 (-1.15,2.06) | 0.09 (-1.41,1.59) | AE(V) | WBVT |  |
| -0.19 (-1.31,0.93) | -0.64 (-1.23,-0.05) | -0.48 (-1.11,0.16) | -0.38 (-1.18,0.43) | **-0.74 (-1.32,-0.17)** | -0.84 (-2.22,0.55) | — | CON |

Note: Effects are expressed as the effect size (95% CI) between interventions. Bold indicates that the longitudinal intervention has a significantly reduction impact than the horizontal intervention. Abbreviations: AE, aerobic exercise; CON, control; HIIT, high-intensity interval training; WBVT: whole body vibration training; M, moderate intensity; MV, moderate to vigorous intensity; V, vigorous intensity; RT, resistance exercise; BFP, body fat percentage; BMI, body mass index; LBM, lean body mass; FM: fat mass; WC, waist circumference.

**Table 3.** Ranking of exercise interventions in order of effectiveness.

| Body fat percentage |  | Body mass index |  | Lean body mass |  | Waist circumference |  | Fat mass |  |
| --- | --- | --- | --- | --- | --- | --- | --- | --- | --- |
| Treatment | SUCRA | Treatment | SUCRA | Treatment | SUCRA | Treatment | SUCRA | Treatment | SUCRA |
| CON | 4.9 | CON | 8.4 | CON | 65.4 | CON | 9.3 | CON | 12.7 |
| RT | 18.3 | RT | 22.4 | RT | 65.5 | RT | 17.1 | RT | 32.7 |
| AE+RT | 60.7 | AE+RT | 68.2 | AE+RT | 35.3 | AE+RT | 75.8 | AE+RT | 65.6 |
| HIIT | 66.3 | HIIT | 65.5 | HIIT | 32.9 | HIIT | 80.9 | HIIT | 51.7 |
| AE(M) | 52.2 | AE(M) | 47.1 | AE(M) | 23.4 | AE(M) | 42.3 | AE(M) | 43.7 |
| AE(MV) | 63.7 | AE(MV) | 41.6 | AE(MV) | 63.9 | AE(MV) | 67.6 | AE(MV) | 73.4 |
| AE(V) | 79.7 | AE(V) | 93.9 | AE(V) | 63.6 | AE(V) | 57.1 | AE(V) | 70.2 |
| WBVT | 54.2 | WBVT | 52.9 | WBVT | — | WBVT | — | WBVT | — |

### Pooled estimates of the secondary outcome

We also performed a NMA on secondary outcomes, including LBM, BMI, WC, and FM, to determine whether the effects of different exercise modalities on these outcomes were consistent with their effects on BFP.

A network meta-analysis was conducted to compare the effects of various exercise modalities on body composition outcomes. For BMI, AE + RT (SMD = -0.78, 95% CI: -1.28 to -0.28, p < 0.001), HIIT (SMD = -0.75, 95% CI: -1.24 to -0.26, p < 0.001), and AE(V) (SMD = -1.57, 95% CI: -2.65 to -0.49, p < 0.001) produced significant reductions relative to control. Based on SUCRA values, AE(V) ranked as the most effective intervention (SUCRA = 93.9), followed by AE + RT (SUCRA = 68.2), HIIT (SUCRA = 65.5), WBVT (SUCRA = 52.9), and AE(M) (SUCRA = 47.1), whereas RT (SUCRA = 22.4) was least effective. AE(M), AE(MV), RT, and WBVT did not differ significantly from control or from each other. For FM, AE + RT, HIIT, AE(M), and AE(MV) all elicited significant decreases, with AE(MV) achieving the highest SUCRA value (SUCRA = 73.4), followed by AE(V) (SUCRA = 70.2), AE + RT (SUCRA = 65.6), HIIT (SUCRA = 51.7), and AE(M) (SUCRA = 43.7), while RT (SUCRA = 32.7) remained the lowest. AE(V) and RT showed no significant differences compared with control or other exercise types. In terms of LBM, none of the interventions demonstrated significant improvements, although SUCRA rankings suggested a potential hierarchy led by RT (SUCRA = 65.5), followed by AE + RT (SUCRA = 35.3), HIIT (SUCRA = 32.9), and AE(M) (SUCRA = 23.4). Regarding WC, AE(MV) was the only modality to significantly reduce values compared with control (SMD = -0.74, 95% CI: -1.32 to -0.17, p < 0.05), yet SUCRA rankings indicated that HIIT (SUCRA = 80.9) had the greatest probability of being most effective, followed by AE + RT (SUCRA = 75.8), AE(MV) (SUCRA = 67.6), AE(V) (SUCRA = 57.1), AE(M) (SUCRA = 42.3), and RT (SUCRA = 17.1).

### Subgroup NMA of primary outcome

A subgroup analysis of BFP was performed based on date of publication outlined in Table 1 to explore whether these covariates influenced the study outcomes (Appendix S14). The corresponding subgroup network diagram is presented in Appendix S12.

Analysis of subgroup-specific heterogeneity by date of publication demonstrated good consistency across all closed loops for BFP. Node-splitting analysis further indicated that the direct and indirect comparison results were consistent across all subgroups.

In the recent studies subgroup, the network meta-analysis of BFP revealed that, compared with the control group, AE + RT (SMD = -1.18, 95% CI [-2.26, -0.10], p < 0.05), HIIT (SMD = -1.01, 95% CI [-1.65, -0.36], p < 0.01), significantly reduced BFP.

SUCRA rankings suggested that AE(V) (SUCRA = 82.1) was most likely the optimal intervention, whereas RT (SUCRA = 24.1) was considered less effective.

In the earlier studies subgroup, the network meta-analysis of BFP showed that, compared with the control group, AE + RT (SMD = -0.79, 95% CI [-1.35, -0.22], p < 0.01), AE(M) (SMD = -0.89, 95% CI [-1.59, -0.19], p < 0.05), HIIT (SMD = -0.79, 95% CI [-1.31, 0.27], p < 0.01) and AE(MV) (SMD = -0.86, 95% CI [-1.47, -0.26], p < 0.01) all contributed to reductions in BFP. SUCRA rankings indicated that AE(M) (SUCRA = 71.5) was most likely the optimal intervention, whereas RT (SUCRA = 27.6) was the least effective intervention.

### CINeMA assessment

Appendix S13 presents the CINeMA assessments for each comparison, along with the SUCRA rankings for primary and secondary outcomes. Overall, the certainty of evidence for most comparisons of BFP, LBM, FM, BMI, and WC ranged from moderate to low.

## DISCUSSION

Building upon prior research, this study provides a critical refinement and expansion of the evidence base by specifically investigating the effects of exercise in women with overweight and obesity and by incorporating a broader spectrum of exercise modalities. This systematic review and network meta-analysis included 44 randomized controlled trials involving 2384 female participants. The analysis revealed a distinct, outcome-specific hierarchy of effectiveness: AE(V) was the most effective for reducing BMI, AE(MV) for decreasing FM and WC. While multiple modalities reduced BFP, RT showed the weakest effect. Notably, our subgroup analysis uncovered a previously unreported finding that the optimal exercise for reducing BFP has varied over time. These results underscore the necessity of sex-specific analysis and provide clinicians with a nuanced, evidence-based framework for prescribing personalized exercise to women.

### Effect of exercise on primary outcomes

Our analysis unequivocally demonstrates that structured exercise is a potent intervention for reducing BFP. The consensus across multiple modalities—including AE + RT, HIIT, and AE (moderate, moderate-to-vigorous, and vigorous) strengthens the foundational role of physical activity in managing body fat.

Previous meta-analyses have consistently demonstrated that AE is effective in improving body composition among individuals with overweight and obesity, particularly through reductions in BFP.^40^ However, by synthesizing a broader and more recent body of evidence in the present study, we observed that the efficacy of AE is significantly influenced by exercise intensity. Specifically, Our network meta-analysis revealed that AE(V) demonstrated a significant effect in reducing body fat percentage and achieved the highest SUCRA score, while both AE(M) and AE(MV) showed lower effect sizes and SUCRA values compared with AE(V), suggesting that it is most likely the optimal intervention among all interventions. This aligns with recent findings suggesting a dose and response relationship between exercise intensity and adiposity outcomes. For instance, Khodadadi et al. (2023) reported that high-intensity interventions yielded greater reductions in BFP compared to moderate or low-intensity protocols.^43^ These findings underscore the importance of tailoring exercise prescriptions not only by modality but also by intensity, to maximize improvements in body composition.

Contrary to earlier meta-analytic findings suggesting that RT may contribute to reductions in BFP,^45^ our results did not identify a statistically significant effect of RT on BFP, and RT showed the lowest SUCRA value among all interventions. This divergence may stem from considerable heterogeneity in RT protocols across studies, including variations in exercise modality, total training volume, repetition schemes, and inter-set work-to-rest ratios. Such variability complicates quantitative synthesis and may obscure potential effects. Compared to AE, RT interventions exhibit greater inconsistency in design and implementation, which likely contributes to the mixed outcomes observed. For instance, Liu et al. (2022) noted that different RT formats (e.g., circuit and traditional) yielded divergent effects on body composition in overweight populations.^16^ Therefore, we do not recommend RT alone as a primary strategy for improving BFP, although it may offer complementary benefits when combined with other modalities.

The present findings reinforce existing evidence regarding the efficacy of AE+RT in improving BFP among individuals with overweight and obesity.^42^ Our network meta-analysis demonstrated that AE+RT significantly reduced BFP in women and ranked as the fourth most effective intervention according to SUCRA probability estimates. This is consistent with prior meta-analyses indicating that multimodal exercise regimens yield superior outcomes in body composition compared to single-modality interventions.^46,47^ RT, while primarily recognized for its role in enhancing muscular strength and preserving lean mass, may exert synergistic effects when combined with aerobic exercise, contributing to reductions in adiposity and improvements in muscle and bone density.^48,17^ These findings support the integration of AE and RT in exercise prescriptions targeting comprehensive body composition improvements.

HIIT could significantly improve BFP with a moderate effect in individuals with overweight and obesity compared with controls. The SUCRA probability ranking showed that HIIT ranks only second to AE(V), suggesting that HIIT is a time-efficient strategy for decreasing BFP.We found that AE(V) and HIIT were the most effective types of exercise to reduce BFP, and the common feature of both exercises was high-intensity stimulation, and a shared characteristic between them was the application of high-intensity stimuli, which may underpin their superior efficacy in adiposity reduction.^43,44^

The findings of this study regarding WBVT do align with previous research, as WBVT did not significantly improve BFP.^45^ This discrepancy may be attributed to the limited number of included studies and the stricter inclusion criteria applied in the present analysis, which only considered interventions lasting eight weeks or longer, whereas most existing studies implemented protocols of approximately six weeks.

### Secondary outcomes

The analysis of secondary outcomes reveals that the best exercise intervention is profoundly outcome-dependent, highlighting the multifaceted nature of body composition changes.

For BMI and FM, the results partially mirrored those of BFP. AE(V) was again ranked highest for BMI reduction, exhibiting a very large effect size, while AE(MV) was ranked highest for FM reduction which is similar to previous research.^18^ This consistency across fat-related metrics (BFP, BMI, FM) reinforces the robustness of high-intensity exercises for fat loss. The strong performance of AE + RT across BFP, BMI, and FM further solidifies its position as a comprehensive and highly effective strategy.

The findings for LBM and WC, however, presented divergent narratives. No intervention significantly improved LBM over control, which is surprising for RT. However, the SUCRA ranking suggested RT had the highest probability of being optimal for LBM, indicating a signal of effect that may have been underpowered in the direct and indirect comparisons. This underscores that while RT’s effect on absolute LBM might be modest in these studies, it may be crucial in preserving LBM during weight loss, a key advantage over purely aerobic regimens.^12,49^ For WC, a marker of abdominal adiposity, AE(MV) was the only intervention with a significant effect, yet AE + RT had the highest SUCRA probability. This discrepancy between statistical significance and SUCRA values for WC warrants caution in interpretation but points towards combined training as a promising approach for targeting central obesity.

### Subgroup NMA of primary outcome

The subgroup analysis based on publication date offers a intriguing perspective on the potential evolution of exercise science. The consistency and invariance tests confirmed the robustness of the network within subgroups, strengthening the validity of subsequent comparisons.

In recent studies, the field appears to have narrowed its focus, with only AE + RT and HIIT demonstrating statistically significant effects on BFP compared to control. This may reflect a contemporary research trend favoring these time-efficient or multimodal hybrid training models. Despite this, AE(V) retained the highest SUCRA ranking, suggesting it remains a potent, though perhaps less frequently studied, intervention in modern literature.

Conversely, in earlier studies, a broader range of interventions—including AE(M), AE(MV), AE + RT, and HIIT—showed significant benefits. Notably, AE(M) was ranked as the optimal intervention in this subgroup. This shift in the most effective modality over time could be attributed to several factors, including changes in study population characteristics, refinements in the exercise protocols, or simply a reflection of the specific hypotheses tested in the literature available for each era. This temporal heterogeneity underscores the importance of interpreting NMA findings within the context of the evolving scientific landscape.

### Limitations

This study has several unavoidable limitations. First, to ensure the reliability of the meta-analysis, two studies with high heterogeneity were excluded during the preliminary assessment, which may have slightly reduced the overall sample size and affected the representativeness of the findings. Second, due to variations in how exercise interventions were described across studies, short-term sprint interval training (SIT) was classified as HIIT. This classification may differ from the definitions used in the original studies, thereby introducing certain limitations in interpreting the results. Finally, the number of studies investigating WBVT was relatively small, with limited sample sizes, resulting in insufficient evidence to comprehensively evaluate its effects and potentially affecting the conclusiveness of judgments regarding this intervention.

## CONCLUSION

This network meta-analysis demonstrates that structured exercise interventions are superior to control for improving body composition. For the primary outcome, AE + RT, HIIT, RT and AE across various intensities all significantly reduce BFP compared to control. AE(V) tends to be the most effective intervention. This pattern is consistent for other adiposity markers, with AE(V) being likely optimal for reducing body mass index and AE(MV) being likely optimal for reducing FM and WC. In contrast, although no intervention significantly improves LBM, RT shows a potential advantage. The subgroup analysis based on publication date reinforce these findings, confirming the consistent effectiveness of exercise and the inferiority of RT alone across different time periods. While the top-ranked exercise type for reducing BFP varies between subgroups, the overall conclusion remains that AE are central for effective fat reduction. Exercise prescriptions for improving body composition should therefore prioritize these modalities over RT alone.

## Contributors

Shiwei Song,Yincheng Wei and Haoze Zhang are joint first authors. All authors carried out the screenings and reviews, and the analysis of the articles. Shiwei Song drafted the manuscript, and Andrew Soundy, Yincheng Wei and Haoze Zhang revised the manuscript. All authors read and approved the final manuscript.

## Ethics approval

This study does not require ethics approval as it is a review based on published studies.

## Data availability statement

All data relevant to the study are included in the article or uploaded as supplemental information.

## Data Availability

All data produced in the present study are available upon reasonable request to the authors

