## Appendix for "Effects of Different Exercise Interventions on Body Composition in Women with Overweight and Obesity: A Systematic Review and Network Meta-analysis"

**Title**：

| Supplemental materials | Page |
| --- | --- |
| **Appendix 1.** Completed PRISMA-NMA checklist. | 3-7 |
| **Appendix 2.** Search strategy of PubMed, Embase, The Cochrane library, Web of Science, and Scopus. | 8-10 |
| **Appendix 3.** Characteristics of the included studies. | 11-16 |
| **Appendix 4**. List of included studies. | 17-19 |
| **Appendix 5.** Risk of bias assessment. | 20-22 |
| **Appendix 6.** Contributions of direct and indirect comparisons to NMA and the  number of studies of each direct comparison of body fat percentage, BMI, lean body mass, waist circumference, and fat mass. | 23-25 |
| **Appendix 7.** Inconsistency of body fat percentage and secondary outcomes  tested by loop-specific heterogeneity estimates, inconsistency model, and node splitting analysis. | 26-29 |
| **Appendix 8.** Forest plots of eligible comparisons of body fat percentage, BMI, lean body mass, waist circumference, and fat mass. | 30-32 |
| **Appendix 9.** The funnel plot graphics of body fat percentage, BMI, lean body mass, waist circumference, and fat mass in NMA. | 33 |
| **Appendix 10.** Area under the curve for cumulative ranking probability of each intervention on body fat percentage, body mass index, lean body mass, fat mass, waist circumference. | 34-36 |
| **Appendix 11.** Forest plot of individual studies. | 37-40 |
| **Appendix 12.** Network plot comparing the effects of different exercise types on body fat percentage between recent (≤5 years) and earlier studies (>5 years). | 41 |
| **Appendix 13.** CINeMA for the primary and secondary outcomes. | 41-43 |
| **Appendix 14.** Results of subgroup analysis of body fat percentage of studies utilizing imaging techniques. | 44-45 |

**Appendix 1.** PRISMA NMA Checklist of Items to Include When Reporting A Systematic Review Involving a Network Meta-analysis

| **Section/Topic** | **Item #** | **Checklist Item** | **Reported on Page #** |
| --- | --- | --- | --- |
| **TITLE** |  |  |  |
| Title | 1 | Identify the report as a systematic review *incorporating a network meta-analysis (or related form of meta-analysis).* | 1 |
| **ABSTRACT** |  |  |  |
| Structured summary | 2 | Provide a structured summary including, as applicable:  **Background:** main objectives  **Methods:** data sources; study eligibility criteria, participants, and interventions; study appraisal; and *synthesis methods, such as network meta-analysis.*  **Results:** number of studies and participants identified; summary estimates with corresponding confidence/credible intervals; *treatment rankings may also be discussed. Authors may choose to summarize pairwise comparisons against a chosen treatment included in their analyses for brevity.*  **Discussion/Conclusions:** limitations; conclusions and implications of findings.  **Other:** primary source of funding; systematic review registration number with registry name. | 1 |
| **INTRODUCTION** |  |  |  |
| Rationale | 3 | Describe the rationale for the review in the context of what is already known*, including mention of why a network meta-analysis has been conducted.* | 1-2 |
| Objectives | 4 | Provide an explicit statement of questions being addressed, with reference to participants, interventions, comparisons, outcomes, and study design (PICOS). | 2-3 |
| **METHODS** |  |  |  |
| Protocol and registration | 5 | Indicate whether a review protocol exists and if and where it can be accessed (e.g., Web address); and, if available, provide registration information, including registration number. |  |
| Eligibility criteria | 6 | Specify study characteristics (e.g., PICOS, length of follow-up) and report characteristics (e.g., years considered, language, publication status) used as criteria for eligibility, giving rationale. *Clearly describe eligible treatments included in the treatment network, and note whether any have been clustered or merged into the same node (with justification).* | 3 |
| Information sources | 7 | Describe all information sources (e.g., databases with dates of coverage, contact with study authors to identify additional studies) in the search and date last searched. | 2-3 |
| Search | 8 | Present full electronic search strategy for at least one database, including any limits used, such that it could be repeated. | Appendix 2 |
| Study selection | 9 | State the process for selecting studies (i.e., screening, eligibility, included in systematic review, and, if applicable, included in the meta-analysis). | 2-3 |
| Data collection process | 10 | Describe method of data extraction from reports (e.g., piloted forms, independently, in duplicate) and any processes for obtaining and confirming data from investigators. | 4-5 |
| Data items | 11 | List and define all variables for which data were sought (e.g., PICOS, funding sources) and any assumptions and simplifications made. | Appendix 3 |
| **Geometry of the network** | **S1** | Describe methods used to explore the geometry of the treatment network under study and potential biases related to it. This should include how the evidence base has been graphically summarized for presentation, and what characteristics were compiled and used to describe the evidence base to readers. | 5-6 |
| Risk of bias within individual studies | 12 | Describe methods used for assessing risk of bias of individual studies (including specification of whether this was done at the study or outcome level), and how this information is to be used in any data synthesis. | 5  Appendix 5 |
| Summary measures | 13 | State the principal summary measures (e.g., risk ratio, difference in means). *Also describe the use of additional summary measures assessed, such as treatment rankings and surface under the cumulative ranking curve (SUCRA) values, as well as modified approaches used to present summary findings from meta-analyses.* | 5-6 |
| Planned methods of analysis | 14 | Describe the methods of handling data and combining results of studies for each network meta-analysis. This should include, but not be limited to:   - *Handling of multi-arm trials;* - *Selection of variance structure;* - *Selection of prior distributions in Bayesian analyses; and* - *Assessment of model fit.* | 5-6 |
| **Assessment of Inconsistency** | **S2** | Describe the statistical methods used to evaluate the agreement of direct and indirect evidence in the treatment network(s) studied. Describe efforts taken to address its presence when found. | 5-6  Appendix 7 |
| Risk of bias across studies | 15 | Specify any assessment of risk of bias that may affect the cumulative evidence (e.g., publication bias, selective reporting within studies). | 5-6 |
| Additional analyses | 16 | Describe methods of additional analyses if done, indicating which were pre-specified. This may include, but not be limited to, the following:   - Sensitivity or subgroup analyses; - Meta-regression analyses; - *Alternative formulations of the treatment network; and* - *Use of alternative prior distributions for Bayesian analyses (if applicable).* | 5-6 |
| **RESULTS†** |  |  |  |
| Study selection | 17 | Give numbers of studies screened, assessed for eligibility, and included in the review, with reasons for exclusions at each stage, ideally with a flow diagram. | 6 |
| **Presentation of network structure** | **S3** | Provide a network graph of the included studies to enable visualization of the geometry of the treatment network. | Figure 2 |
| **Summary of network geometry** | **S4** | Provide a brief overview of characteristics of the treatment network. This may include commentary on the abundance of trials and randomized patients for the different interventions and pairwise comparisons in the network, gaps of evidence in the treatment network, and potential biases reflected by the network structure. | 8 |
| Study characteristics | 18 | For each study, present characteristics for which data were extracted (e.g., study size, PICOS, follow-up period) and provide the citations. | 6-7 |
| Risk of bias within studies | 19 | Present data on risk of bias of each study and, if available, any outcome level assessment. | 7-8 |
| Results of individual studies | 20 | For all outcomes considered (benefits or harms), present, for each study: 1) simple summary data for each intervention group, and 2) effect estimates and confidence intervals. *Modified approaches may be needed to deal with information from larger networks.* | Appendix 11 |
| Synthesis of results | 21 | Present results of each meta-analysis done, including confidence/credible intervals. *In larger networks, authors may focus on comparisons versus a particular comparator (e.g. placebo or standard care), with full findings presented in an appendix. League tables and forest plots may be considered to summarize pairwise comparisons.* If additional summary measures were explored (such as treatment rankings), these should also be presented. | 9-11 |
| **Exploration for inconsistency** | **S5** | Describe results from investigations of inconsistency. This may include such information as measures of model fit to compare consistency and inconsistency models, *P* values from statistical tests, or summary of inconsistency estimates from different parts of the treatment network. | Appendix 7 |
| Risk of bias across studies | 22 | Present results of any assessment of risk of bias across studies for the evidence base being studied. | 13  Appendix 13 |
| Results of additional analyses | 23 | Give results of additional analyses, if done (e.g., sensitivity or subgroup analyses, meta-regression analyses*, alternative network geometries studied, alternative choice of prior distributions for Bayesian analyses,* and so forth). | 12-13 |
| **DISCUSSION** |  |  |  |
| Summary of evidence | 24 | Summarize the main findings, including the strength of evidence for each main outcome; consider their relevance to key groups (e.g., healthcare providers, users, and policy-makers). | 12-13 |
| Limitations | 25 | Discuss limitations at study and outcome level (e.g., risk of bias), and at review level (e.g., incomplete retrieval of identified research, reporting bias). *Comment on the validity of the assumptions, such as transitivity and consistency. Comment on any concerns regarding network geometry (e.g., avoidance of certain comparisons).* | 16 |
| Conclusions | 26 | Provide a general interpretation of the results in the context of other evidence, and implications for future research. | 17 |
| **FUNDING** |  |  |  |
| Funding | 27 | Describe sources of funding for the systematic review and other support (e.g., supply of data); role of funders for the systematic review. This should also include information regarding whether funding has been received from manufacturers of treatments in the network and/or whether some of the authors are content experts with professional conflicts of interest that could affect use of treatments in the network. | Not applicable |

**Appendix 2.** Search strategy of PubMed, Embase, Cochrane, Web of Science, and Scopus.

| **Database** | **Search terms** | | | | |
| --- | --- | --- | --- | --- | --- |
|  | **Exercise** | **Obesity** | **Female** | **Body composition** | **Randomized controlled trial** |
| Pubmed [Title/Abstract] | Exercise [MeSH Terms] OR exercises OR exercise, physical OR exercises, physical OR physical exercise OR physical exercises OR exercise, aerobic OR aerobic exercise OR aerobic exercises OR exercises, aerobic OR exercise training OR exercise trainings OR training, exercise OR trainings, exercise OR physical activity OR activities, physical OR physical activities OR activity, physical OR Tai Chi OR resistance training OR strength training OR combined training OR Qigong OR Whole-Body Vibration Training OR Baduanjin | Obesity [MeSH Terms] OR obese OR overweight | Female [MeSH Terms] OR females OR women [MeSH Terms] OR girls OR girl OR woman OR women groups OR women's group | Body composition [MeSH Terms] OR body component OR lean body mass OR body weight OR body adiposity index OR body fat OR body fat percentage OR Body Compositions | Randomized controlled trial OR randomized OR placebo |
| Embase [Title/Abstract] | Exercise [MeSH Terms] OR exercises OR exercise, physical OR exercises, physical OR physical exercise OR physical exercises OR exercise, aerobic OR aerobic exercise OR aerobic exercises OR exercises, aerobic OR exercise training OR exercise trainings OR training, exercise OR trainings, exercise OR physical activity OR activities, physical OR physical activities OR activity, physical OR Tai Chi OR resistance training OR strength training OR combined training OR Qigong OR Whole-Body Vibration Training OR Baduanjin | Obesity [MeSH Terms] OR obese OR overweight | Female [MeSH Terms] OR females OR women [MeSH Terms] OR girls OR girl OR woman OR women groups OR women's group | Body composition [MeSH Terms] OR body component OR lean body mass OR body weight OR body adiposity index OR body fat OR body fat percentage OR Body Compositions | Randomized controlled trial OR randomized OR placebo |
| Cochrane [Title/Abstract/keywords] | Exercise [MeSH Terms] OR exercises OR exercise, physical OR exercises, physical OR physical exercise OR physical exercises OR exercise, aerobic OR aerobic exercise OR aerobic exercises OR exercises, aerobic OR exercise training OR exercise trainings OR training, exercise OR trainings, exercise OR physical activity OR activities, physical OR physical activities OR activity, physical OR Tai Chi OR resistance training OR strength training OR combined training OR Qigong OR Whole-Body Vibration Training OR Baduanjin | Obesity [MeSH Terms] OR obese OR overweight | Female [MeSH Terms] OR females OR women [MeSH Terms] OR girls OR girl OR woman OR women groups OR women's group | Body composition [MeSH Terms] OR body component OR lean body mass OR body weight OR body adiposity index OR body fat OR body fat percentage OR Body Compositions | —— |
| Web of Science | Exercise OR exercises OR exercise, physical OR exercises, physical OR physical exercise OR physical exercises OR exercise, aerobic OR aerobic exercise OR aerobic exercises OR exercises, aerobic OR exercise training OR exercise trainings OR training, exercise OR trainings, exercise OR physical activity OR activities, physical OR physical activities OR activity, physical OR Tai Chi OR resistance training OR strength training OR combined training OR Qigong OR Whole-Body Vibration Training OR Baduanjin | Obesity OR obese  OR overweight | Female OR females OR women OR girls OR girl OR woman OR women groups OR women's group | Body composition OR body component OR lean body mass OR body weight OR body adiposity index OR body fat OR body fat percentage OR Body Compositions | Randomized controlled trial OR randomized OR placebo |
| Scopus [Title/Abstract/keywords] | Exercise OR exercises OR exercise, physical OR exercises, physical OR physical exercise OR physical exercises OR exercise, aerobic OR aerobic exercise OR aerobic exercises OR exercises, aerobic OR exercise training OR exercise trainings OR training, exercise OR trainings, exercise OR physical activity OR activities, physical OR physical activities OR activity, physical OR Tai Chi OR resistance training OR strength training OR combined training OR Qigong OR Whole-Body Vibration Training OR Baduanjin | Obesity OR obese  OR overweight | Female OR females OR women OR girls OR girl OR woman OR women groups OR women's group | Body composition OR body component OR lean body mass OR body weight OR body adiposity index OR body fat OR body fat percentage OR Body Compositions | Randomized controlled trial OR randomized OR placebo |
| CINAHL | Exercises OR AB Exercise, Physical OR AB Exercises, Physical OR AB Physical Exercise OR AB Physical Exercises OR AB Exercise, Aerobic OR AB Aerobic Exercise OR AB Aerobic Exercises OR AB Exercises, Aerobic OR AB Exercise Training OR AB Exercise Trainings OR AB Training, Exercise OR AB Trainings, Exercise OR AB Physical Activity OR AB Activities, Physical OR AB Activity, Physical OR AB Physical Activities OR AB Tai Chi OR AB resistance training OR AB strength training OR AB combined training OR AB Qigong OR AB Whole-Body Vibration Training OR AB Baduanjin | obese OR AB overweight OR AB Obesity OR AB adipose tissue hyperplasia OR AB adipositas OR AB adiposity OR AB alimentary obesity OR AB body weight, excess OR AB corpulency OR AB fat overload OR AB syndrome nutritional OR AB obesity OR AB obesitas OR AB overweight | females OR AB woman OR AB women OR AB female | body component OR AB lean body mass OR AB body weight OR AB body adiposity index OR AB body fat OR AB body fat percentage OR AB body composition | randomized controlled trial OR AB randomized OR AB placebo |
| SportsDicus | Exercises OR AB Exercise, Physical OR AB Exercises, Physical OR AB Physical Exercise OR AB Physical Exercises OR AB Exercise, Aerobic OR AB Aerobic Exercise OR AB Aerobic Exercises OR AB Exercises, Aerobic OR AB Exercise Training OR AB Exercise Trainings OR AB Training, Exercise OR AB Trainings, Exercise OR AB Physical Activity OR AB Activities, Physical OR AB Activity, Physical OR AB Physical Activities OR AB Tai Chi OR AB resistance training OR AB strength training OR AB combined training OR AB Qigong OR AB Whole-Body Vibration Training OR AB Baduanjin | obese OR AB overweight OR AB Obesity OR AB adipose tissue hyperplasia OR AB adipositas OR AB adiposity OR AB alimentary obesity OR AB body weight, excess OR AB corpulency OR AB fat overload OR AB syndrome nutritional OR AB obesity OR AB obesitas OR AB overweigh | females OR AB woman OR AB women OR AB female | body component OR AB lean body mass OR AB body weight OR AB body adiposity index OR AB body fat OR AB body fat percentage OR AB body composition | randomized controlled trial OR AB randomized OR AB placebo |

**Appendix 3.** Characteristics of the included studies

| **Study** | **Country** | **Characteristics of subject** | | | | | | **Interventions information** | | | | **Outcome** | | |
| --- | --- | --- | --- | --- | --- | --- | --- | --- | --- | --- | --- | --- | --- | --- |
|  |  | **Number** | **Age (mean[SD])** | **BMI (mean[SD])** | **fat% (mean[SD])** | **Fat Mass(mean[SD])** | **Lean Body Mass(mean[SD])** | **Type of exercise** | **Exercise Regimen** | **Period and frequency (week × [times/week])** | **supervised or nonsupervised** | **Primary outcome measure** | **Measurement method and unit** | **Secondary outcomes** |
| Alves et al. 2009^1^ | Brazil | 71 | 39.40 ± 11.00 | 29.70 ± 3.13 |  |  |  | AE（M) | Walking, 40–60% HRR, 40 min/session | 24*3 | supervised |  |  | ④ |
|  |  | 75 | 37.00 ± 10.60 | 29.70 ± 3.14 |  |  |  | CON | Maintain a normal life |  |  |  |  |  |
| Gappmaier et al. 2006^2^ | USA | 13 | 34.80 ± 6.90 |  | 35.10 ± 5.30 |  |  | AE（M) | Walking, 70% MHP, 40 min/session | 13*4 | supervised | fat% | HW，% | ① |
|  |  | 12 | 35.80 ± 9.90 |  | 36.90 ± 6.20 |  |  | AE（M) | Water walking, 70% MHP, 40 min/session | 13*4 | supervised |  |  |  |
|  |  | 13 | 33.60 ± 4.70 |  | 35.0 ± 5.80 |  |  | AE（M) | Water walking, 70% MHP, 40 min/session | 13*4 | supervised |  |  |  |
| Lestari et al. 2025^3^ | Indonesia | 10 | 23.30 ± 1.34 | 30.43 ± 1.39 | 34.70 ± 3.11 | 26.45 ± 5.21 | 35.43 ± 5.96 | AE + RT | Aerobic exercise + resistance training,60–70% HRmax; 60–70% 1RM, 30–40 min/session | 8*4 | supervised | fat% | TANITA，% | ①②④ |
|  |  | 10 | 23.20 ± 1.48 | 30.54 ± 2.26 | 34.79 ± 1.91 | 25.15 ± 3.82 | 33.89 ± 3.12 | CON | Maintain a normal life |  |  |  |  |  |
| Ajjimaporn et al. 2023^4^ | Thailand | 12 | 31.00 ± 2.00 | 27.90 ± 2.50 |  |  |  | AE + RT | High–intensity circuit training, 44–63% HRR | 8*3 | supervised | fat% | BIA，% | ② |
|  |  | 12 | 32.00 ± 3.00 | 26.70 ± 3.00 |  |  |  | CON | Maintain a normal life |  |  |  |  |  |
| Zhang et al. 2017^5^ | China | 15 | 21.50 ± 1.70 |  | 38.10 ± 2.30 | 25.70 ± 3.30 |  | HIIT | Cycling (interval), 90% VO2max, 30–40 min/session | 1–4week*3  5–12week*4 | supervised | fat% | DEXA，% | ② |
|  |  | 15 | 20.90 ± 1.40 |  | 38.00 ± 2.10 | 26.10 ± 3.70 |  | AE（M) | Cycling, 60% VO_2_max | 1–4week*3  5–12week*4 | supervised |  |  |  |
|  |  | 13 | 20.80 ± 1.10 |  | 40.90 ± 2.90 | 27.80 ± 5.40 |  | CON | Maintain a normal life |  |  |  |  |  |
| Tong et al. 2018^6^ | China | 16 | 21.30 ± 1.00 |  | 38.40 ± 2.30 | 25.70 ± 3.50 |  | HIIT | Cycling (sprint), all–out | 1–4week*3  5–12week*4 | supervised | fat% | DEXA，% | ② |
|  |  | 16 | 21.30 ± 10.00 |  | 38.20 ± 2.40 | 26.60 ± 6.10 |  | HIIT | Cycling (interval), 90% VO_2_max | 1–4week*3  5–12week*4 | supervised |  |  |  |
|  |  | 14 | 20.70 ± 1.50 |  | 40.50 ± 2.60 | 27.90 ± 5.10 |  | CON | Maintain a normal life |  |  |  |  |  |
| Lee et al. 2023^7^ | Korea | 8 | 42.25 ± 2.17 | 25.50 ± 4.10 |  |  |  | AE + RT | Treadmill + resistance (TRX), 50% VO_2_max; 60–70% HRmax, 45–48 min/session | 8*5 | supervised | fat% | ST+  Siri/Jackson，% |  |
|  |  | 8 | 43.00 ± 2.41 | 25.80 ± 4.20 |  |  |  | AE + RT | Treadmill + resistance (TRX), 80% VO_2_max; 60–70% HRmax, 30–33 min/session | 8*5 | supervised |  |  |  |
| Manthou et al. 2015^8^ | UK | 16 | 32.10 ± 1.80 | 29.70 ± 1.10 | 40.40 ± 1.30 | 33.80 ± 2.80 | 47.90 ± 1.50 | AE（MV) | Cycling (indoor), 90–95% LT, 75 min/session | 8*2 | supervised | fat% | TANITA，% | ①②③④ |
|  |  | 18 | 31.30 ± 2.10 | 29.00 ± 1.00 | 46.40 ± 1.10 | 29.50 ± 1.80 | 46.40 ± 1.10 | AE（MV) | Cycling (indoor), 90–95% LT, 30 min/session | 8*5 | supervised |  |  |  |
| Ben Cheikh et al. 2025^9^ | Tunisia | 16 | 26.40 ± 4.10 | 29.90 ± 3.10 | 37.40 ± 4.00 |  |  | AE（V） | Water–based aerobic exercise, RPE 9, 30 min/session | 10*3 | supervised | fat% | ST+  Siri/Jackson，% | ④ |
|  |  | 11 | 23.30 ± 1.40 | 30.10 ± 3.10 | 30.10 ± 3.10 |  |  | CON | Maintain a normal life |  |  |  |  |  |
| Aysha et al. 2025^10^ | India | 10 | 23.30 ± 2.50 | 27.60 ± 2.10 |  |  |  | AE（MV) | Treadmill backward walking, 4 km/h, 10% incline, 15–30 min/session | 12*4 | supervised | fat% | ST+  Siri/Jackson，% | ③④ |

|  |  | 10 | 25.40 ± 4.40 | 29.30 ± 2.70 |  |  |  | AE（MV) | Treadmill forward walking, 4 km/h, 10% incline, 15–30 min/session | 12*4 | supervised |  |  |  |
| --- | --- | --- | --- | --- | --- | --- | --- | --- | --- | --- | --- | --- | --- | --- |
|  |  | 10 | 27.00 ± 4.34 | 29.00 ± 2.70 |  |  |  | CON | Maintain a normal life |  |  |  |  |  |
| Rustaden et al. 2017^11^ | Norway | 25 | 39.00 ± 10.00 | 30.20 ± 5.40 | 38.70 ± 6.30 | 33.40 ± 11.20 |  | RT | Free–weight resistance training, 800 reps/session | 12*3 | supervised | fat% | Inbody，% | ①②④ |
|  |  | 25 | 38.00 ± 9.00 | 32.30 ± 6.10 | 41.10 ± 6.20 | 39.30 ± 14.60 |  | RT | Resistance training (BodyPump–style), load to 1RM | 12*3 | supervised |  |  |  |
|  |  | 21 | 42.00 ± 11.00 | 30.80 ± 4.90 | 38.40 ± 6.50 | 33.80 ± 10.80 |  | RT | Resistance training (BodyPump–style), load to 1RM | 12*3 |  |  |  |  |
|  |  | 21 | 40.00 ± 10.00 | 30.80 ± 5.00 | 20.80 ± 6.10 | 36.00 ± 11.20 |  | CON | Maintain a normal life |  |  |  |  |  |
| Ratajczak et al. 2019^12^ | Poland | 22 | 51.00 ± 8.00 | 35.90 ± 5.20 | ≥33% |  |  | AE（MV) | Cycling (stationary), 60–80% HRmax, 45 min/session | 12*3 | supervised | fat% | DEXA，% | ③④ |
|  |  | 17 | 49.00 ± 10.00 | 35.00 ± 3.90 | ≥33% |  |  | AE + RT | Resistance training + cycling (stationary), 60–80% HRmax, 20 + 25 min/session | 12*3 | supervised |  |  |  |
| Said et al. 2017^13^ | Saudi Arabia | 16 | 30.58 ± 3.80 | 32.07 ± 3.98 | 42.70 ± 4.69 | 33.79 ± 7.96 | 46.06 ± 7.10 | AE + RT | High–impact aerobics (jumping), 75–85% HRmax, 40 min/session | 24*4 | supervised |  |  |  |
|  |  | 16 | 29.66 ± 4.20 | 33.12 ± 2.04 | 43.94 ± 2.07 | 35.75 ± 7.80 | 47.42 ± 8.32 | AE + RT | Mixed training (aerobics + machine–based RT), 50–65% HRmax, 30 + 20 min/session | 24*4 | supervised |  |  |  |
| Arboleda–Serna et al. 2022^14^ | Colombia | 10 | 29.70 ± 7.20 | 30.60 ± 3.60 | 46.40 ± 4.00 |  |  | HIIT | HIIT, 90–95% HRmax (work) / 50–60% HRmax (recovery), 22 min/session | 8*3 | supervised | fat% | Omron  Healthcare，% | ③④ |
|  |  | 10 | 29.50 ± 8.10 | 29.70 ± 4.60 | 45.30 ± 4.50 |  |  | AE（M) | Steady–state aerobic exercise, 65–75% HRmax, 30 min/session | 8*3 | supervised |  |  |  |
| Abassi et al. 2023^15^ | Tunisia | 13 | 16.40 ± 1.20 | 33.10 ± 5.62 | 33.80 ± 2.94 |  |  | AE（M) | Aerobic exercise (MAS–based), 70–80% MAS, 6–8 × 30s bouts/set, 2 sets/session | 12*3 | supervised | fat% | BIA，% | ③④ |
|  |  | 13 | 16.40 ± 1.20 | 32.60 ± 3.61 | 33.70 ± 3.44 |  |  | HIIT | HIIT (MAS–based), 100–110% MAS, 6–8 × 30s bouts/set, 2 sets/session | 12*3 | supervised |  |  |  |
|  |  | 12 | 16.40 ± 1.20 | 33.20 ± 5.70 | 33.00 ± 3.09 |  |  | CON | Maintain a normal life |  |  |  |  |  |
| Gokalp et al. 2025^16^ | Turkey | 23 | 46.70 ± 9.77 | 29.09 ± 2.61 | 39.26 ± 4.92 |  |  | AE（MV) | Reformer Pilates, progressive intensity (low–high), 40 min/session | 8*3 | supervised | fat% | Inbody，% | ④ |
|  |  | 24 | 45.37 ± 9.63 | 29.00 ± 2.60 | 39.51 ± 4.96 |  |  | CON | Maintain a normal life |  |  |  |  |  |
| Kim et al. 2025^17^ | Korea | 13 | 31.00 ± 9.60 | 25.50 ± 1.70 | 37.30 ± 4.00 | 25.00 ± 4.40 | 42.00 ± 3.00 | WBVT | Whole–body vibration training, 50–75% HRmax, 40 min/session | 8*3 | supervised | fat% | Inbody，% | ①②③④ |
|  |  | 10 | 29.50 ± 5.10 | 25.80 ± 1.90 | 36.60 ± 3.70 | 25.40 ± 4.30 | 43.90 ± 4.10 | CON | Maintain a normal life |  | supervised |  |  |  |
| Zeng et al. 2021^18^ | China | 18 | 21.13 ± 1.64 |  | 30.18% ± 0.94 | 21.68 ± 3.08 | 50.08 ± 6.25 | AE（M) | Continuous aerobic training (FATmax), FATmax HR, 40 min/session | 12*3 | supervised | fat% | Inbody，% | ①② |
|  |  | 17 | 22.13 ± 1.96 |  | 30.31% ± 0.78 | 21.55 ± 2.65 | 49.47 ± 5.32 | HIIT | HIIT, 90% VO_2_max, 5 × 4 min work + 4 min rest, 40 min/session | 12*3 | supervised |  |  |  |
|  |  | 16 | 23.00 ± 2.07 |  | 30.17% ± 0.89 | 21.66 ± 2.29 | 50.12 ± 4.86 | RT | Resistance training, 15RM, 40 min/session | 12*3 | supervised |  |  |  |

| Goçer et al. 2017^19^ | Turkey | 14 | 46.60 ± 8.60 | 33.00 ± 3.50 |  |  |  | AE（MV) | Treadmill walking, 50%–70% VO_2_max, 30 min/session | 12*5 | supervised |  |  | ③④ |
| --- | --- | --- | --- | --- | --- | --- | --- | --- | --- | --- | --- | --- | --- | --- |
|  |  | 14 | 45.0 ± 9.80 | 32.0 ± 3.40 |  |  |  | AE（MV) | Walking with PEDO, 50%–70% VO_2_max, 30 min/session | 12*5 | supervised |  |  |  |
| Park et al. 2003^20^ | Korea | 10 | 42.20 ± 1.91 | 25.30 ± 1.74 |  |  |  | AE | Phased aerobic training, 60–70% HRmax, 60 min/session | 24*6 | supervised | fat% | BIA，% | ①④ |
|  |  | 10 | 43.40 ± 1.04 | 25.80 ± 1.43 |  |  |  | AE + RT | Combined training (aerobic + resistance), 60%–70% 1RM; aerobic moderate intensity, 60 min/session | 24*6 | supervised |  |  |  |
|  |  | 10 | 43.1 ± 1.67 | 25.50 ± 0.86 |  |  |  | CON | Maintain a normal life |  |  |  |  |  |
|  |  | 14 | 45.00 ± 9.80 | 32.00 ± 3.40 |  |  |  | AE（MV) | Brisk walking, 50%–70% VO_2_max, 30 min/session | 12*5 | supervised |  |  |  |
| Jamka et al. 2021^21^ | Poland | 44 | 55.00 ± 7.00 | 35.87 ± 4.43 |  |  |  | AE（M) | Cycle ergometer training, 50–70% HRmax, 60 min/session | 12*3 | supervised | fat% | Inbody，% | ③④ |
|  |  | 41 | 55.00 ± 7.00 | 35.98 ± 5.10 |  |  |  | AE + RT | Combined training (strength + cycling), 50–60% 1RM; 50–70% HRmax, 60 min/session | 12*3 | supervised |  |  |  |
| Sperlich et al. 2017^22^ | Germany | 11 | 23.00 ± 2.00 | 28.10 ± 2.70 |  | 40.00 ± 4.90 |  | HIIT | High–intensity circuit training (multi–joint functional), near–max HR, 30–45 min/session | 9*3 | supervised |  |  | ②③④ |
|  |  | 8 | 23.00 ± 2.00 | 28.30 ± 3.30 |  | 40.20 ± 4.80 |  | HIIT | Mixed–intensity training (circuit + steady–state cardio), near–max HR (high) / 65% HRmax (low), 45–75 min/session | 9*3 | supervised |  |  |  |
| Batrakoulis et al. 2018^23^ | Greece | 14 | 36.40 ± 5.00 | 28.20 ± 2.80 | 47.50 ± 3.20 |  | 40.80 ± 4.10 | AE + RT | Core + full–body circuit training, progressive duration 18–36 min, work–rest intervals, multi–muscle engagement | 40*3 | supervised | fat% | DEXA，% | ①②③④ |
|  |  | 14 | 36.90 ± 4.30 | 29.10 ± 3.00 | 46.20 ± 3.90 |  | 41.90 ± 3.20 | AE + RT | CINT training–detraining, Weeks 1–20: CINT (same as TR); Weeks 21–40: detraining (daily activity only) | 20*3 | supervised |  |  |  |
|  |  | 21 | 36.00 ± 4.20 | 29.60 ± 3.00 | 46.70 ± 6.50 |  | 42.80 ± 7.20 | CON | Maintain a normal life |  |  |  |  |  |
| Camacho–Cardenosa et  al. 2018^24^ | Spain | 13 | 43.14 ± 7.67 | 29.59 ± 5.25 | 38.89 ± 6.25 |  |  | AE（MV) | Aerobic HIIT (normoxia, FiO_2_=20.9%), 90% Wmax (exercise) + 55–65% Wmax(recovery), 41.5 min/session | 12*3 | supervised | fat% | TANITA，% | ②④ |
|  |  | 15 | 44.43 ± 7.18 | 30.03 ± 6.37 | 40.17 ± 7.20 |  |  | AE（MV) | Aerobic HIIT (hypoxia, FiO_2_=17.2%), 90%Wmax (exercise) + 55–65% Wmax(recovery), 41.5 min/session | 12*3 | supervised |  |  |  |
|  |  | 15 | 40.05 ± 8.66 | 28.74 ± 4.77 | 37.73 ± 5.28 |  |  | HIIT | Sprint–interval HIIT (normoxia, FiO_2_=20.9%), 130% Wmax (sprint) + 55–65% Wmax (recovery), 29.6 min/session | 12*3 | supervised |  |  |  |
|  |  | 18 | 37.40 ± 10.25 | 27.71 ± 4.55 | 37.65 ± 3.93 |  |  | HIIT | Sprint–interval HIIT (hypoxia, FiO_2_=17.2%),130% Wmax (sprint) + 55–65% Wmax(recovery), 29.6 min/session | 12*3 | supervised |  |  |  |
| Jung et al. 2020^25^ | Korea | 10 | 43.80 ± 8.60 | 25.10 ± 3.30 | 37.80 ± 5.00 |  | 43.20 ± 5.50 | RT | Resistance band Pilates (normoxia, FiO_2_=20.9%), standardized 25–movement protocol, 50 min/session, 3 sessions/week | 12*3 | supervised | fat% | DEXA，% | ①④ |
|  |  | 12 | 47.20 ± 6.40 | 27.10 ± 4.30 | 40.20 ± 6.40 |  | 41.30 ± 3.90 | RT | Resistance band Pilates (hypoxia, FiO_2_=14.5%, sim 3000 m), standardized 25–movement protocol, 50 min/session, 3 sessions/week | 12*3 | supervised |  |  |  |
|  |  | 10 | 51.60 ± 6.50 | 25.20 ± 2.00 | 37.00 ± 4.40 |  | 38.90 ± 4.50 | CON | Maintain a normal life |  |  |  |  |  |
| Hu et al. 2021^26^ | China | 15 | 20.90 ± 1.40 | 25.80 ± 2.60 | 38.00 ± 2.10 | 26.10 ± 3.70 | 42.50 ± 4.80 | AE（M) | Continuous cycling, 60% VO2peak, 65 min/session (200 kJ Weeks 1–4; 300 kJ Weeks 5–12) | 12*3 | supervised | fat% | DEXA，% | ②④ |
|  |  | 15 | 21.50 ± 1.70 | 25.50 ± 6.10 | 38.10 ± 2.30 | 38.10 ± 2.30 | 41.60 ± 3.50 | HIIT | Interval cycling, 90% VO2peak (exercise) + passive rest, 31 min/session (200 kJ Weeks 1–4; 300 kJ Weeks 5–12) | 12*3 | supervised |  |  |  |

|  |  | 15 | 21.40 ± 1.00 | 25.60 ± 2.30 | 38.40 ± 2.40 | 25.90 ± 3.40 | 41.40 ± 3.40 | HIIT | Repeated sprint cycling, all–out 6s sprint + 9s rest, 8 min/session, 150 kJ | 12*3 | supervised |  |  |  |
| --- | --- | --- | --- | --- | --- | --- | --- | --- | --- | --- | --- | --- | --- | --- |
|  |  | 15 | 20.90 ± 1.10 | 25.90 ± 2.40 | 40.90 ± 2.70 | 27.70 ± 4.80 | 39.80 ± 4.20 | CON | Maintain a normal life |  |  |  |  |  |
| Chainok et al. 2022^27^ | Thailand | 12 | 20.85 ± 1.97 | 31.20 ± 4.96 |  |  |  | AE + RT | Combined RT + AE (treadmill + resistance circuit), 25–27.5 min each, 50–55 min/session | 8*3 | supervised | fat% | BIA，% | ② |
|  |  | 12 | 20.67 ± 2.17 | 30.90 ± 3.72 |  |  |  | HIIT | Muay Thai HIIT, 10 sets (work period progressive + 3 min rest), 25–40 min/session, high intensity | 8*3 | supervised |  |  |  |
| Cakmakci et al. 2011^28^ | Turkey | 34 | 36.15 ± 9.59 | 33.76 ± 3.69 |  |  |  | AE + RT | Pilates (mat + ball), progressive intensity (HR 60%–70% HRmax), 1 h/session | 8*4 | supervised | fat% | BIA，% | ①③④ |
|  |  | 27 | 38.96 ± 10.02 | 32.46 ± 2.14 |  |  |  | CON | Maintain a normal life |  |  |  |  |  |
| Jamka et al. 2022^29^ | Poland | 21 | 51.00 ± 8.00 | 35.17 ± 3.86 | 46.90 ± 3.70 |  |  | AE（MV) | Cycle aerobic training (bicycle), 45 min/session, moderate–high intensity(50%–80% HRmax) | 12*3 | supervised |  |  | ①②④ |
|  |  | 17 | 48.00 ± 11.00 | 34.93 ± 3.82 | 46.10 ± 5.10 |  |  | AE + RT | Cycling aerobic training (cycle), 45 min/session, moderate–high intensity(50%–80% HRmax) | 12*3 | supervised |  |  |  |
| Sarsan et al. 2006^30^ | Turkey | 20 | 42.50 ± 10.07 | 33.73 ± 2.92 |  |  |  | AE（MV) | Aerobic training (brisk walking + cycling), 3–5 days/week, moderate–high intensity(50%–85% HRR) | 1–4week*3  5–8week*4  9–12week*5 | supervised |  |  | ③④ |
|  |  | 20 | 41.65 ± 7.62 | 35.38 ± 4.98 |  |  |  | RT | Resistance training (multi–muscle strength), 3 days/week, moderate–high intensity (40%–80% 1RM, progressive) | 12*3 | supervised |  |  |  |
|  |  | 20 | 43.60 ± 6.46 | 35.54 ± 3.67 |  |  |  | CON | Maintain a normal life |  |  |  |  |  |
| Arslan 2011^31^ | Turkey | 29 | 41.55 ± 6.72 | 33.99 ± 3.89 | 39.32 ± 2.64 |  |  | AE（ MV) | Step–aerobic dance (8 weeks), 3 times/week, 40–50 min/session, moderate–high intensity (50%–80% HRmax,progressive) | 8*3 | supervised | fat% | TANITA，% | ①③④ |
|  |  | 20 | 37.00 ± 9.09 | 32.30 ± 2.05 | 38.82 ± 3.41 |  |  | CON | Maintain a normal life |  |  |  |  |  |
| Domene et al. 2016^32^ | UK | 10 | 33.00 ± 11.00 | 26.70 ± 1.70 | 30.90 ± 5.50 |  |  | AE（V） | Latin–themed high–intensity aerobic dance, 1–2 times/week, 1 h/session, high intensity ( 6.7 METs, progressive frequency) | 1–4 week*1  5–8 week*2 | supervised | fat% | TANITA ，% | ④ |
|  |  | 10 | 35.00 ± 13.00 | 27.60 ± 2.00 | 31.70 ± 5.80 |  |  | CON | Maintain a normal life |  |  |  |  |  |
| Zhu et al. 2024^33^ | China | 10 | 20.60 ± 1.96 | 26.80 ± 2.20 | 32.50 ± 2.40 |  |  | HIIT | Anaerobic power bicycle interval training, 6s all–out sprint + 9s rest, 4 min/session, high intensity (49–66 kJ total work) | 1–4 week*3  5–12 week*4 | supervised | fat% | BIA，% | ④ |
|  |  | 10 | 19.80 ± 1.90 | 27.10 ± 3.90 | 32.70 ± 3.40 |  |  | HIIT | Aerobic power bicycle interval training, 1 min supramaximal work + 1.5 min rest, 16–21 min/session, supramaximal intensity(200 kJ total work) | 1–4 week*3  5–12 week*4 | supervised |  |  |  |

|  |  | 10 | 19.50 ± 1.00 | 27.40 ± 3.10 | 33.60 ± 3.10 |  |  | HIIT | Aerobic power bicycle interval training, 4  min submaximal work + 3 min rest, 20–28 min/session, submaximal intensity (200 kJ total work) | 1–4 week*3  5–12 week*4 | supervised |  |  |  |
| --- | --- | --- | --- | --- | --- | --- | --- | --- | --- | --- | --- | --- | --- | --- |
|  |  | 9 | 21.00 ± 2.50 | 25.10 ± 2.50 | 30.70 ± 3.00 |  |  | AE（M) | Aerobic power bicycle continuous training, 51–61 min/session, moderate intensity(200 kJ total work) | 1–4 week*3  5–12 week*4 | supervised |  |  |  |
|  |  | 9 | 21.00 ± 2.10 | 26.30 ± 1.80 | 31.30 ± 1.80 |  |  | CON | Maintain a normal life |  |  |  |  |  |
| Osalou et al. 2025^34^ | Turkey | 14 | 36.10 ± 3.38 | 31.67 ± 1.53 | 42.20 ± 3.50 | 35.69 ± 5.29 | 44.54 ± 5.51 | HIIT | HIIT (treadmill, 12 weeks), 3 times/week, 30 min/session, high intensity | 12*3 | supervised | fat% | Ohaus ，% | ①③④ |
|  |  | 10 | 36.10 ± 3.38 | 32.17 ± 1.46 | 42.19 ± 2.55 | 33.93 ± 3.50 | 42.98 ± 6.37 | CON | Maintain a normal life |  |  |  |  |  |
| Dupuit et al. 2022^35^ | France | 8 | 60.90 ± 4.80 | 30.30 ± 3.50 | 35.20 ± 4.90 |  |  | HIIT | Concurrent training (HIIT + resistance), moderate-to-high intensity, 60 min/session | 12*3 | supervised | fat% | DEXA，% | ①② |
|  |  | 8 | 58.80 ± 5.30 | 31.50 ± 3.40 | 33.00 ± 4.60 |  |  | CON | Maintain a normal life |  |  |  |  |  |
| Melinda et al. 2003^36^ | USA | 87 | 61.00 ± 6.91 | 30.50 ± 4.28 | 47.60 ± 4.99 | 38.50 ± 9.99 |  | AE + RT | Moderate-intensity aerobic + recommended resistance training, 40–75% HRmax, 45 min/session | 12*5 | supervised | fat% | DEXA，% | ②③④ |
|  |  | 86 | 60.60 ± 7.10 | 30.60 ± 3.78 | 47.40 ± 4.73 | 38.40 ± 8.52 |  | CON | stretching | 12*1 | supervised |  |  |  |
| Christine et al. 2010^37^ | Canada | 155 | 61.20 ± 5.40 | 29.10 ± 4.50 | 42.20 ± 4.90 | 30.90 ± 8.20 |  | AE (MV) | Aerobic treadmill or outdoor walking/jogging training, 60–85% HRmax, 45 min/session | 12*5 | supervised | fat% | DEXA，% | ②③④ |
|  |  | 152 | 60.60 ± 5.70 | 29.20 ± 4.30 | 42.40 ± 5.70 | 31.30 ± 8.60 |  | CON | Maintain a normal life |  |  |  |  |  |
| Paulo et al. 2019^38^ | Brazil | 12 | 62.90 ± 7.77 | 29.40 ± 4.24 | 44.00 ± 3.62 | 30.40 ± 6.98 | 36.30 ± 6.19 | HIIT | Bodyweight stair-squat interval training, >80% HRmax, 20–30 min/session | 12*3 | supervised | fat% | DEXA，% | ①④ |
|  |  | 12 | 63.00 ± 10.34 | 29.90 ± 3.62 | 44.30 ± 3.45 | 34.80 ± 13.51 | 36.00 ± 5.75 | AE + RT | Combined aerobic–resistance training, 70% HRmax/1RM, 60 min/session | 12*3 |  |  |  |  |
| Jessica et al. 2006^39^ | USA | 85 | 60.70±6.70 | 30.40 ± 4.10 | 47.50 ± 4.80 | 38.40 ± 9.60 | 39.60 ± 5.60 | AE (M) | Aerobic walking/jogging training, 60–75% HRmax, ≥45 min/session | 12*5 | supervised | fat% | DEXA，% | ① |
|  |  | 85 | 60.60±6.80 | 30.50 ± 3.70 | 47.40 ± 4.60 | 38.40 ± 8.40 | 39.90 ± 4.90 | CON | Stretching and flexibility training, low intensity, 45 min/session | 12*1 | supervised |  |  |  |
| Zhaleh et al. 2024^40^ | Iran | 12 | 29.50±3.70 | 35-50 | 42.99 ± 3.40 |  |  | HIIT | High-intensity interval running training, 85–90% HRmax, 40 min/session | 10*5 | supervised | fat% | BIA，% | ③④ |
|  |  | 12 | 30.00±2.50 |  | 43.70 ± 2.50 |  |  | HIIT | Combined HIIT and circuit resistance training, 70–80% 1RM, 40–60 min/session | 10*5 | supervised |  |  |  |
| Marine et al. 2019^41^ | France | 8 | 67.10 ± 7.20 | 31.20 ± 3.00 |  | 30.60 ± 5.30 | 49.80 ± 3.70 | AE (M) | Continuous cycling training, 55–60% peak power output, 40 min/session | 12*3 | supervised | fat% | DEXA，% | ①② |
|  |  | 10 | 59.90 ± 5.90 | 31.50 ± 4.30 |  | 27.60 ± 10.70 | 45.10 ± 15.60 | HIIT | High-intensity interval cycling training, 80–90% HRmax, 30 min/session | 12*3 | supervised |  |  |  |
|  |  | 9 | 61.10 ± 5.40 | 31.40 ± 4.00 |  | 28.10 ± 5.80 | 47.60 ± 4.20 | HIIT | Combined HIIT and resistance training, 70–80% 1RM, 40–60 min/session | 12*3 | supervised |  |  |  |
| Haifeng et al. 2021^42^ | China | 11 | 20.90 ± 1.70 | 25.60 ± 2.40 | 44.10 ± 4.10 |  |  | HIIT | All-out sprint interval cycling, >100% VO₂peak, 15 min/session | 12*3 | supervised | fat% | DEXA，% | ①② |
|  |  | 12 | 19.70 ± 1.30 | 26.10 ± 3.20 | 43.40 ± 4.80 |  |  | HIIT | Supramaximal sprint interval cycling, 120% VO₂peak, 20 min/session | 12*3 | supervised |  |  |  |
|  |  | 12 | 19.70 ± 1.10 | 26.00 ± 2.90 | 44.60 ± 5.00 |  |  | HIIT | Interval cycling, 90% VO₂peak, 25 min/session | 12*3 | supervised |  |  |  |
|  |  | 11 | 21.00 ± 2.40 | 25.10 ± 3.00 | 44.10 ± 4.50 |  |  | AE (M) | Moderate-intensity continuous cycling, 60% VO₂peak, 40 min/session | 12*3 | supervised |  |  |  |
|  |  | 13 | 21.20 ± 2.20 | 25.20 ± 1.80 | 43.50 ± 4.00 |  |  | CON | No exercise training |  |  |  |  |  |
| Elvis et al. 2013^43^ | Spain | 14 | 42.20 ± 5.80 | 28.30 ± 3.40 |  | 30.20 ± 6.60 | 43.00 ± 4.80 | AE (M) | Aerobic training, 60–70% HRmax, 60 min/session | 20*3 | supervised | fat% | DEXA，% | ①②③④ |
|  |  | 15 | 38.70 ± 6.20 | 29.70 ± 4.00 |  | 32.60 ± 8.30 | 43.40 ± 3.80 | RT | Resistance training, 60–70% 1RM, 60 min/session | 20*3 | supervised |  |  |  |
|  |  | 16 | 36.20 ± 7.20 | 27.90 ± 3.40 |  | 30.10 ± 7.90 | 42.90 ± 4.30 | AE + RT | Combined aerobic and resistance training, 60–70% HRmax, 60 min/session | 20*3 | supervised |  |  |  |
| Yati et al. 2019^44^ | Australia | 20 | 54.10 ± 3.60 | 28.30 ± 3.70 | 36.90 ± 4.30 |  |  | HIIT | Sprint interval training, >90% HRR, 20 min/session | 8*3 | supervised | fat% | DEXA，% | ①② |
|  |  | 20 | 53.30 ± 3.40 | 27.30 ± 4.10 | 36.50 ± 5.10 |  |  | CON | No exercise training |  |  |  |  |  |

BMI: body mass index, Fat%: body fat percentage, AE: aerobic exercise, RT: resistance exercise, HIIT: high-intensity interval training, WBVT: Whole body vibration training, CON: control, M: moderate intensity, V: vigorous intensity, MV: moderate to vigorous intensity, HR: heart rate, HRR: heart rate reserve, MHP: maximal heart period, HRmax: maximum heart rate, VO_2_max: maximal oxygen uptake, LT: lactate threshold, TRX: total resistance exercise, RPE: rating of perceived exertion, MAS: maximal aerobic speed, OMNI: OMNI scale of perceived exertion, FATmax: maximum fat oxidation, PEDO: Pedometer, FiO_2_: fraction of inspired oxygen, Wmax: maximal workload, HW: hydrostatic weighing, TANITA: TANITA body composition analyzer, BIA: bioelectrical impedance analysis, DEXA: dual-energy X-ray absorptiometry, ST + Siri/Jackson: skinfold thickness + Siri/Jackson equation, InBody: InBody body composition analyzer, Omron Healthcare: Omron body composition monitor, Ohaus: Ohaus weighing scale, ①lean body mass, ②fat mass, ③waist circumference, ④body mass index.

**Appendix 4.** List of included studies.

**Appendix 5.** Risk of bias assessment.

| Study | Random sequence generation | Allocation concealment | | Blinding of outcome assessors | Incomplete outcome | Selective outcome reporting | Other risks of bias | Risk category |
| --- | --- | --- | --- | --- | --- | --- | --- | --- |
| Abassi et al. 2023 | unclear | | unclear | low | low | low | low | low |
| Ajjimaporn et al. 2023 | low | | unclear | unclear | low | low | unclear | low |
| Alves et al. 2009 | unclear | | unclear | unclear | low | low | low | low |
| Arboleda-Serna et al. 2022 | low | | low | high | low | low | low | moderate |
| Arslan 2011 | low | | low | low | low | low | low | low |
| Aysha et al. 2025 | low | | low | unclear | low | low | low | low |
| Batrakoulis et al. 2018 | low | | low | unclear | low | low | low | low |
| Ben Cheikh et al. 2025 | low | | low | high | low | low | low | moderate |
| Cakmakçi 2011 | unclear | | unclear | unclear | low | low | unclear | moderate |
| Camacho-Cardenosa et al. 2018 | unclear | | unclear | low | low | low | unclear | low |
| Chainok et al. 2022 | unclear | | unclear | unclear | unclear | low | unclear | moderate |
| Domene et al. 2016 | low | | unclear | unclear | low | low | low | low |
| Gappmaier et al. 2006 | unclear | | unclear | unclear | low | low | low | low |
| Goçer et al. 2017 | unclear | | unclear | unclear | low | low | unclear | moderate |
| Gokalp et al. 2025 | low | | unclear | high | low | low | low | moderate |
| Hu et al. 2021 | low | | low | low | low | low | low | low |
| Jamka et al. 2021 | low | | low | low | low | low | low | low |
| Jamka et al. 2022 | low | | unclear | unclear | low | low | unclear | moderate |
| Jung et al. 2020 | low | | unclear | low | low | low | low | low |
| Kim et al. 2025 | low | | low | unclear | low | high | low | low |
| Lee et al. 2023 | low | | low | unclear | low | low | unclear | low |
| Lestari et al. 2025 | unclear | | unclear | unclear | low | low | unclear | moderate |
| Manthou et al. 2015 | low | | unclear | unclear | low | low | low | low |
| Osalou et al. 2025 | unclear | | unclear | low | high | low | low | low |
| Park et al. 2003 | high | | high | high | high | unclear | high | high |
| Ratajczak et al. 2019 | low | | unclear | unclear | low | low | low | low |
| Rustaden et al. 2017 | low | | low | low | low | low | low | low |
| Said et al. 2017 | low | | high | low | low | low | low | low |
| Sarsan et al. 2006 | low | | low | unclear | high | low | low | moderate |
| Sperlich et al. 2017 | unclear | | unclear | unclear | low | unclear | unclear | high |
| Tong et al. 2018 | unclear | | unclear | low | low | low | low | low |
| Zeng et al. 2021 | low | | unclear | unclear | low | low | unclear | low |
| Zhang et al. 2017 | unclear | | unclear | low | low | low | unclear | low |
| Zhu et al. 2024 | unclear | | unclear | unclear | unclear | low | high | high |
| Dupuit et al. 2022 | unclear | | unclear | low | low | low | low | moderate |
| Melinda et al. 2003 | low | | low | low | low | low | low | low |
| Christine et al. 2010 | low | | low | low | low | low | moderate | low |
| Paulo et al. 2019 | low | | unclear | unclear | low | low | low | moderate |
| Jessica et al. 2006 | low | | low | low | low | low | low | low |
| Zhaleh et al. 2024 | low | | low | unclear | low | low | low | low |
| Marine et al. 2019 | moderate | | moderate | low | low | low | low | moderate |
| Haifeng et al. 2021 | moderate | | moderate | low | low | low | low | moderate |
| Elvis et al. 2013 | low | | unclear | low | low | low | low | low |
| Yati et al. 2019 | unclear | | unclear | low | low | low | low | moderate |

**Appendix 6.** Contributions of direct and indirect comparisons to NMA and the number of studies of each direct comparison of body fat percentage, body mass index, lean body mass, fat mass, waist circumference.

**Note:** A-CON, B-RT, C-AE + RT, D-HIIT, E-AE(M), F-AE(MV), G-AE(V), H-WBVT.

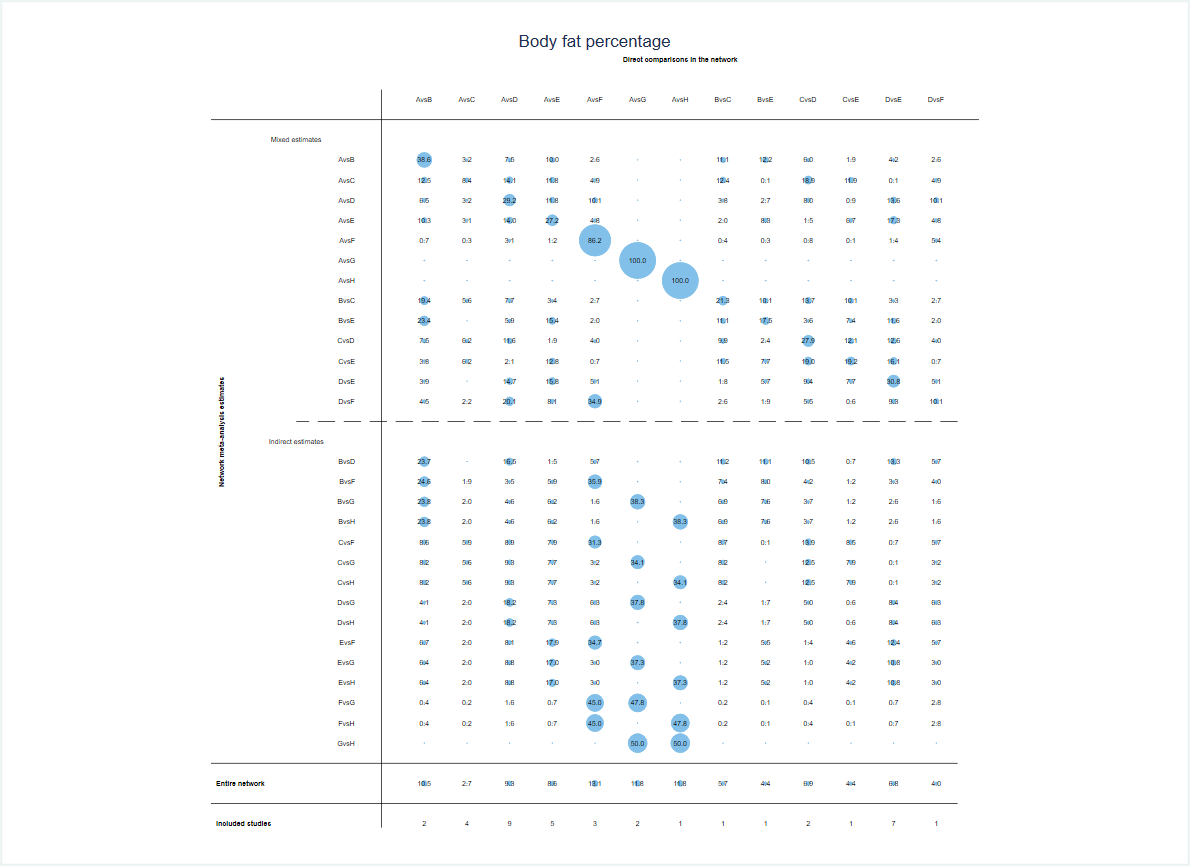

**Appendix 6-1** Body fat percentage.

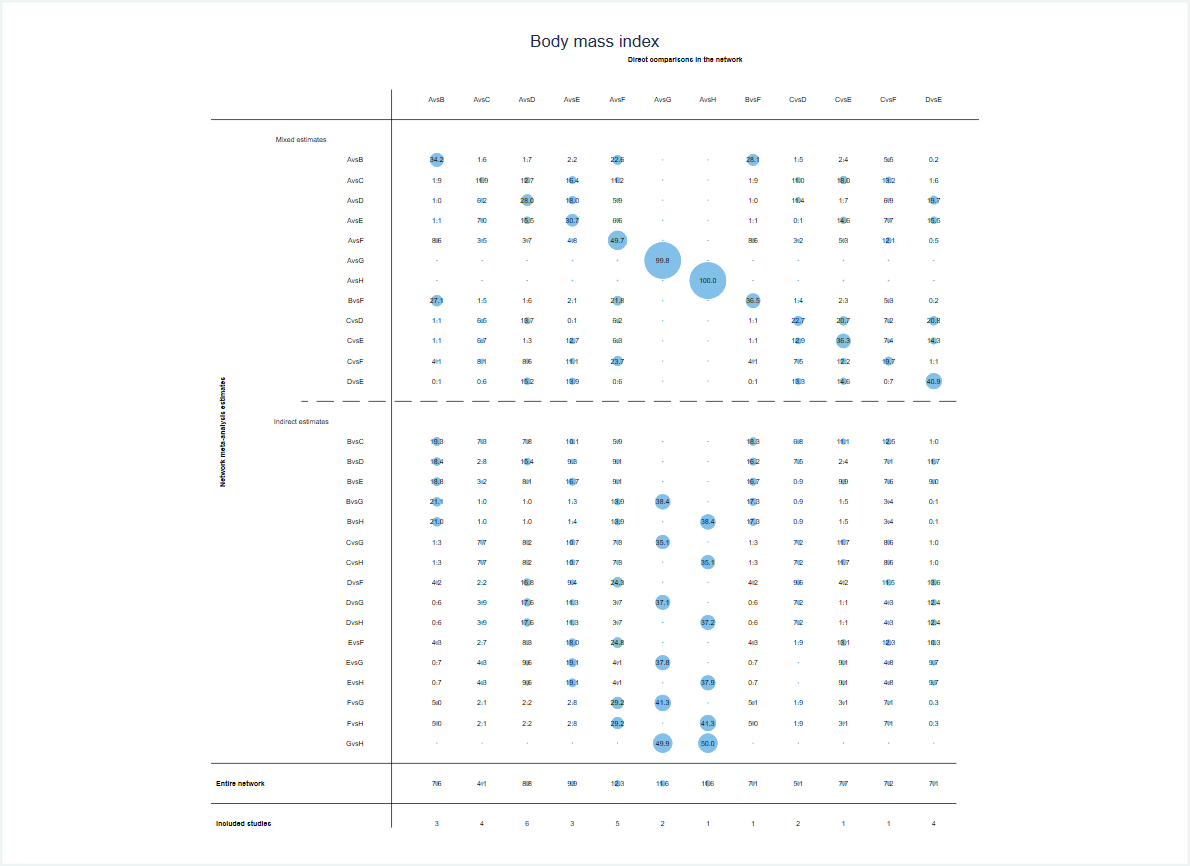

**Appendix 6-2** Body mass index.

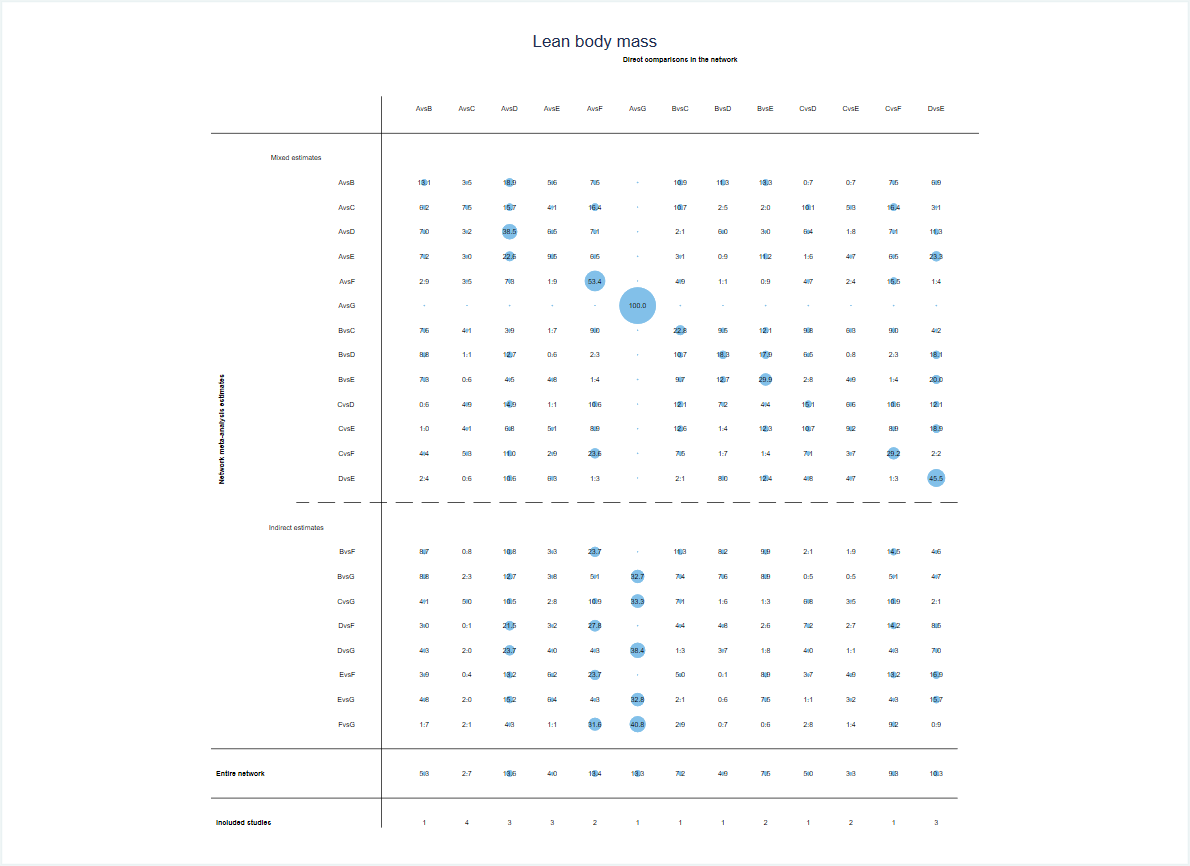

**Appendix 6-3** Lean body mass.

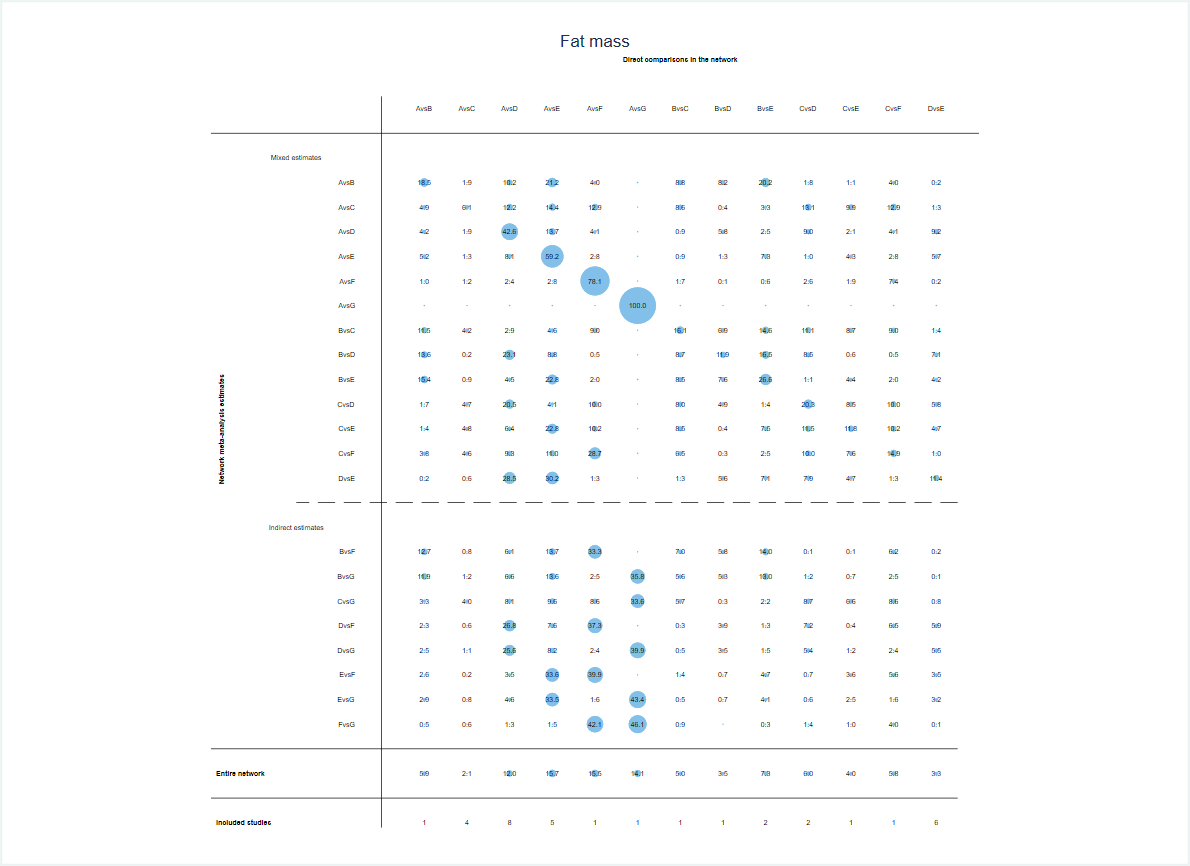

**Appendix 6-4** Fat mass.

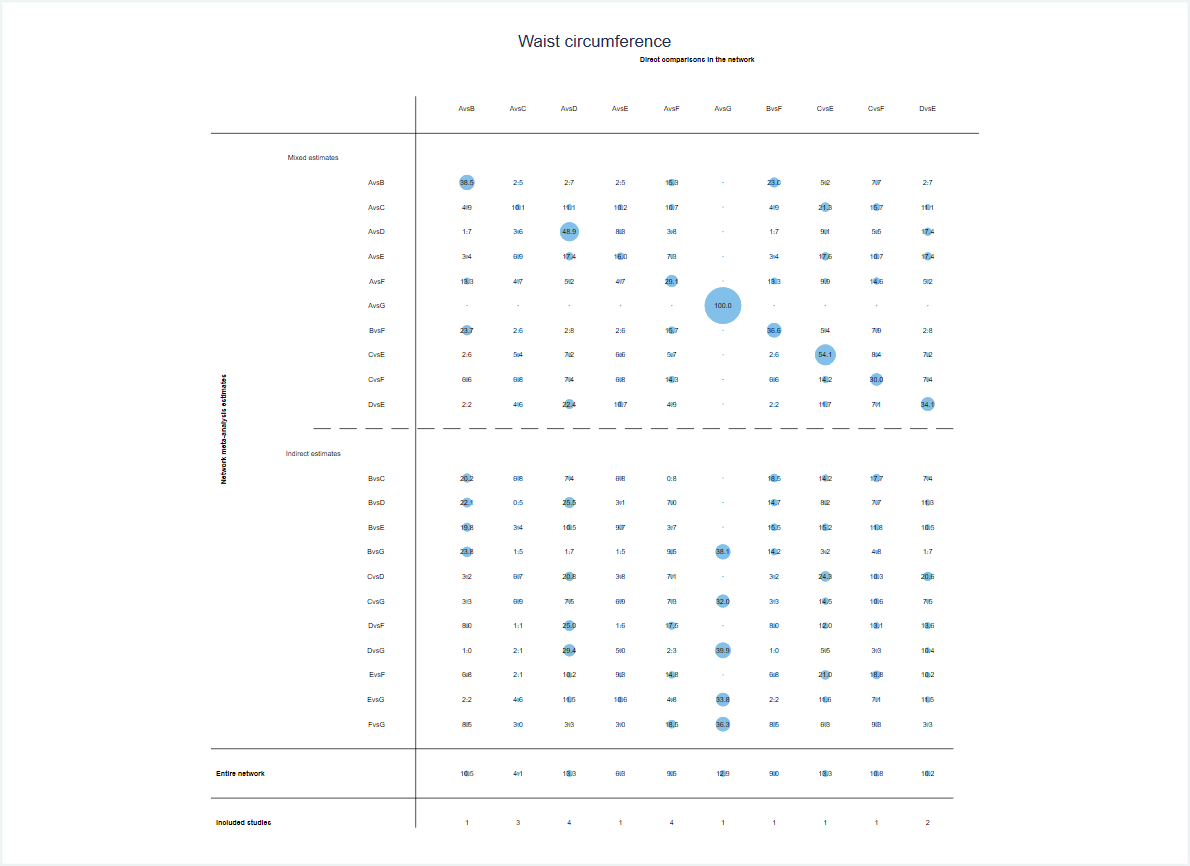

**Appendix 6-4** Waist circumference.

**Appendix 7.** Inconsistency of body fat percentage and secondary outcomes tested by loop-specific heterogeneity estimates, inconsistency model and node splitting analysis.

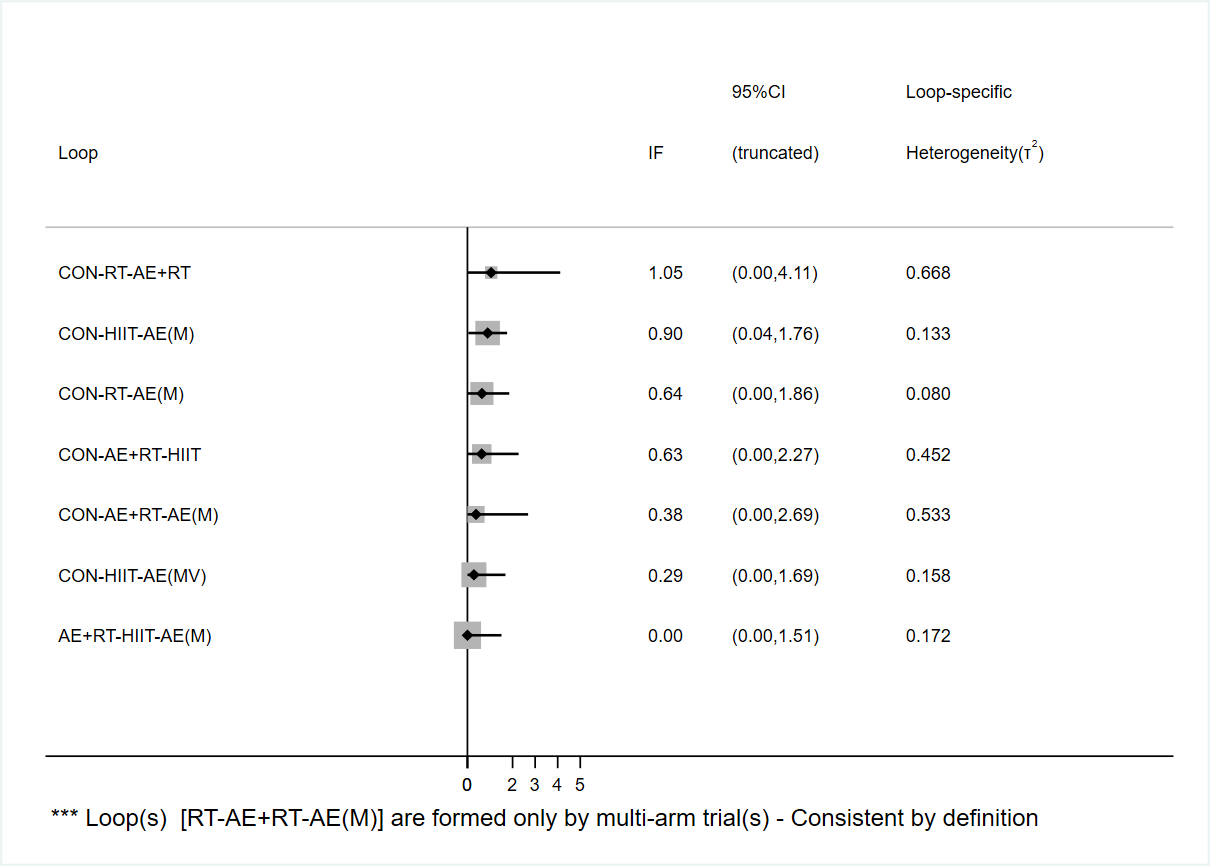

**Appendix 7-1** Body fat percentage.

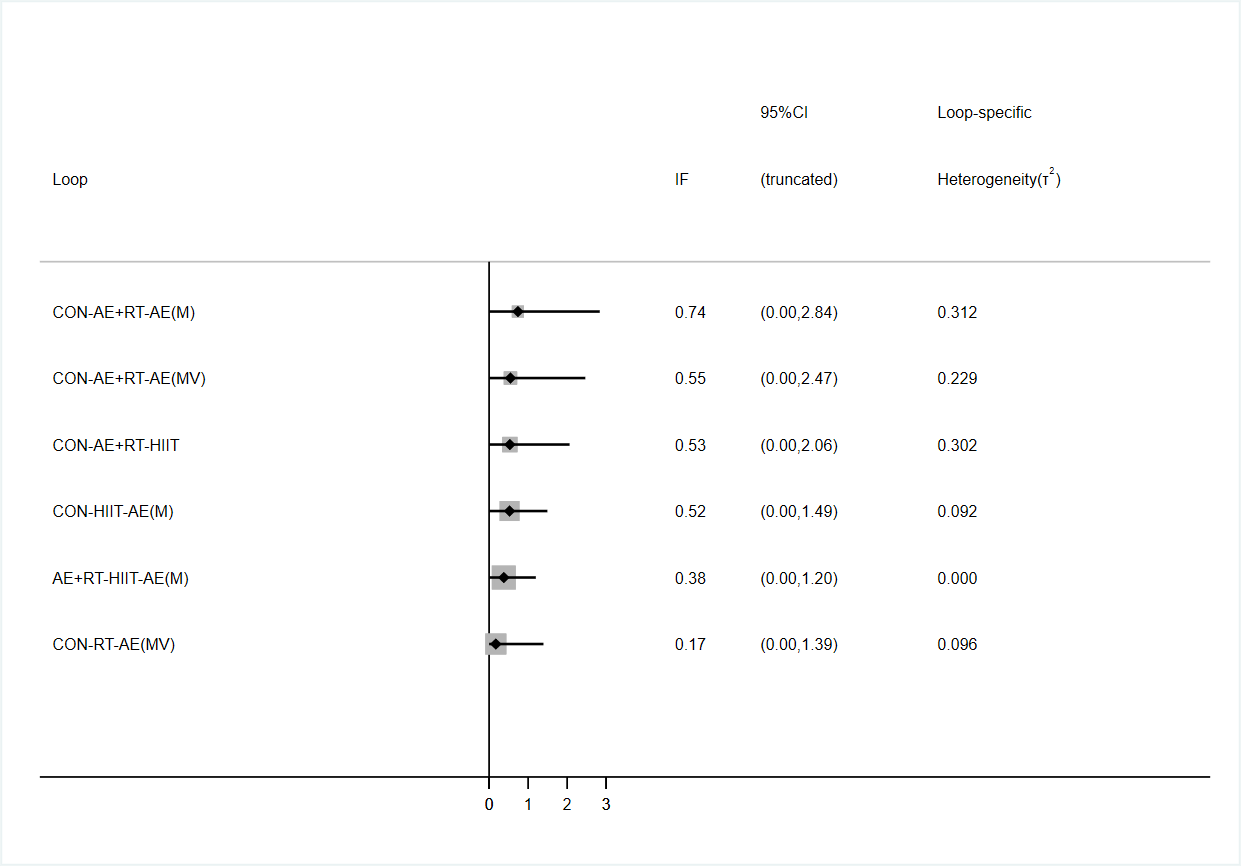

**Appendix 7-2** Body mass index.

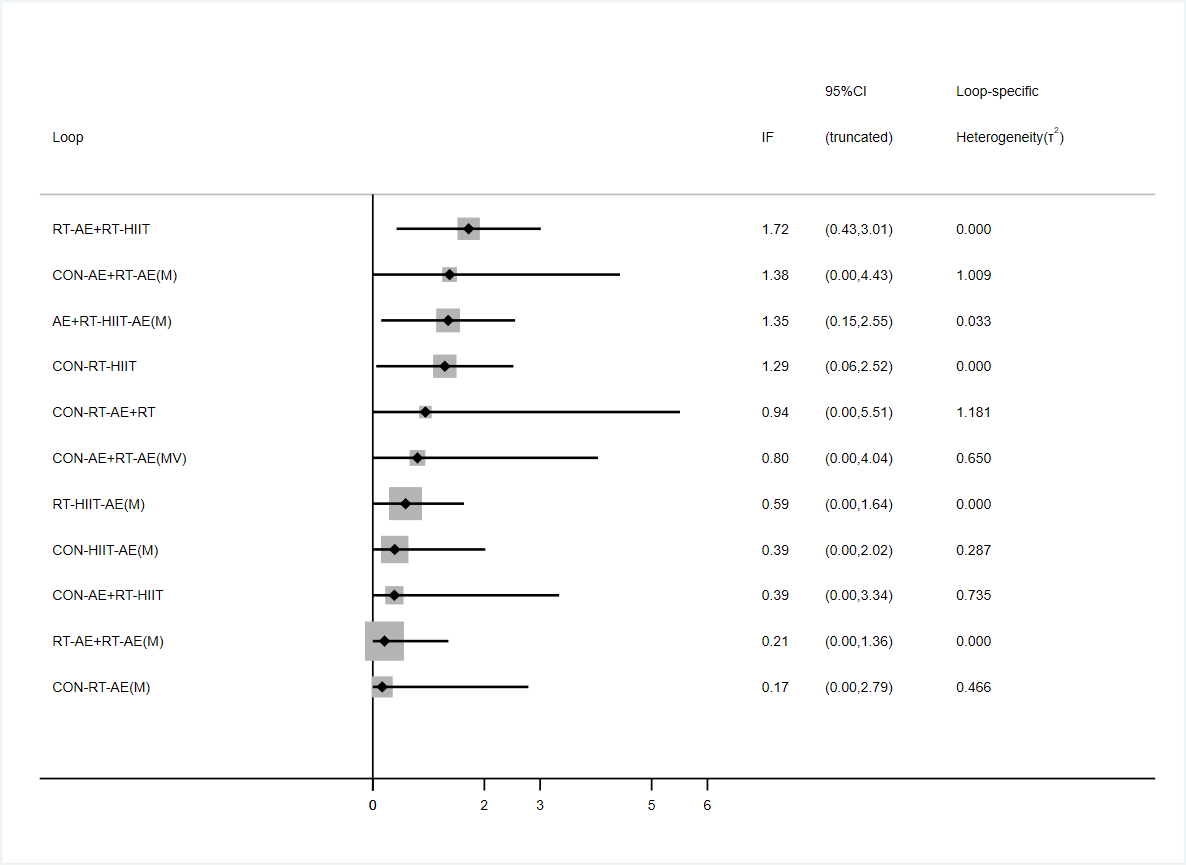

**Appendix 7-3** Lean body mass.

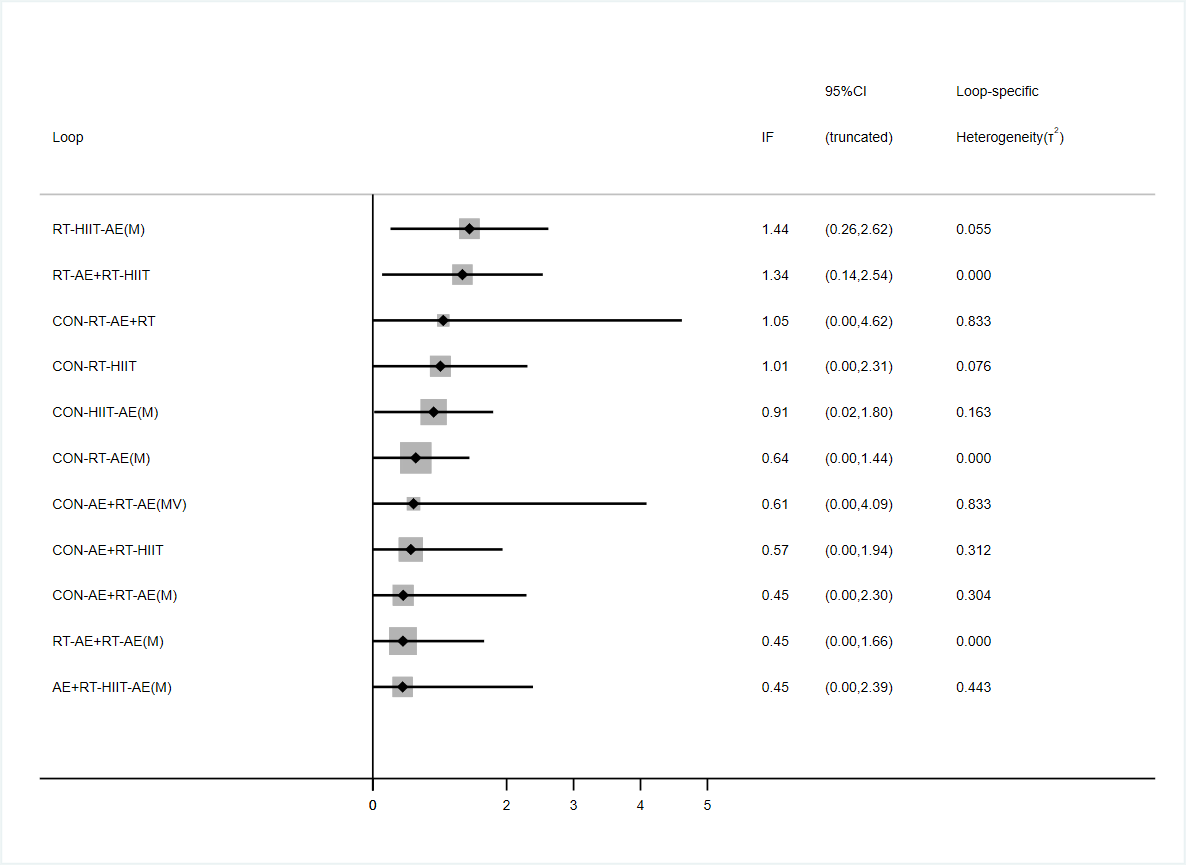

**Appendix 7-4** Fat mass.

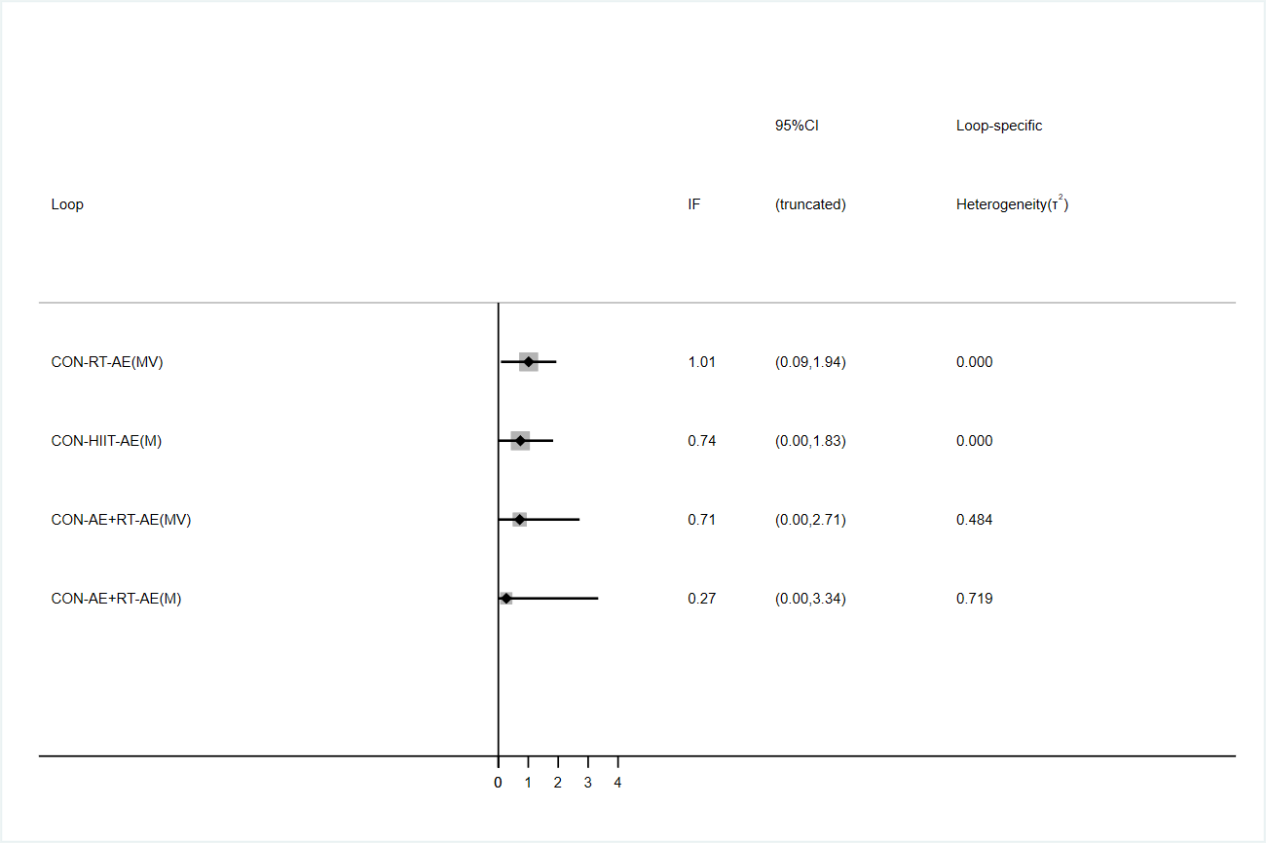

**Appendix 7-5** Waist circumference.

| **Inconsistency model** |  |  |  |  |  |
| --- | --- | --- | --- | --- | --- |
|  | **Body fat percentage** | **Body mass index** | **Lean body mass** | **Fat mass** | **Waist circumference** |
| chi2 | 11.67 | 3.73 | 3.42 | 12.05 | 2.42 |
| Prob＞chi2 | 0.07 | 0.81 | 0.98 | 0.21 | 0.79 |

**Appendix 7-6** Inconsistency model.

| Side | Direct |  | Indirect |  | Difference |  |  |
| --- | --- | --- | --- | --- | --- | --- | --- |
|  | Coef. | Std. Err. | Coef. | Std. Err. | Coef. | Std. Err. | P>z |
| **Body fat percentage** |  |  |  |  |  |  |  |
| A B | -0.025 | 0.420 | -0.621 | 0.548 | 0.596 | 0.690 | 0.388 |
| A C | -1.096 | 0.323 | -0.479 | 0.375 | -0.617 | 0.493 | 0.211 |
| A D | -0.901 | 0.213 | -0.862 | 0.380 | -0.039 | 0.434 | 0.928 |
| A E | -0.901 | 0.277 | -0.505 | 0.373 | -0.397 | 0.466 | 0.395 |
| A F | -0.819 | 0.313 | -1.115 | 0.640 | 0.295 | 0.712 | 0.678 |
| B C | -0.197 | 0.592 | -0.873 | 0.490 | 0.677 | 0.768 | 0.378 |
| B E | -0.259 | 0.601 | -0.675 | 0.475 | 0.417 | 0.766 | 0.587 |
| C D | -0.343 | 0.443 | 0.119 | 0.329 | -0.462 | 0.552 | 0.403 |
| C E | -0.071 | 0.603 | 0.140 | 0.344 | -0.211 | 0.694 | 0.761 |
| D E | 0.276 | 0.230 | -0.524 | 0.484 | 0.801 | 0.538 | 0.137 |
| D F | -0.202 | 0.610 | 0.093 | 0.367 | -0.295 | 0.712 | 0.678 |
| **Body mass index** |  |  |  |  |  |  |  |
| A B * | -0.231 | 0.385 | 0.601 | 1.271 | -0.832 | 1.331 | 0.532 |
| A C | -1.073 | 0.350 | -0.411 | 0.387 | -0.662 | 0.520 | 0.204 |
| A D | -0.665 | 0.290 | -1.055 | 0.539 | 0.391 | 0.611 | 0.522 |
| A E | -0.471 | 0.410 | -0.648 | 0.512 | 0.177 | 0.657 | 0.787 |
| A F | -0.429 | 0.295 | -0.533 | 0.652 | 0.104 | 0.717 | 0.884 |
| B F | -0.260 | 0.659 | -0.307 | 0.558 | 0.047 | 0.864 | 0.956 |
| C D | -0.128 | 0.499 | 0.139 | 0.384 | -0.266 | 0.630 | 0.672 |
| C E | -0.142 | 0.606 | 0.438 | 0.419 | -0.580 | 0.737 | 0.431 |
| C F | 0.033 | 0.661 | 0.465 | 0.408 | -0.432 | 0.777 | 0.578 |
| D E | 0.383 | 0.345 | -0.320 | 0.608 | 0.703 | 0.700 | 0.315 |
| **Lean body mass** |  |  |  |  |  |  |  |
| A B | 0.085 | 0.832 | -0.087 | 0.562 | 0.173 | 1.003 | 0.863 |
| A C | 0.645 | 0.414 | -0.220 | 0.496 | 0.865 | 0.646 | 0.181 |
| A D | 0.000 | 0.456 | 0.801 | 0.536 | -0.801 | 0.702 | 0.254 |
| A E | 0.400 | 0.460 | 0.539 | 0.548 | -0.140 | 0.710 | 0.844 |
| A F | -0.152 | 0.528 | 0.301 | 0.858 | -0.453 | 1.009 | 0.653 |
| B C | -0.222 | 0.760 | 0.711 | 0.609 | -0.933 | 0.974 | 0.338 |
| B D | 1.153 | 0.716 | -0.108 | 0.573 | 1.261 | 0.916 | 0.169 |
| B E | 0.474 | 0.562 | 0.524 | 0.819 | -0.050 | 0.992 | 0.960 |
| C D | -0.344 | 0.798 | 0.161 | 0.471 | -0.505 | 0.927 | 0.586 |
| C E | 0.869 | 0.523 | -0.460 | 0.484 | 1.329 | 0.716 | 0.063 |
| C F | -0.074 | 0.781 | -0.527 | 0.639 | 0.453 | 1.009 | 0.653 |
| D E | -0.175 | 0.447 | 0.631 | 0.616 | -0.805 | 0.761 | 0.290 |
| **Fat mass** |  |  |  |  |  |  |  |
| A B | 0.062 | 0.575 | -0.170 | 0.413 | 0.232 | 0.708 | 0.743 |
| A C | -1.154 | 0.330 | -0.603 | 0.350 | -0.551 | 0.481 | 0.251 |
| A D | -0.795 | 0.221 | -1.405 | 0.384 | 0.609 | 0.442 | 0.168 |
| A E | -0.695 | 0.250 | -0.049 | 0.359 | -0.646 | 0.438 | 0.141 |
| A F | -0.742 | 0.513 | -0.942 | 0.649 | 0.200 | 0.827 | 0.809 |
| B C | -0.343 | 0.603 | -1.080 | 0.462 | 0.737 | 0.759 | 0.331 |
| B D | -1.778 | 0.581 | -0.450 | 0.382 | -1.328 | 0.700 | 0.058 |
| B E | -0.071 | 0.408 | -0.930 | 0.526 | 0.859 | 0.666 | 0.197 |
| C D | -0.178 | 0.455 | 0.022 | 0.328 | -0.200 | 0.561 | 0.722 |
| C E | 0.034 | 0.610 | 0.530 | 0.330 | -0.496 | 0.694 | 0.475 |
| C F | -0.015 | 0.597 | 0.185 | 0.573 | -0.200 | 0.827 | 0.809 |
| D E | 0.650 | 0.251 | -0.069 | 0.424 | 0.719 | 0.493 | 0.145 |
| **Waist circumference** |  |  |  |  |  |  |  |
| A B * | -0.582 | 0.609 | 1.421 | 1.244 | -2.003 | 1.395 | 0.151 |
| A C | -0.527 | 0.371 | -0.905 | 0.544 | 0.379 | 0.658 | 0.565 |
| A D | -0.424 | 0.357 | -0.924 | 1.036 | 0.500 | 1.096 | 0.648 |
| A E | -0.681 | 0.706 | -0.209 | 0.530 | -0.471 | 0.884 | 0.594 |
| A F * | -0.867 | 0.318 | -0.110 | 0.718 | -0.757 | 0.785 | 0.335 |
| B F * | -0.960 | 0.614 | 1.043 | 1.238 | -2.003 | 1.395 | 0.151 |
| C E | 0.164 | 0.629 | 0.378 | 0.638 | -0.213 | 0.896 | 0.812 |
| C F | 0.400 | 0.639 | -0.357 | 0.456 | 0.757 | 0.785 | 0.335 |
| D E | 0.287 | 0.512 | -0.342 | 0.783 | 0.628 | 0.936 | 0.502 |

**Note:** A-CON, B-RT, C-AE + RT, D-HIIT, E-AE(M), F-AE(MV), G-AE(V), H-WBVT.

* All the evidence about these contrasts comes from the trials which directly compare them.

**Appendix 7-7** Node splitting analysis.

**Appendix 8.** Forest plots of eligible comparisons of body fat percentage, body mass index, lean body mass, fat mass, waist circumference.

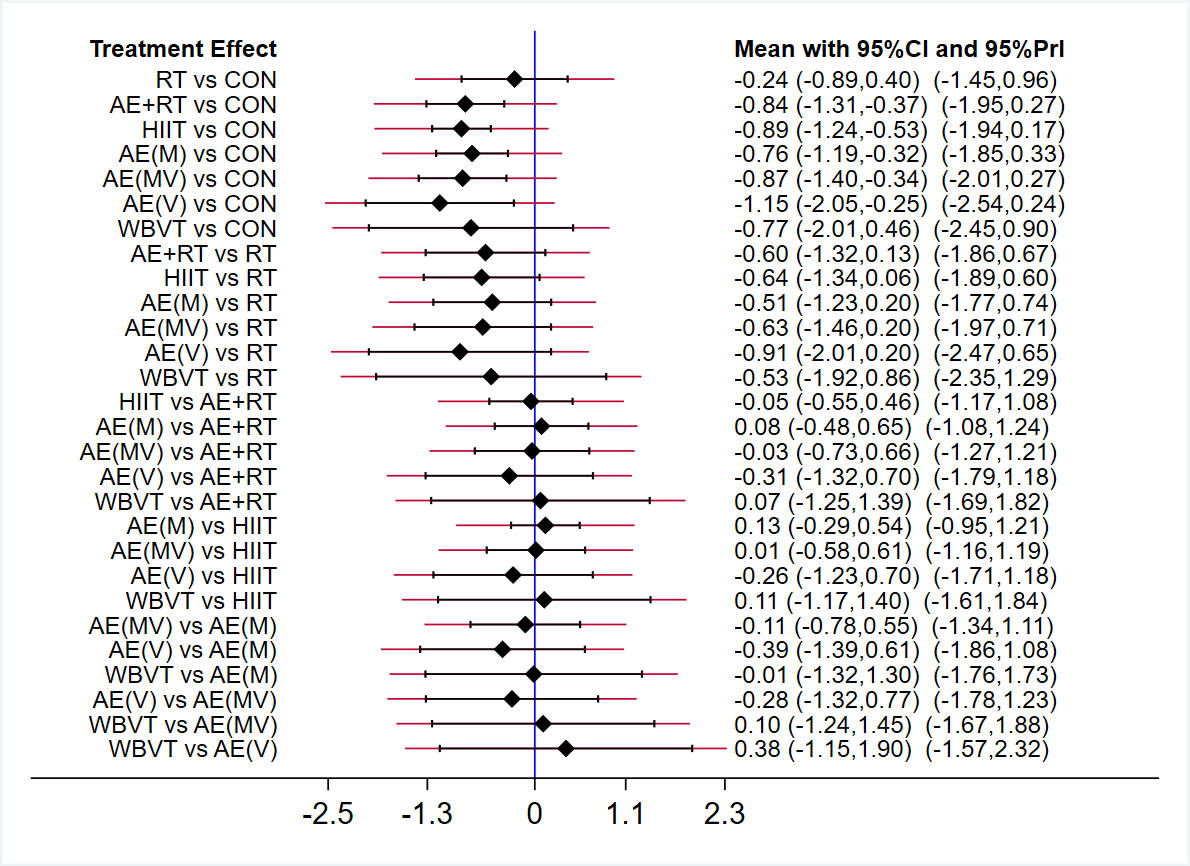

**Appendix 8-1** Body fat percentage.

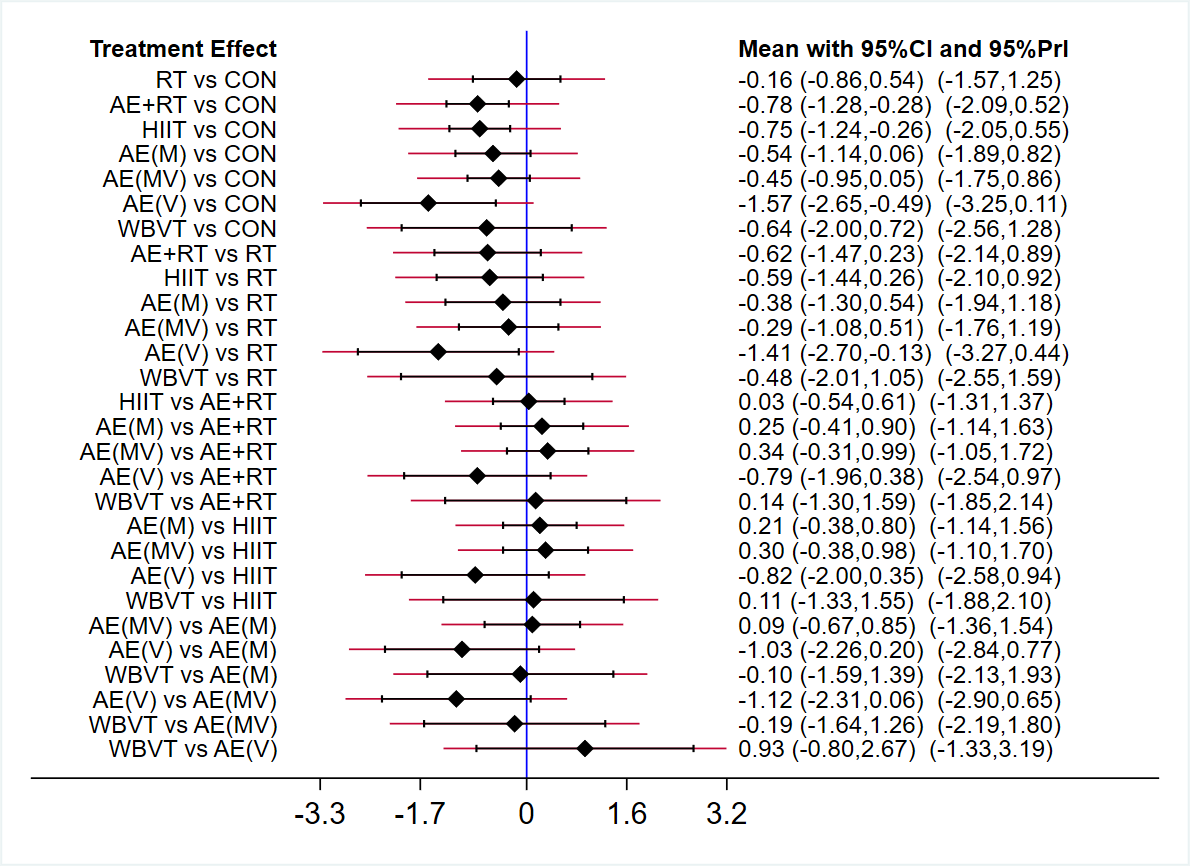

**Appendix 8-2** Body mass index.

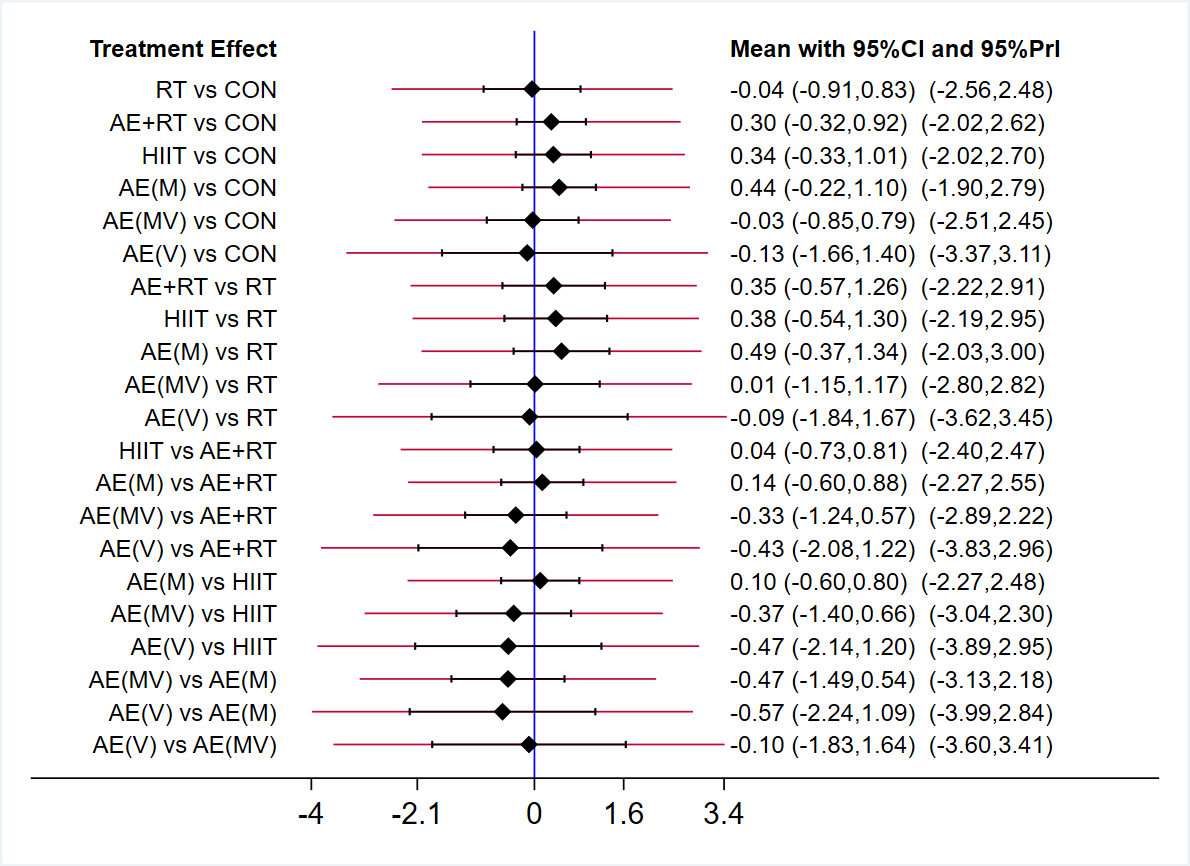

**Appendix 8-3** Lean body mass.

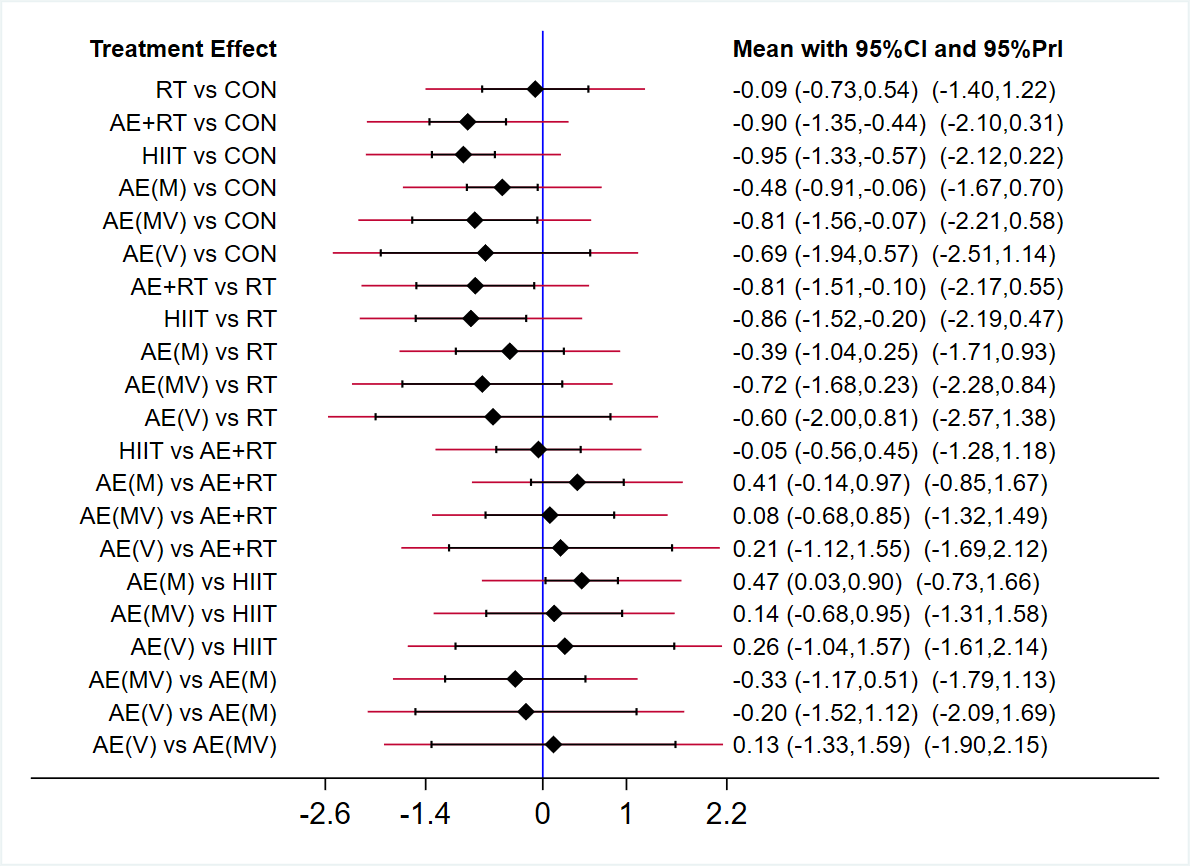

**Appendix 8-4** Fat mass.

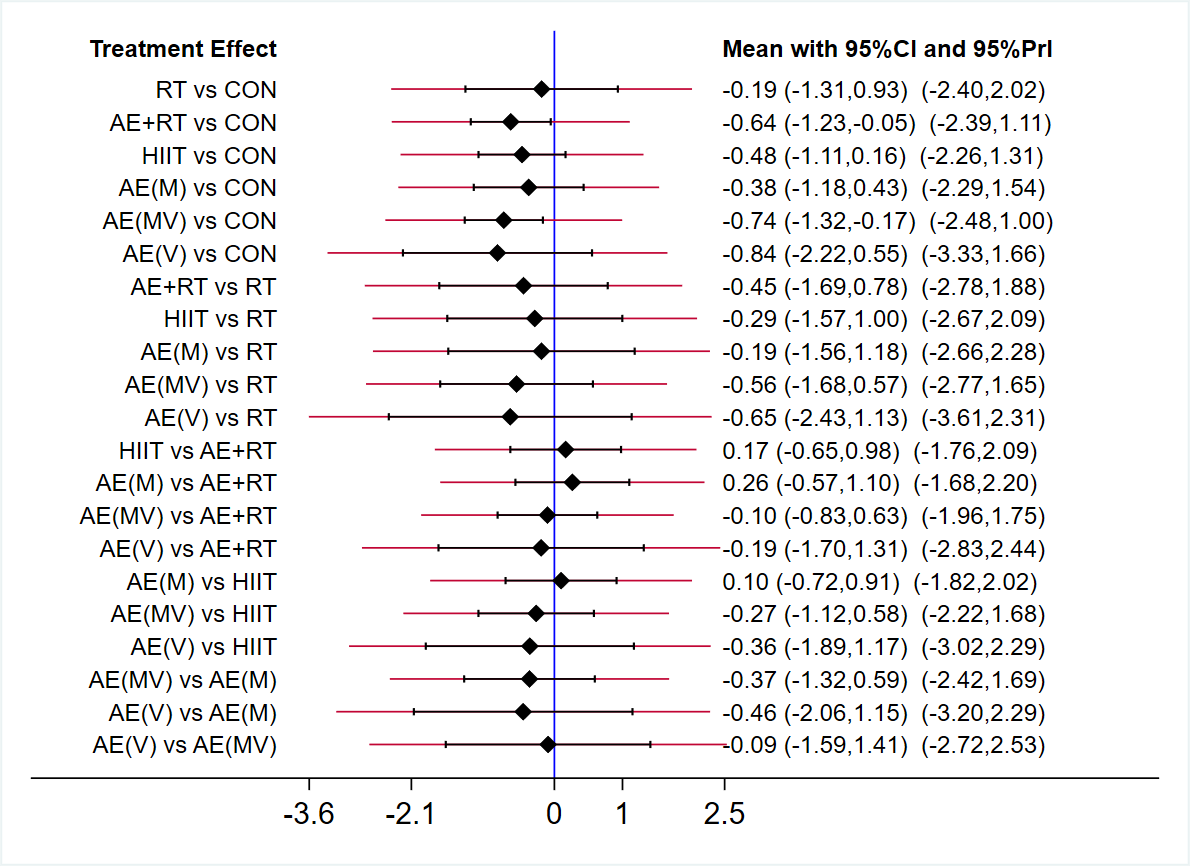

**Appendix 8-5** Waist circumference.

**Appendix 9.** Funnel plot of body fat percentage, body mass index, lean body mass, fat mass, waist circumference in network meta-analysis.

**Note:** A-CON, B-RT, C-AE + RT, D-HIIT, E-AE(M), F-AE(MV), G-AE(V), H-WBVT.

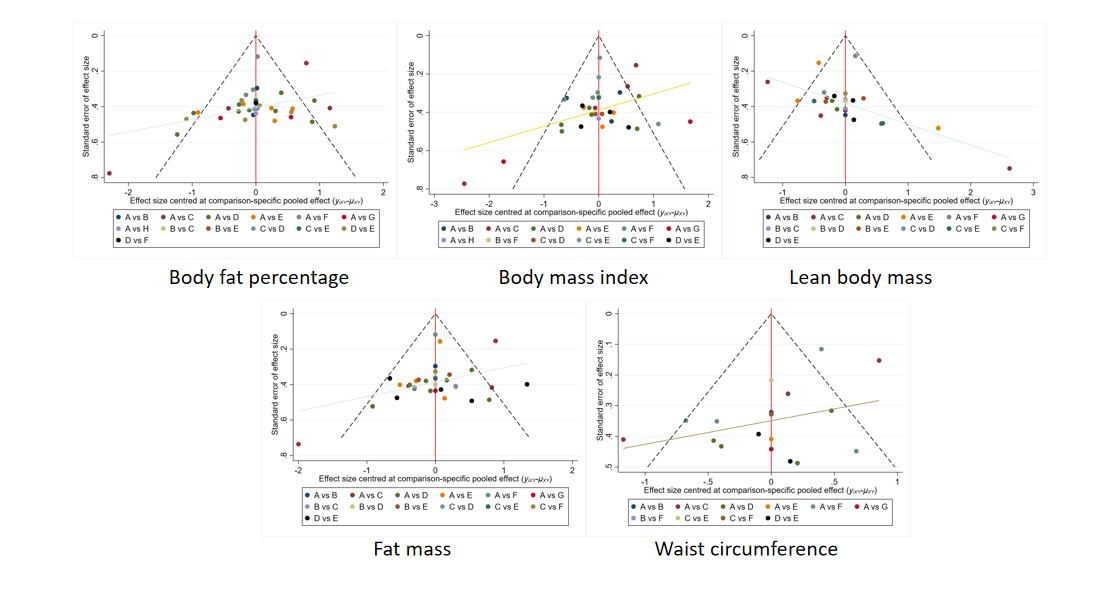

**Appendix 10.** Area under the curve for cumulative ranking probability of each intervention on body fat percentage, body mass index, lean body mass, fat mass, waist circumference.

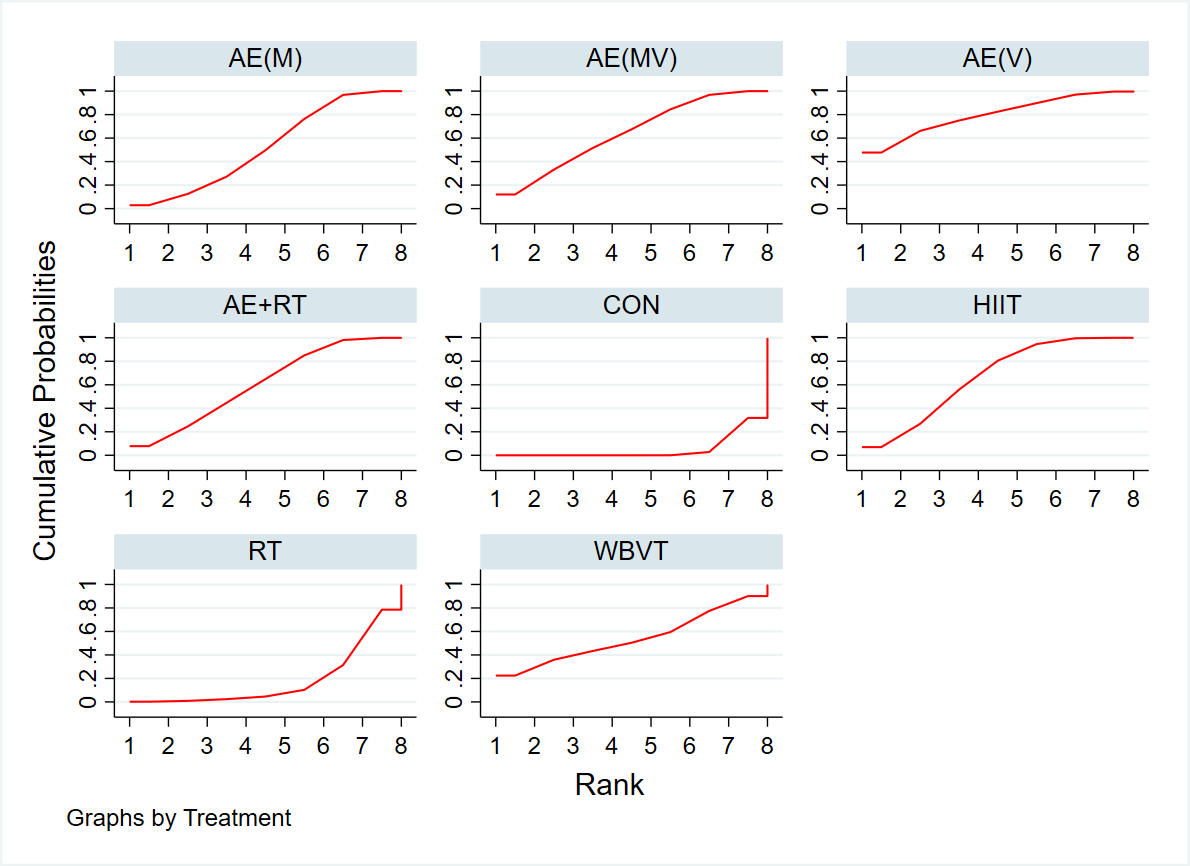

**Appendix 10-1** Body fat percentage.

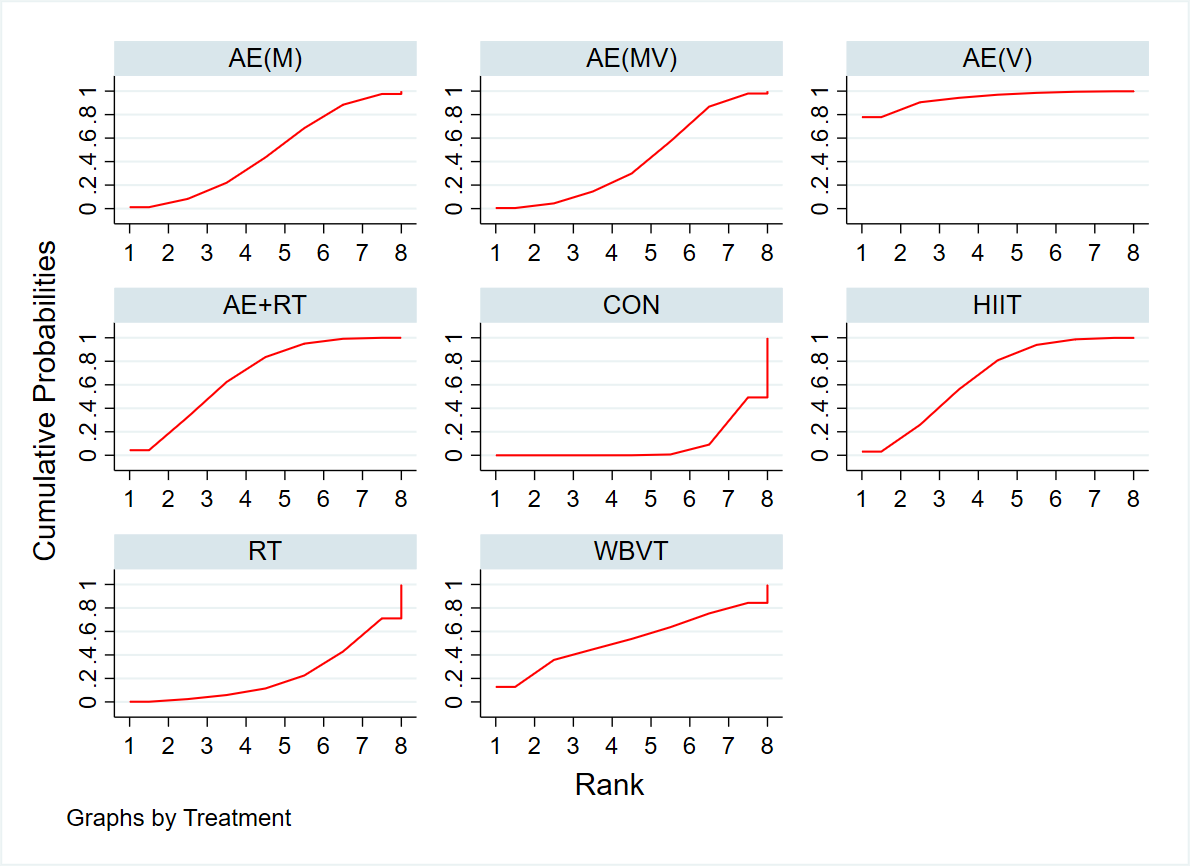

**Appendix 10-2** Body mass index.

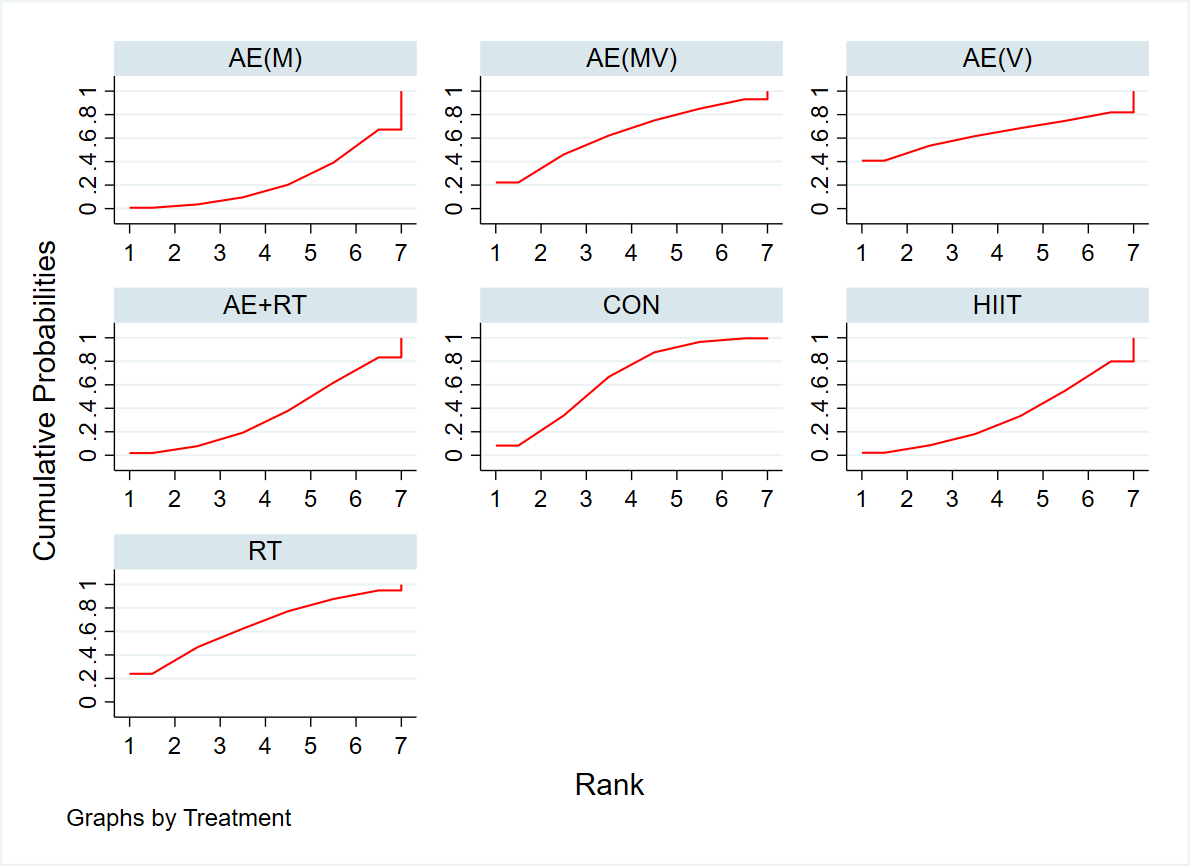

**Appendix 10-3** Lean body mass.

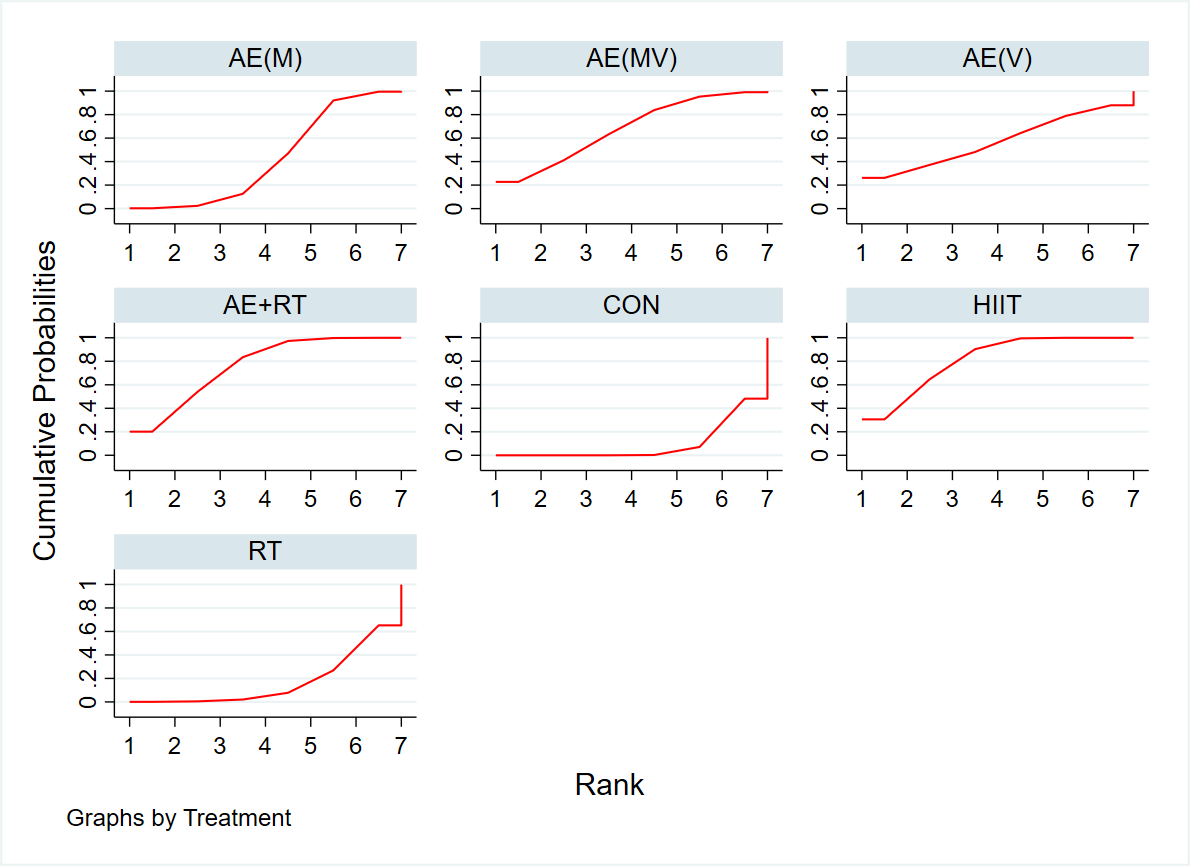

**Appendix 10-3** Fat mass.

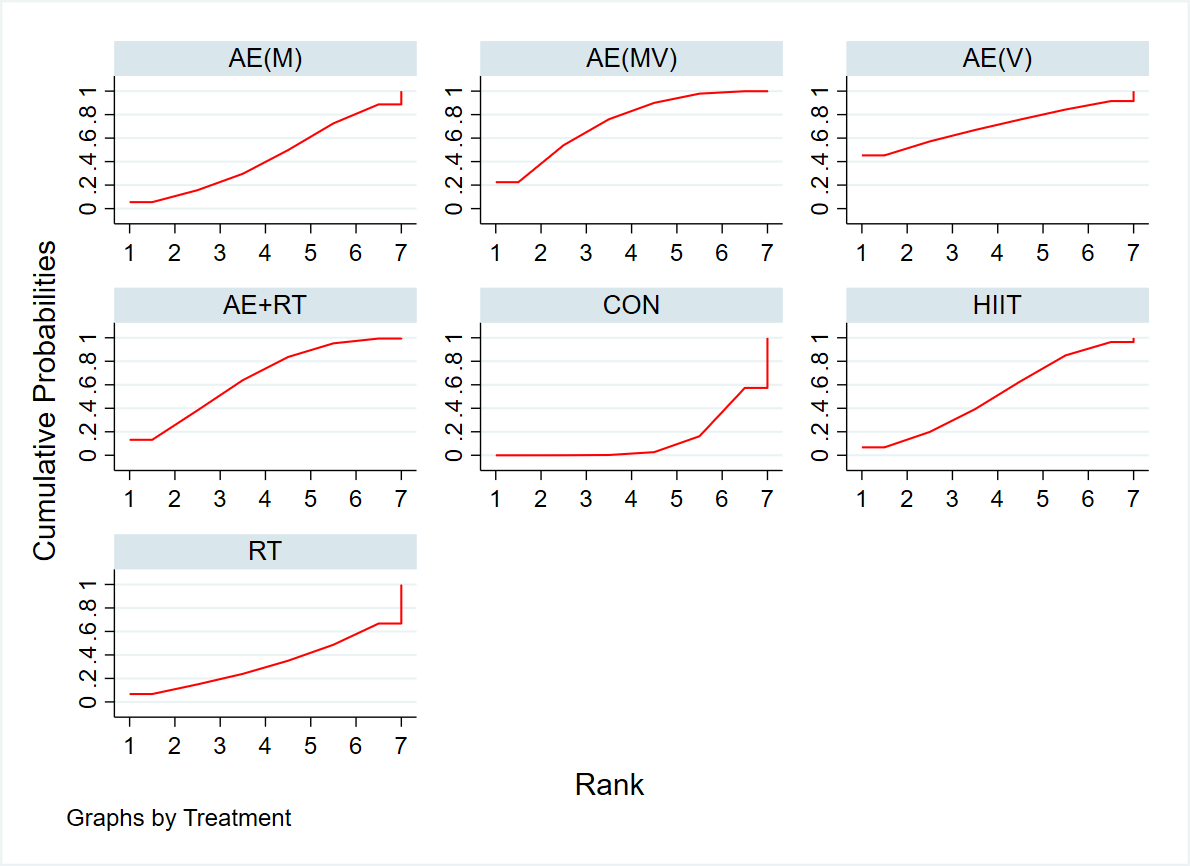

**Appendix 10-4** Waist circumference.

**Appendix 11.** Forest plot of individual studies.

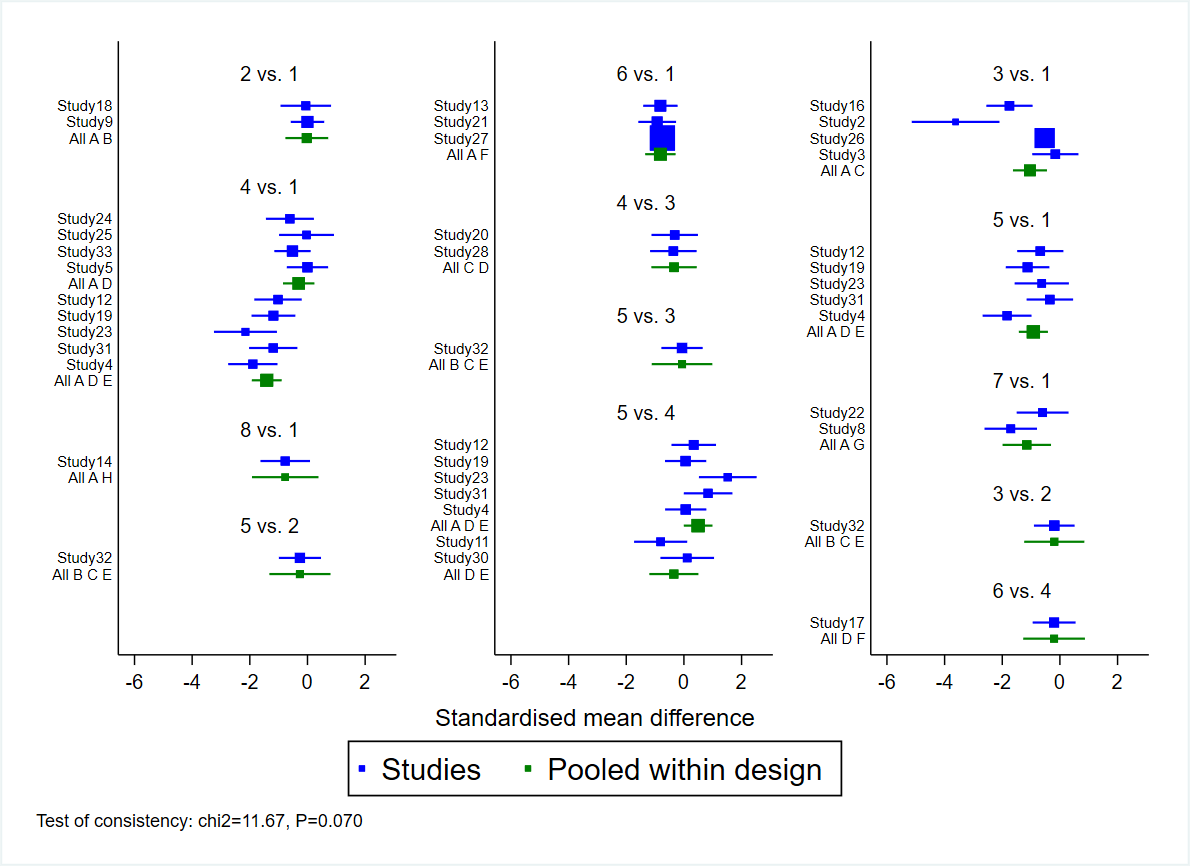

**Appendix 11-1** Body fat percentage.

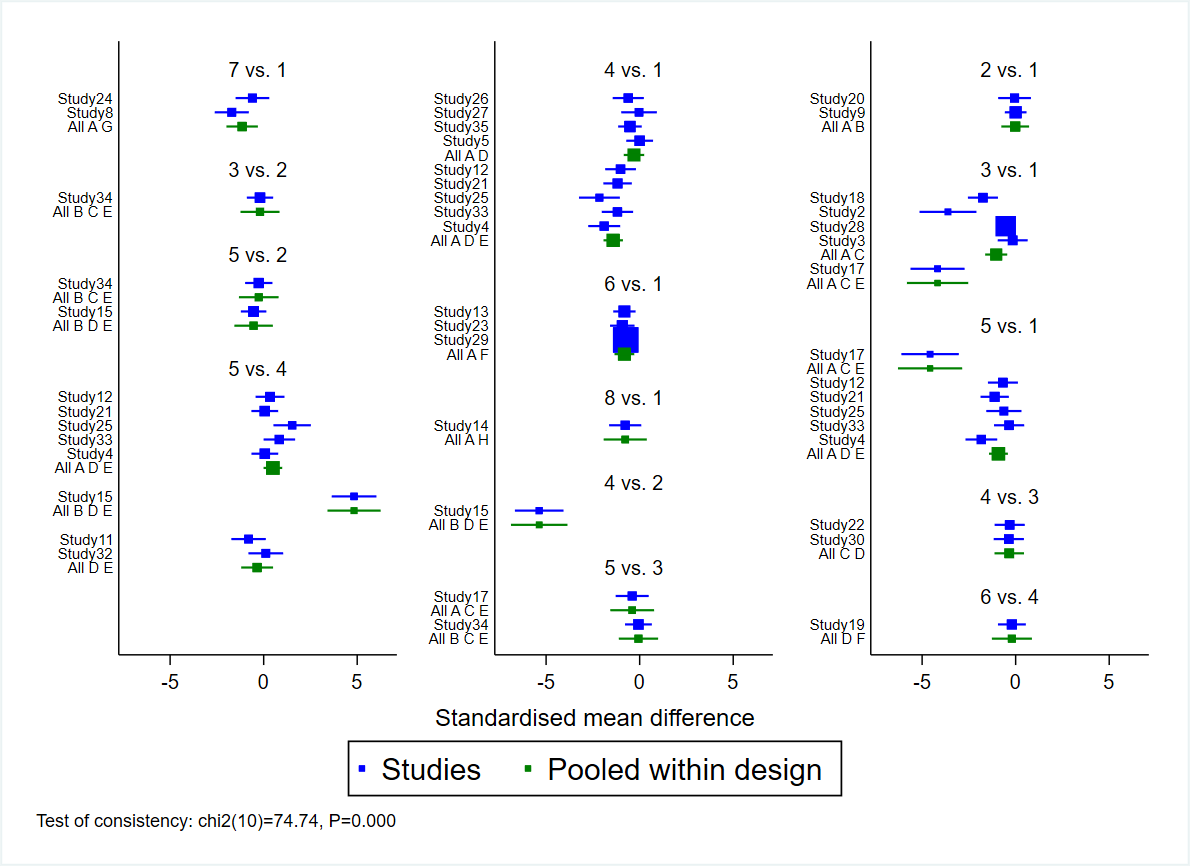

**Appendix 11-2** Body fat percentage (before study exclusion).

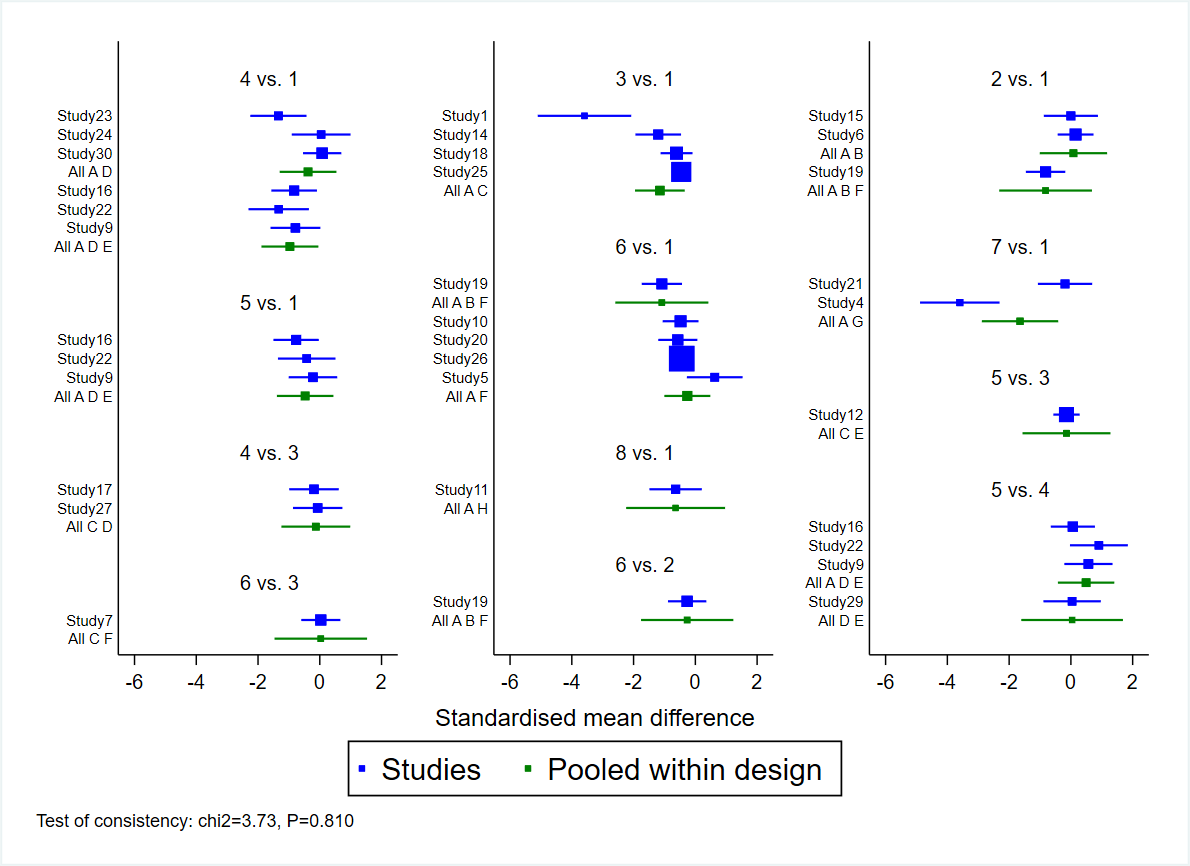

**Appendix 11-3** Body mass index.

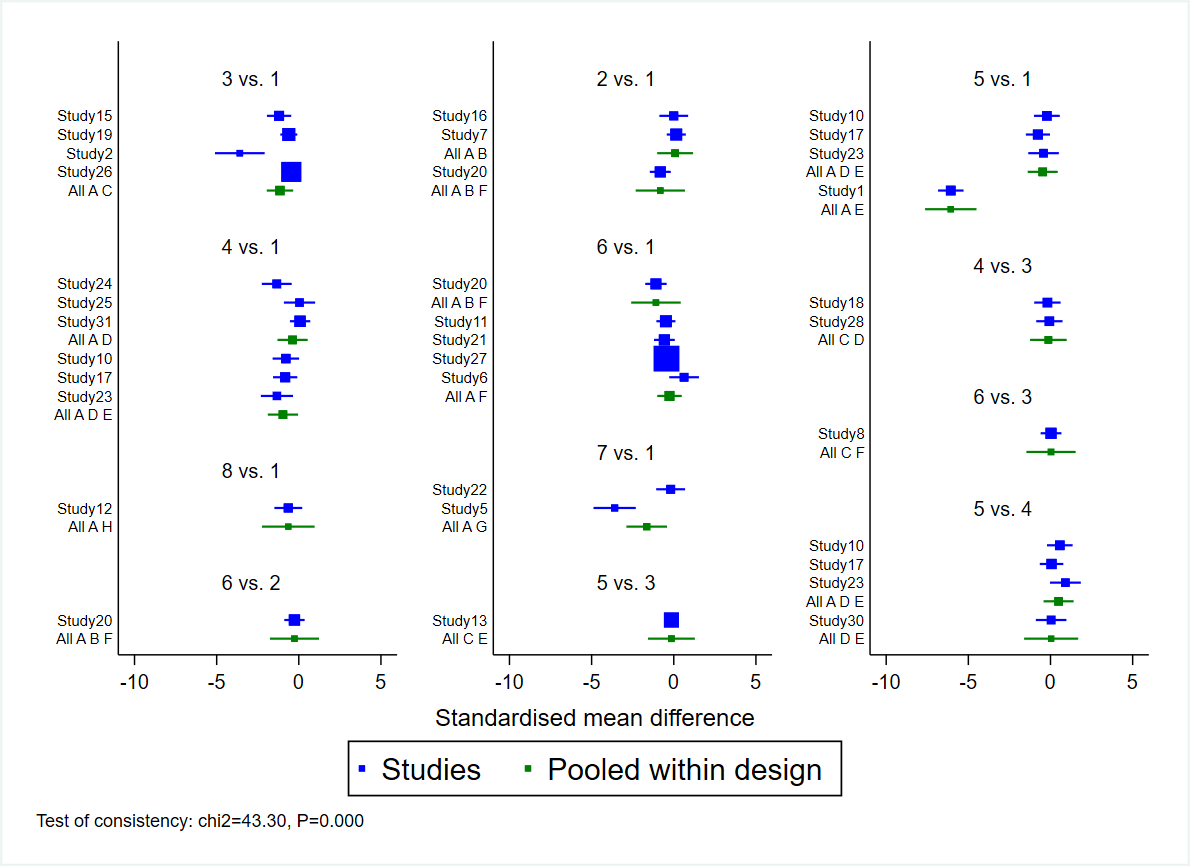

**Appendix 11-4** Body mass index (before study exclusion).

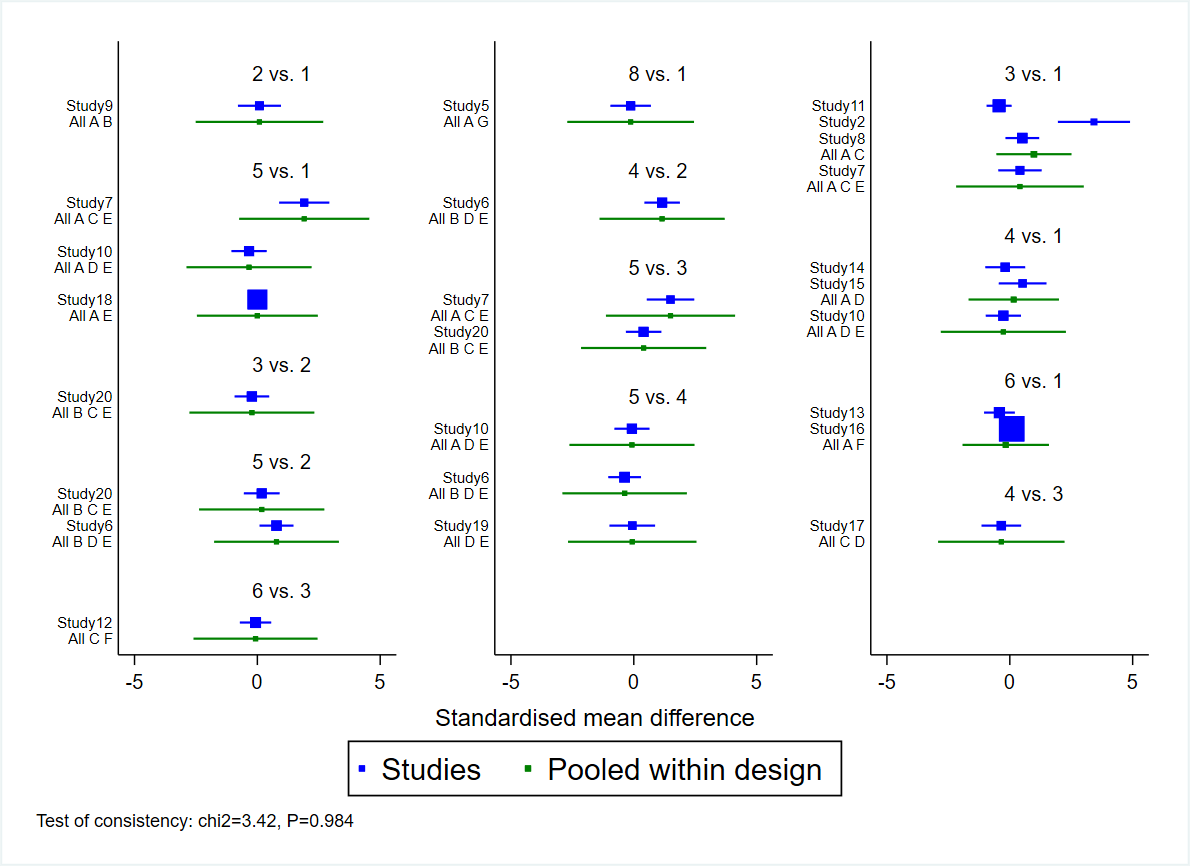

**Appendix 11-5** Lean body mass.

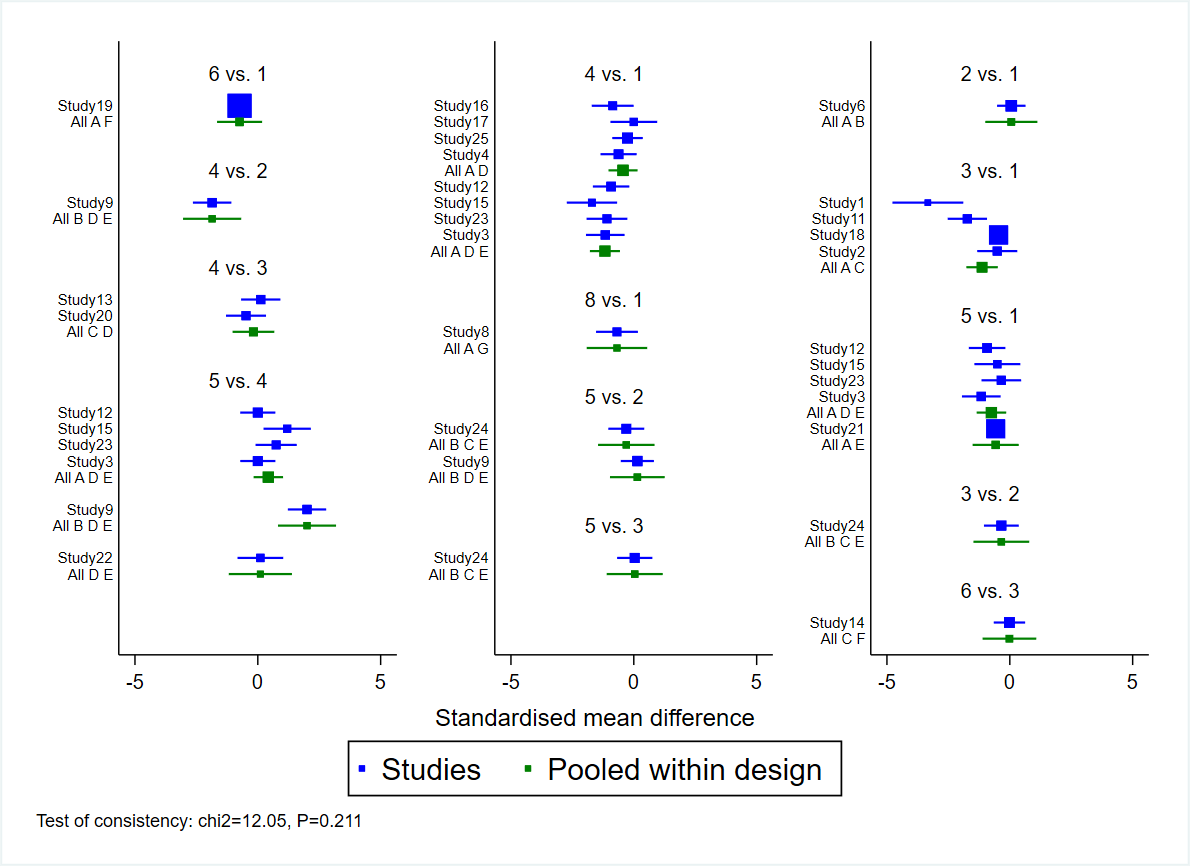

**Appendix 11-6** Fat mass.

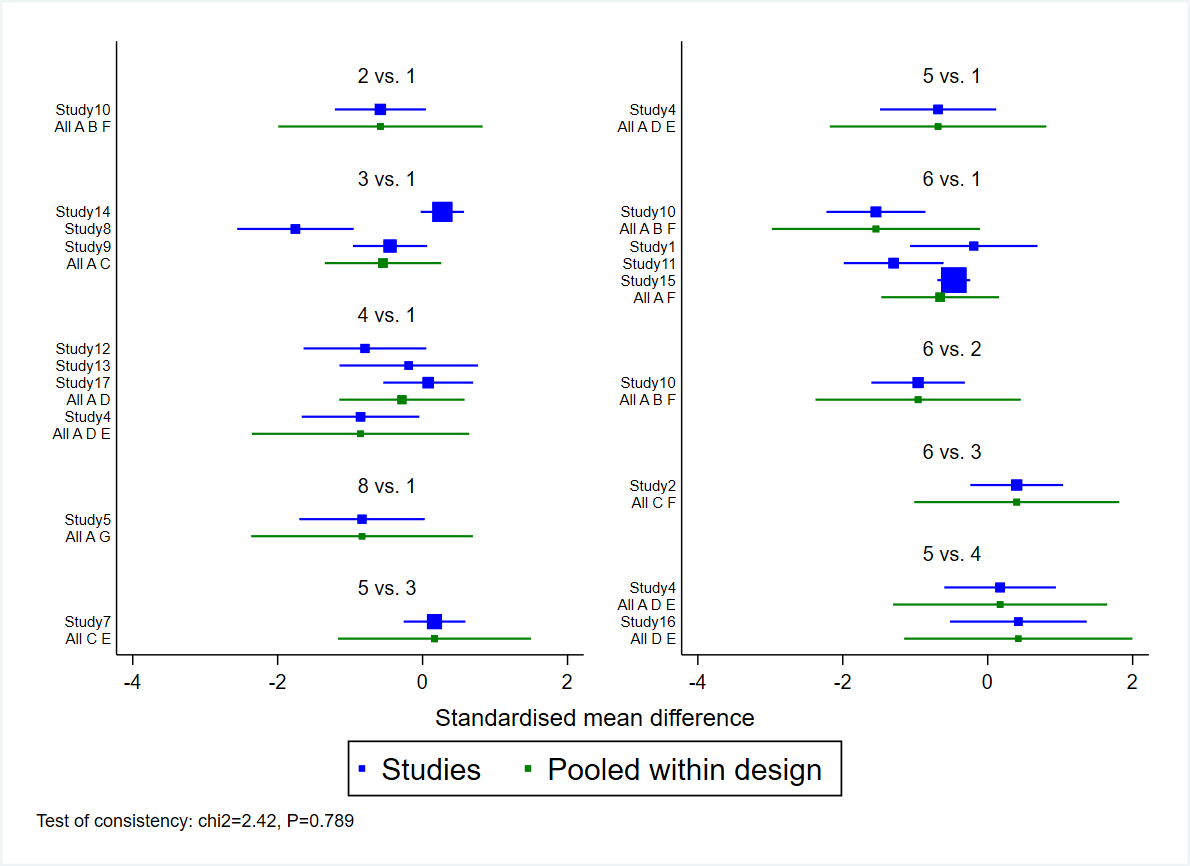

**Appendix 11-7** Waist circumference.

**Appendix 12.** Network plot comparing the effects of different exercise types on body fat percentage between recent (≤5 years) and earlier studies (>5 years).

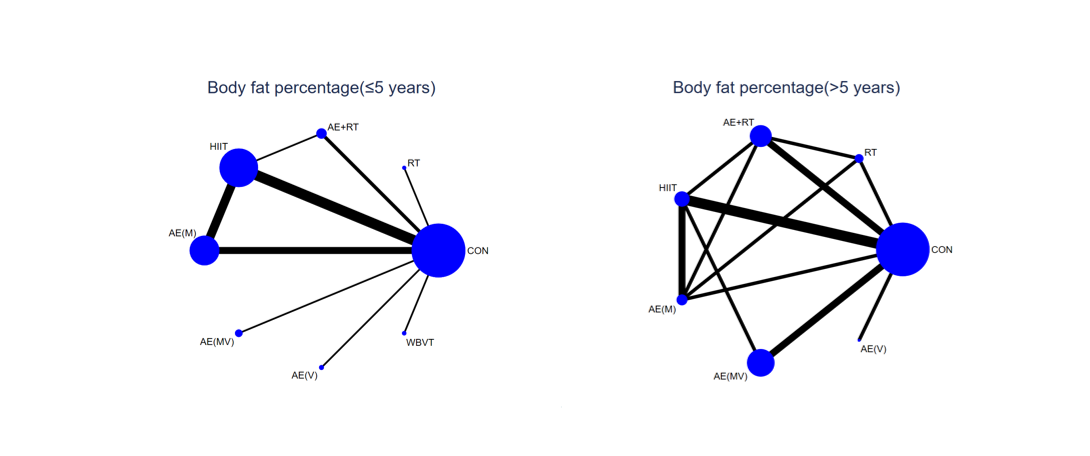

**Appendix 13.** CINeMA for the primary and secondary outcomes.

**1. Summary of study limitations of the included studies.**

The colours of the line indicate the summative ROB assessment of each comparison based on ROB assessment of each included studies (low ROB comparison [green], moderate ROB comparison [yellow] and high ROB comparison [red]).

Note: 1-CON, 2-RT, 3-AE + RT, 4-HIIT, 5-AE(M), 6-AE(MV), 7-AE(V), 8-WBVT.

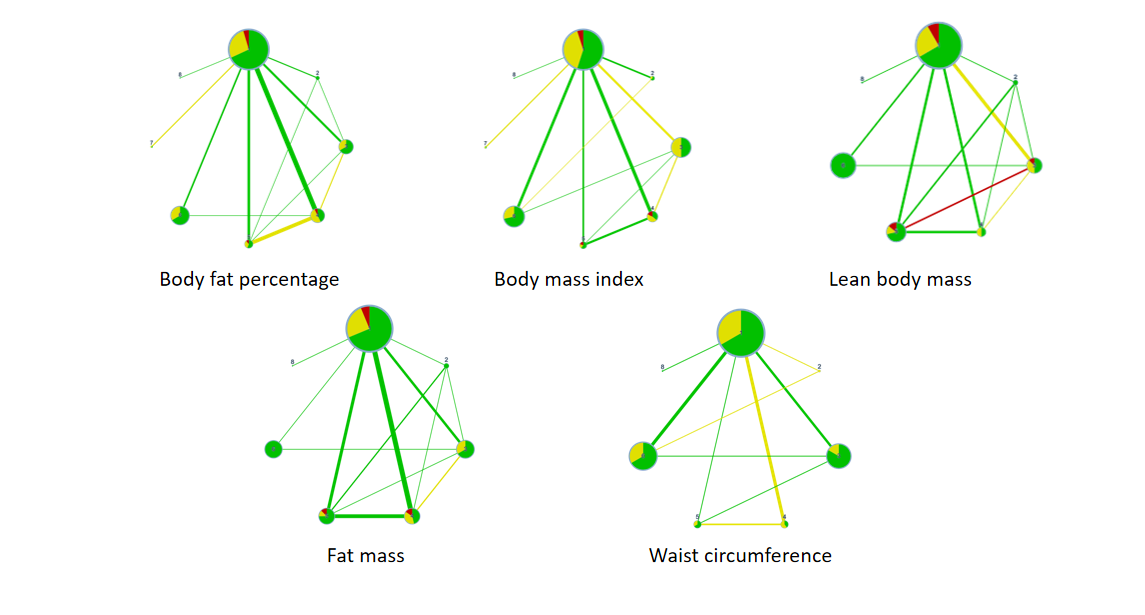

**2.Table of reasons for downgrading.**

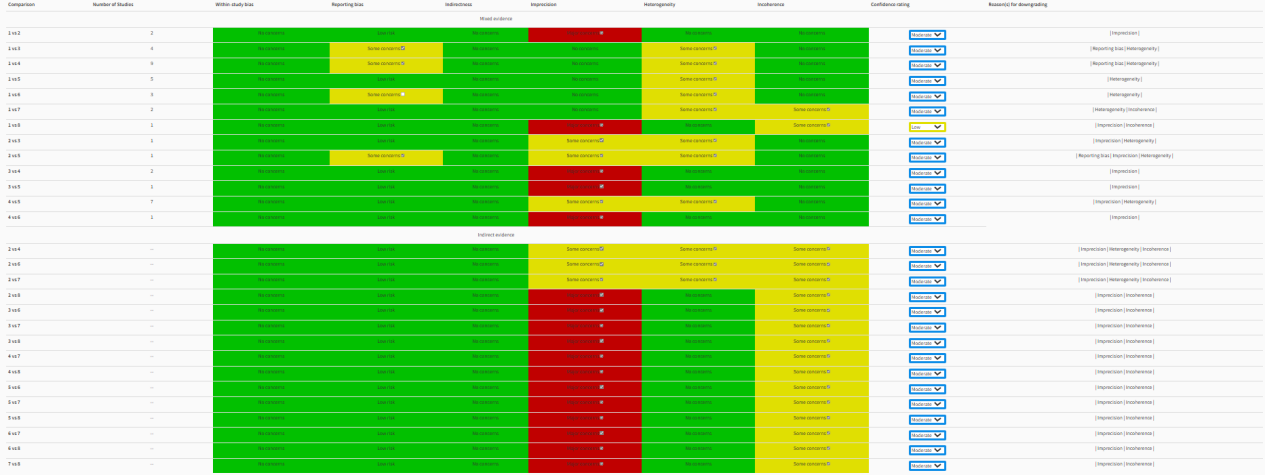

**Appendix 13-1** Body fat percentage.

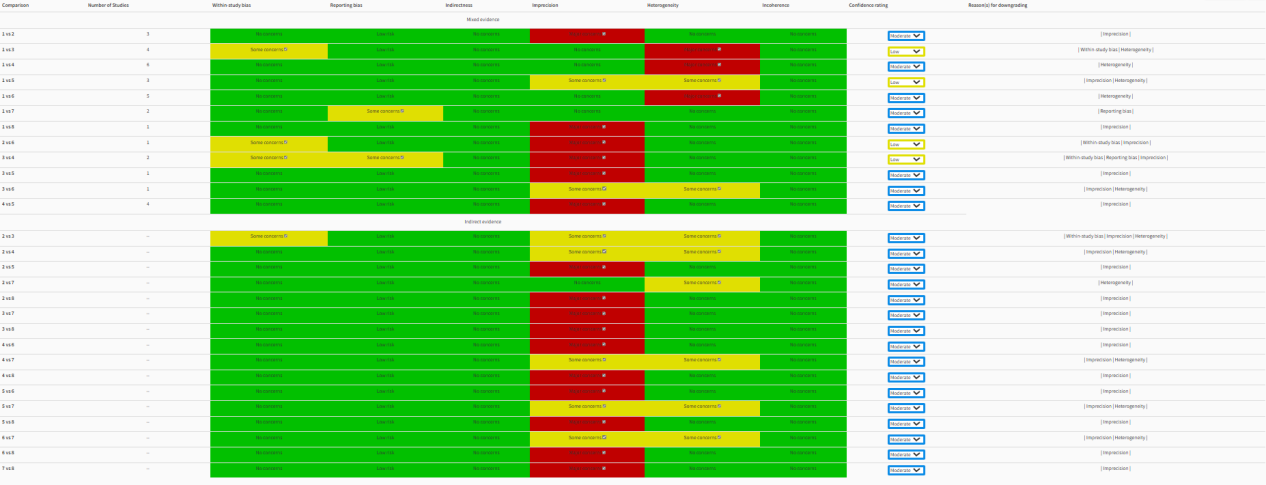
**Appendix 13-2** Body mass index.

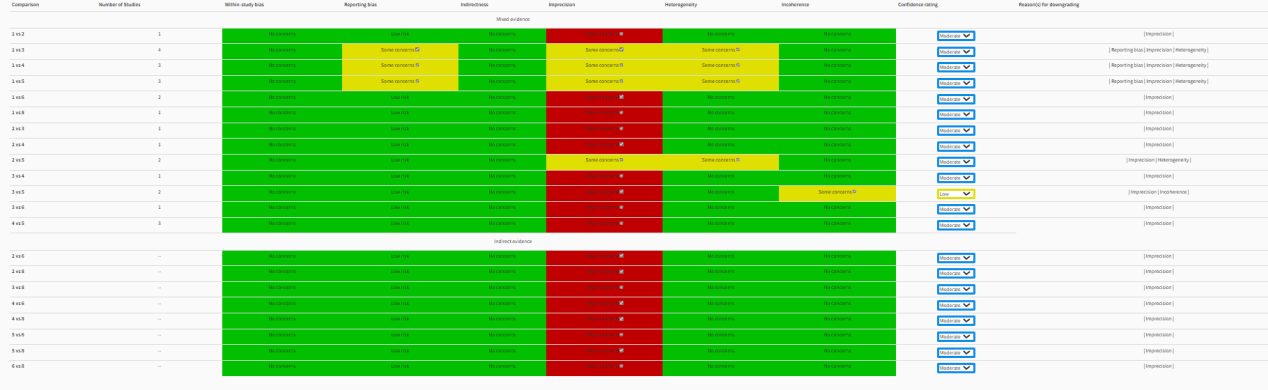

**Appendix 13-3** Lean body mass.

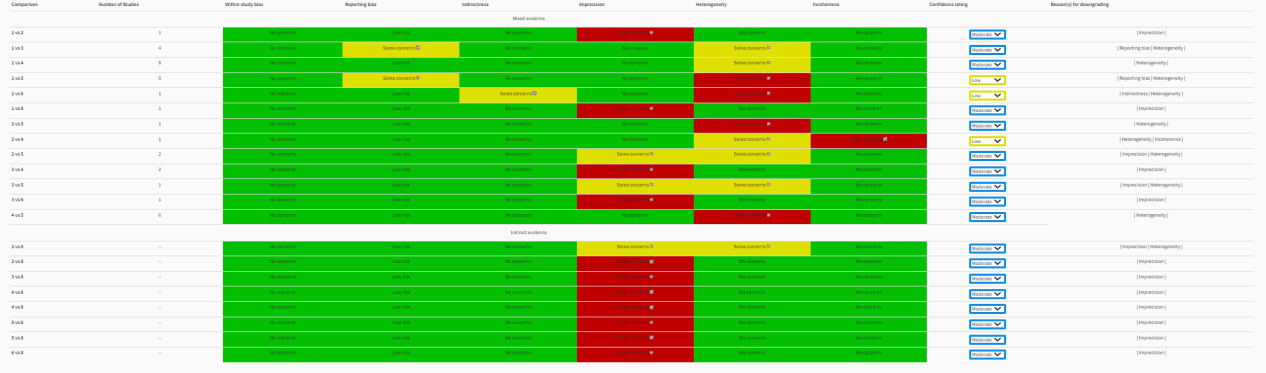

**Appendix 13-4** Fat mass.

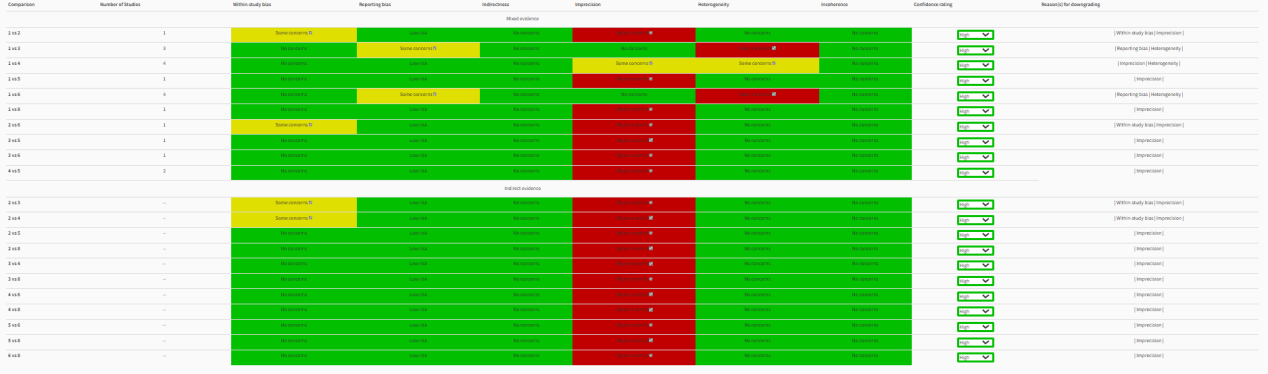

**Appendix 13-5** Waist circumference.

**Appendix 14.** Results of subgroup analysis of body fat percentage of studies utilizing imaging techniques.

| RT |  |  |  |  |  | **BFP(≤5 years)** |  |
| --- | --- | --- | --- | --- | --- | --- | --- |
| 1.12 (-0.88,3.13) | AE + RT |  |  |  |  |  |  |
| 0.95 (-0.86,2.76) | -0.17 (-1.29,0.94) | HIIT |  |  |  |  |  |
| 0.66 (-1.19,2.51) | -0.46 (-1.70,0.78) | -0.29 (-1.01,0.43) | AE(M) |  |  |  |  |
| 0.76 (-1.54,3.06) | -0.36 (-2.26,1.53) | -0.19 (-1.88,1.50) | 0.10 (-1.64,1.83) | AE(MV) |  |  |  |
| 1.65 (-0.75,4.05) | 0.53 (-1.49,2.54) | 0.70 (-1.13,2.52) | 0.99 (-0.88,2.85) | 0.89 (-1.42,3.20) | AE(V) |  |  |
| 0.71 (-1.67,3.10) | -0.41 (-2.40,1.59) | -0.23 (-2.04,1.57) | 0.05 (-1.79,1.90) | -0.04 (-2.34,2.25) | -0.93 (-3.33,1.46) | WBVT |  |
| -0.06 (-1.75,1.63) | **-1.18 (-2.26,-0.10)** | **-1.01 (-1.65,-0.36)** | -0.72 (-1.48,0.04) | -0.82 (-2.38,0.75) | -1.71 (-3.41,0.00) | -0.77 (-2.45,0.91) | CON |
| RT |  |  |  |  |  | **BFP(>5 years)** |  |
| 0.46 (-0.34,1.27) | AE + RT |  |  |  |  |  |  |
| 0.46 (-0.38,1.31) | 0.00 (-0.64,0.65) | HIIT |  |  |  |  |  |
| 0.57 (-0.29,1.43) | 0.11 (-0.64,0.85) | 0.10 (-0.58,0.78) | AE(M) |  |  |  |  |
| 0.54 (-0.41,1.49) | 0.08 (-0.72,0.87) | 0.07 (-0.62,0.77) | -0.03 (-0.90,0.85) | AE(MV) |  |  |  |
| 0.28 (-1.18,1.73) | -0.19 (-1.55,1.18) | -0.19 (-1.53,1.15) | -0.29 (-1.71,1.13) | -0.26 (-1.64,1.11) | AE(V) | WBVT |  |
| -0.32 (-1.08,0.43) | **-0.79 (-1.35,-0.22)** | **-0.79 (-1.31,-0.27)** | **-0.89 (-1.59,-0.19)** | **-0.86 (-1.47,-0.26)** | -0.60 (-1.84,0.64) | —— | CON |

AE: aerobic exercise, RT: resistance exercise, HIIT: high-intensity interval training, CON: control, WBVY: Whole body vibration training M: moderate intensity, V: vigorous intensity, MV: moderate to vigorous intensity, BFP: body fat percentage. Effects are expressed as the effect size (95% CI) between interventions. Bold indicates that the longitudinal intervention has a significantly reduction impact than the horizontal intervention.

Ranking of exercise interventions in order of effectiveness.

| Recent studies (≤5 years) | | Earlier studies (>5 years) | |
| --- | --- | --- | --- |
| Treatment | SUCRA | Treatment | SUCRA |
| CON | 12.5 | CON | 6.5 |
| RT | 24.1 | RT | 27.6 |
| AE+RT | 69.1 | AE+RT | 63.1 |
| HIIT | 63.1 | HIIT | 62.9 |
| AE(M) | 46.2 | AE(M) | 71.5 |
| AE(MV) | 52.3 | AE(MV) | 68.8 |
| AE(V) | 82.1 | AE(V) | 49.6 |
| WBVT | 50.5 | WBVT | —— |

AE: aerobic exercise, RT: resistance exercise, HIIT: high-intensity interval training, WBVT: Whole body vibration training, CON: control, M: moderate intensity, V: vigorous intensity, MV: moderate to vigorous intensity
